# Proteomic associations with fluctuation and long-term changes in BMI: A 40-year follow-up study

**DOI:** 10.1101/2025.01.28.25321236

**Authors:** Alvaro Obeso, Gabin Drouard, Teemu Palviainen, Xiaoling Wang, Miina Ollikainen, Karri Silventoinen, Jaakko Kaprio

## Abstract

**Introduction:** While some studies have explored associations between weight change and blood proteins, most have been intervention-based, offering limited insight into proteomic associations with long-term weight gain. It remains unclear whether plasma proteins are related to BMI fluctuation over time. This study investigates associations of long-term BMI changes and fluctuations with over 1,000 plasma proteins involved in cardiometabolic and inflammation functions.

**Data and Methods:** The study included 304 Finnish adult twins (117 men) born before 1958 from the Older Finnish Twin Cohort, with BMI data spanning five time points (1975, 1981, 1990, 2011, and 2012–2014). Proteomic data were derived from blood samples collected at the last BMI measurement. Linear mixed-effects models analyzed individual BMI trajectories, producing intercepts (baseline BMI) and slopes (BMI change rates). BMI fluctuation was calculated as the average squared deviation from expected BMI across time points. Associations between BMI changes/fluctuation and (i) 1,231 plasma proteins related to cardiometabolic and inflammatory functions and (ii) polygenic risk scores for BMI (PRS_BMI_), as well as interaction effects between PRS_BMI_ and baseline BMI on protein-BMI relationships were studied. Within-pair analyses using monozygotic twins were conducted to account for shared confounding factors.

**Results:** A total of 135 proteins were associated with changes in BMI over 40 years, while 17 proteins were linked to fluctuation in BMI: 12 associations (10 with BMI changes and 2 with fluctuation) remained significant in within-twin pair analyses. PRS_BMI_ associated with BMI changes but not with fluctuations. PRS_BMI_-protein interactions explaining BMI changes or fluctuation was found, though a single interaction between the CD72 protein and baseline BMI was observed.

**Conclusion:** This study highlights significant associations between plasma proteins and long-term BMI changes and fluctuations, with no evidence of PRS_BMI_-protein interactions influencing BMI trends. These findings underscore the substantial role of environmental factors in shaping proteome-BMI associations over adulthood.

## Introduction

Obesity is considered a major global public health challenge, with the proportion of adults with obesity more than doubling from 7% to 16% over the past three decades (1). While obesity is primarily caused by energy consumption exceeding energy expenditure, the underlying factors are multifaceted and influenced by environmental, behavioural, and genetic factors, as well as their interactions (2). Body Mass Index (BMI) has been widely used to classify overweight and obesity in epidemiological studies. However, obesity is a multifaceted concept, and the definition depends on whether it is used for epidemiological or clinical settings. In clinical settings, the definition of obesity needs to include other anthropometric and metabolic measures, emphasizing the need to understand how BMI is associated with other biological traits (3). Nonetheless, the measurement of BMI remains at the core of the diagnostic process and the results obtained based on BMI are highly likely to be transposable to obesity research since not only it has been seen that it present a high correlations with other measures of obesity such fat mass of the percentage of body fat (3), but also it has been reported associations between BMI and the development of obesity (4).

The proteome refers to a set of proteins produced by an organism or present within an organism or tissue at a given time. The proteome can be measured in various tissues, but it is most commonly examined in blood in human studies. The proteome is dynamic and intricate, undergoing changes over time in response to environmental factors that affect genome function, with genetic makeup also playing a significant role (5). Recent developments in proteomics research enable the examination of the connections between changes in BMI and protein levels.

The associations between plasma proteins and obesity have been extensively studied, with a number of proteins involved in cardiometabolic (6) and inflammatory processes identified (7, 8, 9). However, longitudinal proteomic studies are still rare, with the majority being interventions (10, 11, 12, 13). These studies have compared blood protein levels before and after weight loss, reporting various associations. Previous study have demonstrated that certain blood proteins initially fluctuated in response to the acute energy-deficient state, returning to their original levels once weight maintenance was achieved (11). Conversely, another group of proteins showed a decline in concentration during weight loss, with levels remaining stable during weight maintenance (11). It also showed that the associations between inflammatory and cardiovascular biomarkers and BMI levels before and during the weight loss process underwent a transformation, with the emergence of new markers associated with this period of weight loss (13). This suggests that blood proteins may exhibit disparate responses to short-term weight changes, which can be caused by dietary alterations. Some of these changes may reflect the weight loss process, while others may also reflect weight status. Additionally, weight loss typically involves a loss of both fat and lean body mass, while weight gain is primarily due to an accumulation of fat mass (16, 17, 18).

Therefore, the physiological and metabolic processes underlying weight gain and weight loss likely differ.

However, it is important to consider the time span of short-term weight loss vs weight gain over years or decades. Further investigation is needed to determine if associations between plasma proteins and long-term changes in BMI exist, and if these proteins are influenced by baseline weight. Finally, long-term studies of weight change have shown that individuals may lose weight in the short term but gain weight in the long term (19, 20). These fluctuations in BMI (i.e., BMI fluctuation over time) may be due to various reasons, including dieting and subsequent rebound. BMI fluctuation may also indicate a disease state, with erratic weight changes potentially indicating poor health condition For example, studies have shown negative effects of fluctuation in blood pressure and weight on coronary heart disease, coronary heart disease mortality and all-cause mortality (21, 22). To our knowledge, no study has examined how long-term fluctuation in BMI may be reflected at the proteome level.

BMI and its changes are influenced by genetic and environmental factors. Heritability estimates have ranged between 0.47-0.90 in twin studies and between 0.24–0.81 in family studies (23, 24, 25, 26). Additionally, genome-wide association studies (GWAS) have identified multiple genetic variants associating with BMI and its fluctuations during adulthood (27). These variants can be aggregated as polygenic risk scores (PRS), which can be used to estimate an individual’s genetic predisposition to traits such as BMI (28). Furthermore, some of these genetic associations are pleiotropic, meaning that they associate with multiple traits. Connections between proteins and genetic variants associated with BMI have been also identified (29, 30). In conclusion, while there is evidence of connections between genetic variants, BMI, and the proteome, the extent to which proteins associate with BMI changes over time remains underexplored.

To gain a deeper understanding of the associations between BMI changes and the proteome, we studied a sample of Finnish twins over a 40-year follow-up for whom plasma proteomic data was available. Additionally, we conducted within-pair analyses on monozygotic (MZ) twin pairs to investigate whether the identified associations persisted when controlling for shared environmental and genetic factors between the co-twins. Finally, an association analysis including interaction effects was carried out to study how genetic predisposition to BMI (PRS_BMI_) modulates the associations between BMI change and the proteome. This analysis reveals whether these associations are accentuated in individuals with a higher PRS_BMI_.

## Data and methods

### Cohort

The data was obtained from the Older Finnish Twin Cohort (FTC), a population-based study consisting of twins born before 1958 in Finland. The twins completed up to four survey questionnaires in 1975, 1981, 1990 and 2011 and reported their weight and height used to calculate BMI (kg/m^2^) (31). A fifth wave of data was derived from a subset of the twins who participated in the Essential Hypertension Epigenetics (EH-Epi) study. The EH-Epi study focused on twin pairs discordant for hypertension, so the sample was enriched for persons with high blood pressures and use of antihypertensive medications (32) During 2012 – 2014, research nurses conducted interviews and measured height, weight, and waist circumference. Fasting venous blood samples were collected from the EH-Epi twins at the mean age of 62 (age range: 55 – 69 years) to generate proteomic data. Participants with complete BMI measurements for all five waves were selected for the current analyses. The correlation between measured and self-reported BMI values was 0.95 for both men and women in the EH-Epi study showing high reliability of self-reported BMI (33). The research was conducted according to the principles of the Declaration of Helsinki, and the data collection was approved by the ethics committee of the Hjelt Institute, University of Helsinki and the ethics committee of the Helsinki and Uusimaa Hospital District, Finland. A linear regression model with inclusion/exclusion as a binary independent variable was conducted to assess the randomness of participant selection (Supplementary Table 1). While BMI was not a criterion for selection, participants had lower BMI in the latest surveys (3^rd^, 4^th^ and 5^th^ surveys) while in the earliest surveys (1^st^ and 2^nd^ surveys) their BMI was higher compared with the twins of the older Finnish Twin Cohort who were not selected for the current study. However, these differences were small and not statistically significant suggesting that the EH-Epi sample is broadly representative of the full cohort.

### Quantification of plasma Proteins

The proteome was measured from plasma samples of 415 twins from the EH-Epi using the Olink platform (Olink Explore 3072, Olink Proteomics AB, Uppsala, Sweden). A detailed description of the data and the pre-processing steps can be found elsewhere (6). Briefly, the data are presented as Normalized Protein eXpression (NPX) values, where NPX is Olink’s unit for quantifying relative protein concentrations on a log2 scale. Proteins detected in less than 80% of the samples were excluded. NPX values of remaining proteins that were below the limit of detection (LoD), representing less than 1% of all data points, were replaced with the LoD value of the corresponding plate. A small number of outlier samples were identified and excluded (N=14). Data were extracted from this sample for the current study participants, all of whom had complete proteomic data (n=304). In the current study, only proteins from the Cardiometabolic (I/II) and Inflammation (I/II) panels were used (736 and 737 proteins in Cardiometabolic and Inflammation panels, respectively). Protein descriptions are shown in the supplementary material (Supplementary Table 2).

### Polygenic Risk Score of BMI

PRS for BMI (PRS_BMI_) was calculated to test its interactions with changes and fluctuations in BMI during adulthood. The technical details of genotyping, imputation, and PRS_BMI_ calculations have been described elsewhere (34). The PRS_BMI_ was derived using GWAS summary statistics for BMI (27). The total number of single nucleotide polymorphisms (SNPs) used for the PRS_BMI_ calculations was 996,919. The number of individuals included in the GWAS was 692,578. The PRS_BMI_ was regressed against the top ten genetic principal components to correct for population stratification (35). The residuals were obtained and scaled to a mean of zero and unit variance, and subsequently used in relevant analyses without further processing. The distribution of PRS_BMI_ values for the sample used (consisting of individuals with BMI measurements from the five surveys conducted; N=304) was similar to the distribution of PRS_BMI_ values in the Older Finnish twin cohort (those not meeting the criteria for having BMI measurements from all five surveys; N=143) (see Supplementary Figures 1 and 2 for PRS_BMI_ of included and excluded samples).

### Calculation of changes in BMI

Linear mixed-effects (LME) modeling, i.e., models that include both fixed and random effects (36), was applied to longitudinal BMI data from the five surveys (1975, 1981, 1990, 2011 and 2012 – 2014) to calculate the linear trends and changes in BMI over time. The BMI trajectories were modelled with person identifiers as random effects to estimate baseline BMI (intercept) and BMI trend (slope) for each participant. The analyses were performed using R software (version 4.2.3) packages lme4 (version 1.1-34), lmerTest (version 3.1-3), modelsummary (version 1.4.3), dplyr (version 1.1.4) and optimx (version 2023-10.21).

### Calculation of the fluctuation in BMI

The expected BMI for each measurement point was calculated using the formula *y= xn+m*, where *n* is the change in BMI expressed as kg.m-2 per year (i.e. slope), *m* is the baseline BMI (i.e. BMI at first measure in 1975; age range 18 – 30 years) and *x* is the difference between the age of interest and the baseline age. Once the expected BMI values at each time point had been obtained, the difference between each of these expected BMI values and the corresponding observed BMI values was calculated. Finally, the differences between the observed and fitted BMI values were squared, and the resulting values were then averaged.

### Associations between proteins, changes or fluctuation in BMI and PRS_BMI_

LME models were used to investigate the associations between BMI changes and fluctuations with protein levels. This was achieved through the implementation of two distinct models.

We then assessed whether changes or fluctuations in BMI were associated with PRS_BMI._ The subsequent step was to examine interactions between protein levels and PRS_BMI_ in the case of a significant association in previous analyses (models 4 and 5, respectively). Next, interactions between protein levels and BMI at baseline (i.e., BMI at age range 18 – 30 years) were also examined.

Finally, we conducted sensitivity analyses. For analyses focusing on changes in BMI, we repeated them after excluding individuals with BMI > 30 kg/m^2^ in any of the surveys. This was done to investigate whether the identified associations were not limited to the participants with obesity. For analyses focusing on BMI fluctuation, BMI trajectories (i.e., slope and intercept) were incorporated as covariates into the models to assess whether the significant associations between BMI fluctuation and plasma proteins were attenuated when the BMI trajectories were taken into account.

All the analyses were performed using the R software (version 4.2.3) and the R package lme4 (version 1.1-34). The models used are displayed in the caption of the main and supplementary tables.

### Within-pair analysis

To assess the influence of environmental factors, and to rule out the effect of genetic differences on the significant associations between changes and fluctuation in BMI with proteins, within-pair analyses were conducted. These analyses were carried out using MZ twin pairs only (N of pairs = 50). Since MZ twins in a pair share, in practice, identical DNA sequence, any differences in their BMI, rate of BMI change, or BMI fluctuation suggest environmental influence. For example, if MZ twins who experience greater weight gain than their co-twins also exhibit higher levels of a specific protein, this association would likely be unrelated to genetic factors. We calculated within-pair differences (referred to with a Δ symbol below) between the changes in BMI, BMI fluctuation and the protein levels we previously identified as significantly associated with these two variables. We also calculated the mean of the baseline BMI values for each pair.

All the analyses were performed using the R software (version 4.2.3) and the R package lme4 (version 1.1-34) and the models used are displayed in the caption of the main and supplementary tables.

## Results

### Cohort and BMI trajectories

A total of 305 Finnish twins (118 men) with complete BMI and proteomic information were included in the current study. The mean BMI increased over the years in both men (mean of 5.27 kg/m²) and women (mean of 5.73 kg/m²). This corresponds to an annual increase of 0.13 kg/m^2^ in men and 0.14 kg/m^2^ in women, equating to around 400 g weight gain per year for a 170 cm tall person. We observed sex differences in BMI, height, and weight, as well as changes in BMI, from the earliest stages of the follow-up and maintained throughout (Table 1). Baseline BMI (i.e., intercept) and changes in BMI over the 40-year follow-up period (i.e., slope) were positively correlated in both men (r = 0.22, p-value = 0.01) and women (r = 0.23, p-value <0.01). If classified into BMI categories, most participants in wave 5 were of normal weight, with overweight being more common (19% in men and 29% in women) than obesity (11%) and normal weight in women (23%). The proportions of participants with prescribed antihypertensive medication were similar in men (57%) and women (56%). The longitudinal changes in BMI are illustrated in Figure 1.

**Figure 1:**
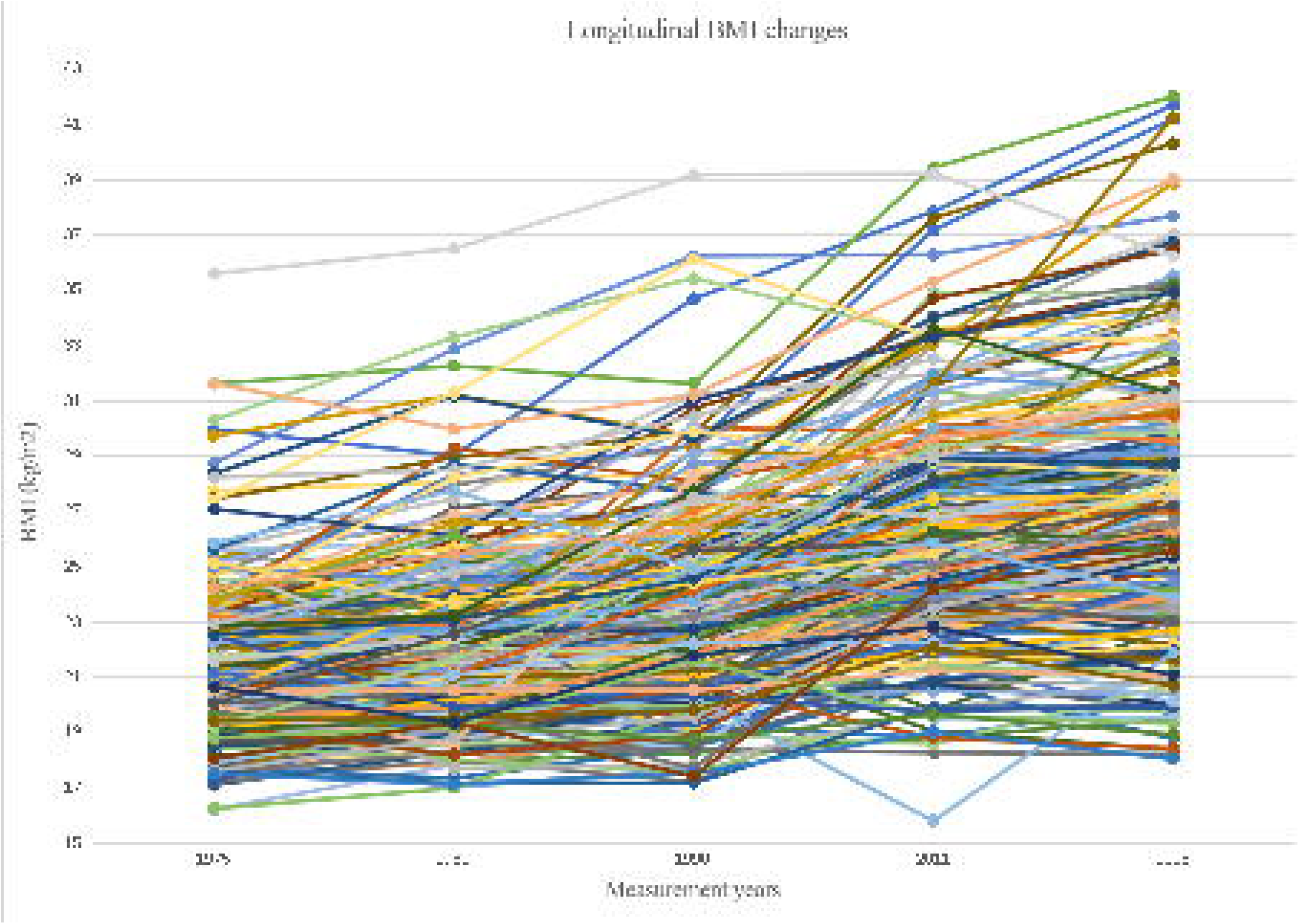
Spaghetti plot of individual BMI changes over time. Caption: BMI has been self-reported in 5 different waves comprising a 42-year period

**Table 1.** Descriptive and trajectories information of Older Finnish Twin Cohort by sex. Age, height, weight and BMI mean values for each wave among with BMI trajectories (i.e Slope and Intercept) are displayed along with the standard deviation, and the p value obtained from t-test to assess sex differences. **Abbreviations:** SD= standard deviation.

|  | Males (n=118) |  | Females (n=186) |  | p value |
| --- | --- | --- | --- | --- | --- |
|  | Mean | SD | Mean | SD |  |
| **Wave 1 (1975)** |  |  |  |  |  |
| Age | 24 | 3,65 | 24 | 3,73 | 0,57 |
| Height | 1,78 | 0,06 | 1,63 | 0,05 | <2,00E-16 |
| Weight | 70 | 10,82 | 55 | 6,9 | <2,00E-16 |
| BMI | 22,34 | 2,84 | 20,24 | 2,53 | 1,10E-09 |
| **Wave 2 (1981)** |  |  |  |  |  |
| Age | 30,11 | 3,6 | 30,13 | 3,75 | 0,95 |
| Height | 1,78 | 0,06 | 1,63 | 0,05 | <2,00E-16 |
| Weight | 75,5 | 11,67 | 56 | 7,18 | <2,00E-16 |
| BMI | 23,63 | 3,1 | 21,06 | 2,79 | 3,41E-09 |
| **Wave 3 (1990)** |  |  |  |  |  |
| Age | 39,34 | 3,54 | 38,92 | 3,75 | 0,22 |
| Height | 1,78 | 0,06 | 1,63 | 0,05 | <2,00E-16 |
| Weight | 80 | 13,01 | 60 | 8,82 | <2,00E-16 |
| BMI | 24,89 | 3,5 | 22,22 | 3,39 | 1,92E-10 |
| **Wave 4 (2011)** |  |  |  |  |  |
| Age | 60,15 | 3,59 | 60,14 | 3,81 | 0,86 |
| Height | 1,78 | 0,06 | 1,62 | 0,05 | <2,00E-16 |
| Weight | 84,5 | 14,44 | 67 | 11,43 | <2,00E-16 |
| BMI | 26,75 | 4,01 | 24,91 | 4,31 | 3,78E-05 |
| **Wave 5 (2015)** |  |  |  |  |  |
| Age | 62,19 | 3,83 | 62,41 | 3,83 | 0,08 |
| Height | 1,77 | 0,07 | 1,61 | 0,05 | <2,00E-16 |
| Weight | 86,25 | 15,27 | 68 | 12,96 | <2,00E-16 |
| BMI | 27,61 | 4,51 | 25,97 | 5,11 | 4,34E-03 |
| **BMI trajectories** |  |  |  |  |  |
| Slope | 0,13 | 0,06 | 0,14 | 0,08 | 4,28E-04 |
| Intercept | 22,54 | 2,84 | 20,73 | 2,53 | 4,79E-08 |

### Associations between proteins and changes in BMI

After correcting significance levels for multiple testing using the Bonferroni method (number of tests=1231, corrected p-value= 4,06 e-05), a total of 135 out of 1231 proteins were significantly associated with changes in BMI: 73/736 proteins belonged to the cardiometabolic panel and 62/737 proteins to the inflammation panel. A negative association with changes in BMI was observed for 31 proteins, while a positive association was observed for 104 proteins (Supplementary Table 3). The strongest negative association was observed with the protein apolipoprotein F (coefficient:-0.14; p=5.88e-10), indicating that one unit increase in apolipoprotein F expression was associated with a 0.14 kg/m^2^ decrease in changes in BMI per year. The strongest positive association was observed between changes in BMI and the expression of the pigment epithelium-derived factor (coefficient: 0.18; p=1.86e-11), indicating that one unit increase in pigment epithelium-derived factor expression was associated with 0.18 kg/m^2^ decrease in BMI changes per year. The full summary is available in the supplementary material (Supplementary Table 3).

In the sensitivity analyses excluding participants with BMI values greater than 30 kg/m² in any of the surveys, the number of proteins associated with changes in BMI decreased to 40.

Approximately half of these proteins belonged to the cardiometabolic panel and the other half to the inflammatory panel (19 and 21 proteins respectively). Pigment-derived factor remained the protein showing the strongest positive association with changes in BMI (coefficient: 0.11; Bonferroni p=9.90e-04), while serum paraoxonase/lactonase 3 was the protein with the strongest negative association (coefficient:-0.09; Bonferroni p=8.41e-05) (Supplementary Table 4).

### Associations between proteins and fluctuation of BMI

Fluctuation of BMI over time was associated with 17 proteins: 10 belonged to the inflammation panel and 7 to the cardiometabolic panel (Table 2). All proteins were found to be positively associated with BMI fluctuation, with the strongest association observed for interleukin-10 receptor subunit beta (coefficient: 1.50; p=1.61e-02). In a sensitivity analysis, where BMI slope and BMI intercept were included as covariates, the associations were non-significant (Supplementary Table 5).

**Table 2:** Proteins showing a significant association with fluctuation in BMI after carrying out simple linear mixed effects. Significant association results displayed from the use of LME model: BMI_fluctuation ∼ Sex + Age at blood sample + Protein + (1|Family ID) with the fluctuations in BMI as an outcome, the protein level, sex and the age when the blood sample was taken as fixed effects. Estimate, standard error, nominal p value and p value after Bonferroni correction are displayed for the proteins (characterised by the protein ID, protein description and belonging panel) that showed significant association with BMI fluctuation. **Abbreviations:** SE: Standard error.

| Protein Description | Protein ID | Estimate | SE | p value |  | Protein panel |
| --- | --- | --- | --- | --- | --- | --- |
|  |  |  |  | Nominal | Bonferroni |  |
| Leptin | P41159 | 0,46 | 0,08 | 2,30E-08 | 2,84E-05 | Cardiometabolic |
| Interleukin-1 receptor antagonist protein | P18510 | 0,7 | 0,12 | 2,41E-08 | 2,97E-05 | Inflammation |
| Fatty acid-binding protein, adipocyte | P15090 | 0,8 | 0,15 | 3,61E-07 | 4,45E-04 | Cardiometabolic |
| Angiopoietin-related protein 4 | Q9BY76 | 1,13 | 0,23 | 9,69E-07 | 1,19E-03 | Inflammation |
| Disintegrin and metalloproteinase domain-containing protein 12 | O43184 | 0,94 | 0,19 | 1,99E-06 | 2,45E-03 | Inflammation |
| Na(+)/H(+) exchange regulatory cofactor NHE-RF3 | Q5T2W1 | 0,59 | 0,13 | 3,55E-06 | 4,37E-03 | Inflammation |
| Growth/differentiation factor 15 | Q99988 | 1,03 | 0,22 | 3,85E-06 | 4,74E-03 | Cardiometabolic |
| Bile salt sulfotransferase | Q06520 | 0,51 | 0,11 | 8,00E-06 | 9,86E-03 | Inflammation |
| 2-iminobutanoate/2-iminopropanoate deaminase | P52758 | 0,62 | 0,14 | 8,24E-06 | 1,02E-02 | Inflammation |
| Coiled-coil domain-containing protein 80 | Q76M96 | 1,03 | 0,23 | 8,33E-06 | 1,03E-02 | Cardiometabolic |
| Interleukin-10 receptor subunit beta | Q08334 | 1,5 | 0,34 | 1,30E-05 | 1,61E-02 | Inflammation |
| Aflatoxin B1 aldehyde reductase member 4 | Q8NHP1 | 0,43 | 0,1 | 1,41E-05 | 1,74E-02 | Inflammation |
| Pterin-4-alpha-carbinolamine dehydratase | P61457 | 0,67 | 0,15 | 1,71E-05 | 2,11E-02 | Inflammation |
| Scavenger receptor cysteine-rich type 1 protein M130 | Q86VB7 | 0,85 | 0,19 | 1,74E-05 | 2,14E-02 | Cardiometabolic |
| Pantetheinase | O95497 | 0,53 | 0,12 | 2,05E-05 | 2,52E-02 | Inflammation |
| Glutathione S-transferase A1 | P08263 | 0,34 | 0,08 | 2,27E-05 | 2,79E-02 | Cardiometabolic |
| E-selectin | P16581 | 0,68 | 0,16 | 3,60E-05 | 4,43E-02 | Cardiometabolic |

### Associations between PRS_BMI_ with BMI changes and fluctuation of BMI

The associations between changes and fluctuations in BMI and the PRS_BMI_ were quantified using LME models. PRS_BMI_ was positively associated with changes in BMI (estimate: 0.01, p-value = 0.01) but not with fluctuation in BMI (Supplementary Table 6).

### BMI changes-Protein associations with interactions

After introducing an interaction term between PRS_BMI_ and proteins associated with changes in BMI, no significant interaction was found. However, when investigating whether baseline BMI and protein levels interact in predicting changes in BMI, one significant interaction was identified (Table 3 and Figure 2). This interaction involved the B cell differentiation antigen CD72 protein with a positive estimate of 0.01 (Bonferroni p= 0.05). We did not perform interaction analyses on BMI fluctuation, as we did not identify any associations between BMI fluctuation and PRS_BMI_.

**Figure 2:**
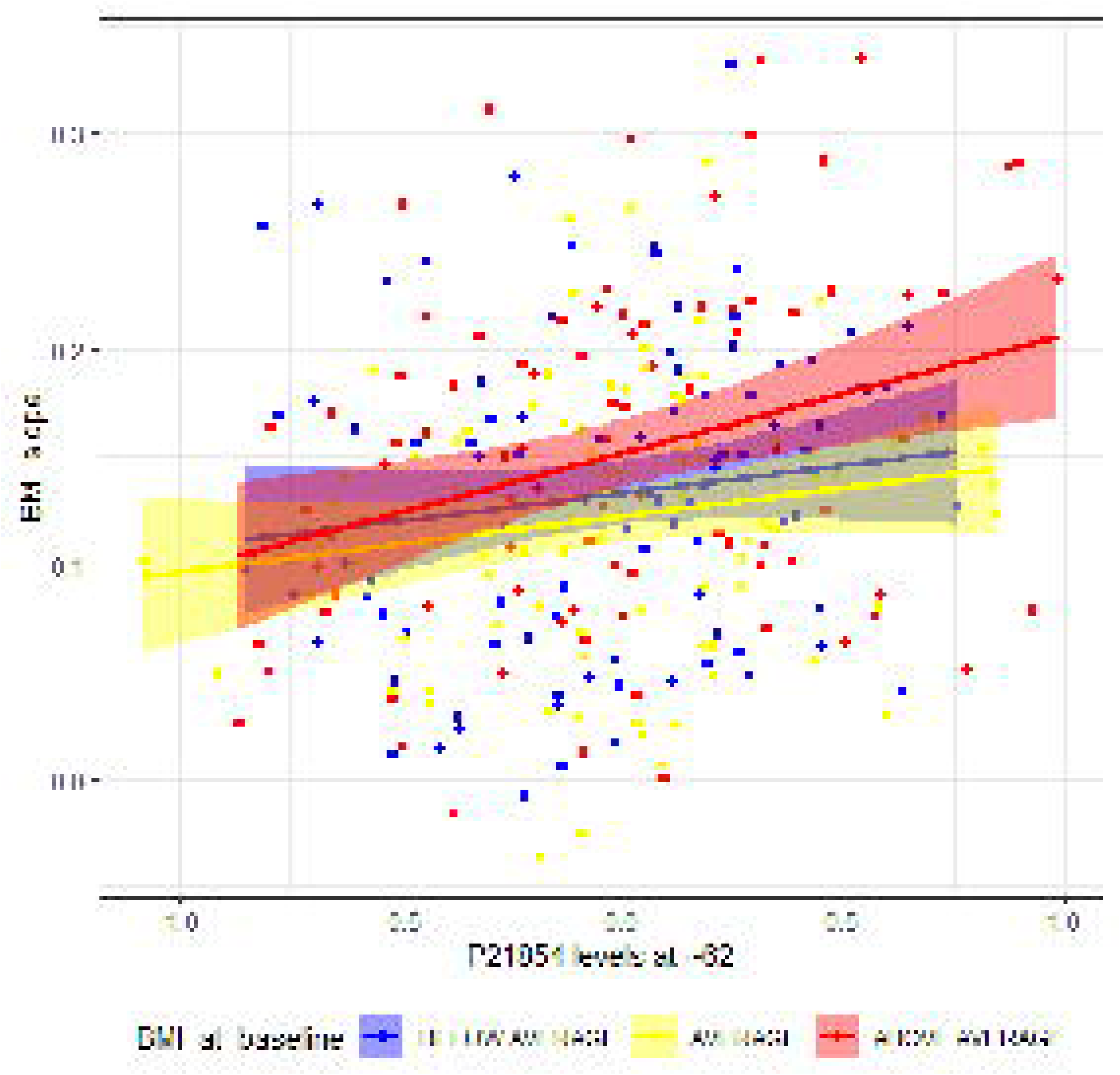
Graphical illustration of interactions between plasma protein levels with BMI baseline in predicting BMI changes during adulthood. Graphical illustration of the association between changes in BMI during adulthood (from ∼24 to ∼62 years old of age) and the interaction between protein B cell differentiation antigen CD72 and BMI baseline (at ∼24 years old) (interaction coefficient: 0.01, p=0.05)

**Table 3:** Linear mixed-effects models to assess associations of BMI changes with interactions between the BMI baseline and the proteins previously appeared to be significantly associated with the changes in BMI. Significant association results displayed from the use of LME model: Change_BMI ∼ Sex + Age at blood sample + Protein + Protein:Baseline_BMI + Baseline_BMI + (1|Family ID) where changes in BMI is the outcome and the protein level at PRS∼62 years old, the age of blood sampling the baseline BMI (∼24 years old) and the interaction between protein level and the baseline BMI were fixed effects. **Abbreviations:** SE: Standard error.

| Protein description | BMI baseline |  |  |  |
| --- | --- | --- | --- | --- |
|  | Estimate | SE | p value | Bonferroni |
| B-cell differentiation antigen CD72 | 0,004 | 0,001 | 7,47E-03 | 1 |

### Within-pair analyses

Ten proteins associated with BMI change showed significant associations within MZ twin pairs: Seven were identified as part of the cardiometabolic panel (Leptin, High affinity immunoglobulin alpha and immunoglobulin mu Fc receptor, Insulin-like growth factor-binding protein 2, Creatine kinase B-type, Somatotropin, BPI fold-containing family B member 1 and Insulin-like growth factor-binding protein 1) and three as part of the inflammation panel (Apolipoprotein F, Growth hormone receptor and Ectonucleotide pyrophosphatase/ phosphodiesterase family member 7) (Supplementary Table 7). The protein with the most robust positive association was growth hormone receptor (coefficient: 0.14; p=4.00e-3), while the one with the most pronounced negative association was apolipoprotein F (coefficient:-0.31; p=1.61e-3). Only two of the 17 proteins associated with BMI fluctuation showed significant associations in within-pair analyses (see Supplementary Table 8). These proteins were E-selectin and Na(+)/H(+) exchange regulatory cofactor NHE-RF3 protein.

## Discussion

The current study aimed to investigate (i) the associations between changes in BMI and BMI fluctuations over time during adulthood with plasma proteins, (ii) whether these associations persisted after controlling for shared confounding between MZ co-twins, and (iii) whether the associations between changes and fluctuation in BMI with protein levels are modulated by PRS_BMI_. Overall, 135 proteins were identified to associate with changes in BMI, while 17 were associated with fluctuation in BMI. When conducting within-pair analyses in MZ pairs, 10 and 2 proteins remained significantly associated with BMI changes and BMI fluctuation, respectively. These results suggest that genetic factors are not the sole drivers of the observed associations, but rather highlight the importance of the environment not shared by co-twins. In addition, PRS_BMI_ was significantly associated with BMI changes. Nevertheless, no significant interactions between PRS_BMI_ with protein levels in explaining changes in BMI were observed. However, we identified one significant interaction between baseline BMI and the B cell differentiation antigen CD72 protein in explaining changes in BMI. This indicates that the association between the cell differentiation antigen CD72 protein and changes in BMI was stronger in individuals whose baseline BMI was high, independent of the direct effect of baseline BMI on BMI change.

Several proteins were identified as being associated with BMI changes during adulthood. Some of these proteins, including those belonging to the complement factors (specifically complement factor I, B, D, and H), sex hormone binding protein (SHBP), galectin-3 binding protein (Gal-3BP), afamin, and antithrombin III, have been documented in intervention studies focusing on proteome changes after weight loss in individuals with overweight or obesity as a starting point (11, 12, 13, 14). The direction of associations in these intervention studies was consistent with our results. For instance, in several studies, the authors observed that complement factors, Gal-3BP, and afamin were negatively associated, while SHBP and antithrombin III were positively associated with weight loss (11, 12, 13, 14). In contrast, our study found opposite associations. This discrepancy can be attributed to our study design, which primarily focused on overall weight gain in the general population, rather than weight loss seen in these intervention studies. Our findings are thus in line with existing literature indicating that these proteins covary with BMI changes in the same direction.

Other proteins, such as insulin-like growth factor binding proteins (*IGFBP1* and *IGFBP2*) and serum amyloid P, have also been reported in the literature on weight loss, although with less frequency. In the case of IGFBPs, previously published studies demonstrated a positive association between these proteins and weight loss (13, 14). Our findings indicate negative correlations between these proteins and changes in BMI. Regarding serum amyloid P, previous studies reported a negative association between this protein and weight loss (11, 14). However, we found a positive association with changes in BMI. In both cases (the IGFBP and Serum amyloid P), the results are consistent, as the direction of BMI change aligns.

Finally, when comparing our results with previously longitudinal study which examined plasma proteome changes over a 10-year period, reported several proteins identified in the current study as also associated with changes in BMI over a period of 40 years (37). These proteins include leptin, tissue-type plasminogen activator, cathepsin D, and hepatocyte growth factor (HGF), all of which exhibited a positive association with BMI changes(37). This is consistent with the findings of the current study indicating that these proteins are likely good candidate biomarkers for long-term weight gain in adults. Additionally, another longitudinal study previously conducted where the long-term associations between BMI changes during adolescence and protein levels was examines, found some complement factors (I, B and H), serum amyloid P component, and SHBP to be associated with the BMI changes (38). Those proteins exhibited similar patterns of association to those observed in the current study. As weight gain in adults is mainly in fat mass, this suggests that the above proteins may be associated with changes in body fat rather than lean mass during adolescence.

We identified several proteins associated with BMI fluctuations. Most of them (except for fatty acid-binding protein, adipocyte, 2-iminobutanoate/2-iminopropanoate deaminase, aflatoxin B1 aldehyde reductase member 4 and pantetheinase), were previously reported to be associated with weight loss (14). The majority of the proteins showed a negative association with weight loss, except for Na(+)/H(+) exchange regulatory cofactor NHE-RF3, Coiled-coil domain-containing protein 80 and pterin-4-alpha-carbinolamine dehydratase, which exhibited a positive association. Previously published intervention study also identified a negative association between pantetheinase and weight loss (13). Additionally, a few of these proteins (leptin and E-selectin) were identified as positively associated with long-term changes in BMI in the previously published study (14). However, to the best of our knowledge, no previous studies have examined the associations between longitudinal BMI fluctuation and protein levels, which hinders our ability to compare our results with existing literature on BMI fluctuation. Further studies investigating the proteomic associations with BMI fluctuation are warranted.

From the previously identified associations between proteins with changes or fluctuation in BMI, several remained significant in within-pair analyses conducted in MZ twin pairs. This suggests that some associations between protein levels and BMI trajectories persist when controlling for genetic factors, indicating either an environmental influence on these associations or possible causal relationships. Environmental correlations between protein levels and BMI as well as changes in BMI during adolescence have been previously reported (35), and our study complements the literature with a longer follow-up over adulthood.

Additionally, to our knowledge, no previous study has investigated the environmental effects underlying associations between the plasma proteome and fluctuations in BMI, and we demonstrate that two proteins are associated (Na(+)/H(+) exchange regulatory cofactor NHE- RF3 and E-selectin) with fluctuations in BMI independently of genetic factors.

Finally, only one interaction between baseline BMI and protein levels was positive and significant in predicting changes in BMI. This interaction involved the protein B cell differentiation antigen CD72. In individuals with the same protein level, those with a higher BMI at baseline gained more BMI during adulthood, in addition to the individual effect of baseline BMI on changes in BMI. To the best of our knowledge, no previous studies have reported the B cell differentiation antigen CD72 protein to be associated with changes in BMI alone, or to interact with baseline BMI.

The current study has several strengths. The longitudinal data derived from a twin cohort with five BMI measurements enabled the characterization of BMI trajectories with a high level of depth. To date, and to the best of our knowledge, our study represents the longest follow-up period of any proteomic study investigating changes and fluctuations in BMI. Moreover, the data used in the study is broadly representative of the general population, even though the sample is enriched for persons with hypertension However, it is important to note that our study faces limitations. The most significant limitation is the relatively small sample size, which may have reduced the statistical power of the study and increased the uncertainty of the estimates obtained. Furthermore, since protein levels were measured only once, we could not study temporal changes in protein expression. Finally, BMI is a measure that is less informative about a person’s obesity status than other measures such as body fat mass or adiposity. Nevertheless, the collection of more sophisticated measurements of body fat is challenging within the scope of long-term observational studies.

## Conclusion

The current study identified numerous associations between long-term changes and fluctuation in BMI with various cardiometabolic and inflammation-related protein levels. These proteins may be of interest to identify individuals whose BMI may increase sharply over time. Finally, our findings suggest that genetic factors are not the only drivers of the associations between the blood plasma proteome and BMI trajectories but highlight the role of the environment in shaping the long-term BMI change and its link with older age proteome. While this study focused on adults, more longitudinal studies of weight change and the associated proteins in children and adolescents are needed. Follow-up studies starting at earlier ages would enable the identification of proteins that could serve as markers for monitoring and preventing the development of obesity from a young age.

## Data Availability Statement

The Finnish Twin Cohort data used in the analysis is deposited in the Biobank of the Finnish Institute for Health and Welfare (https://thl.fi/en/web/thl-biobank/forresearchers). It is available to researchers after written application and following the relevant Finnish legislation.

## Code availability

All R scripts are available from the corresponding authors.

## Data Availability

Data Availability Statement: The Finnish Twin Cohort data used in the analysis is deposited in the Biobank of the Finnish Institute for Health and Welfare (https://thl.fi/en/web/thl-biobank/forresearchers). It is available to researchers after written application and following the relevant Finnish legislation.

## Acknowledgements

Not applicable.

## Author contributions

The study design was developed and discussed by AO, GD and JK. The statistical analyses were performed by AO. The phenotype data was processed by AO and proteomic data processing was performed by GD. JK, XW, MO, participated in the data collection. Polygenic risk scores were calculated by TP. AO wrote the original manuscript.

AO, GD, JK, XW, MO, TP and KS participated in the improvement of the manuscript by critically revising it and read and approved the final version of the manuscript.

---

**Supplementary Figure 1:**
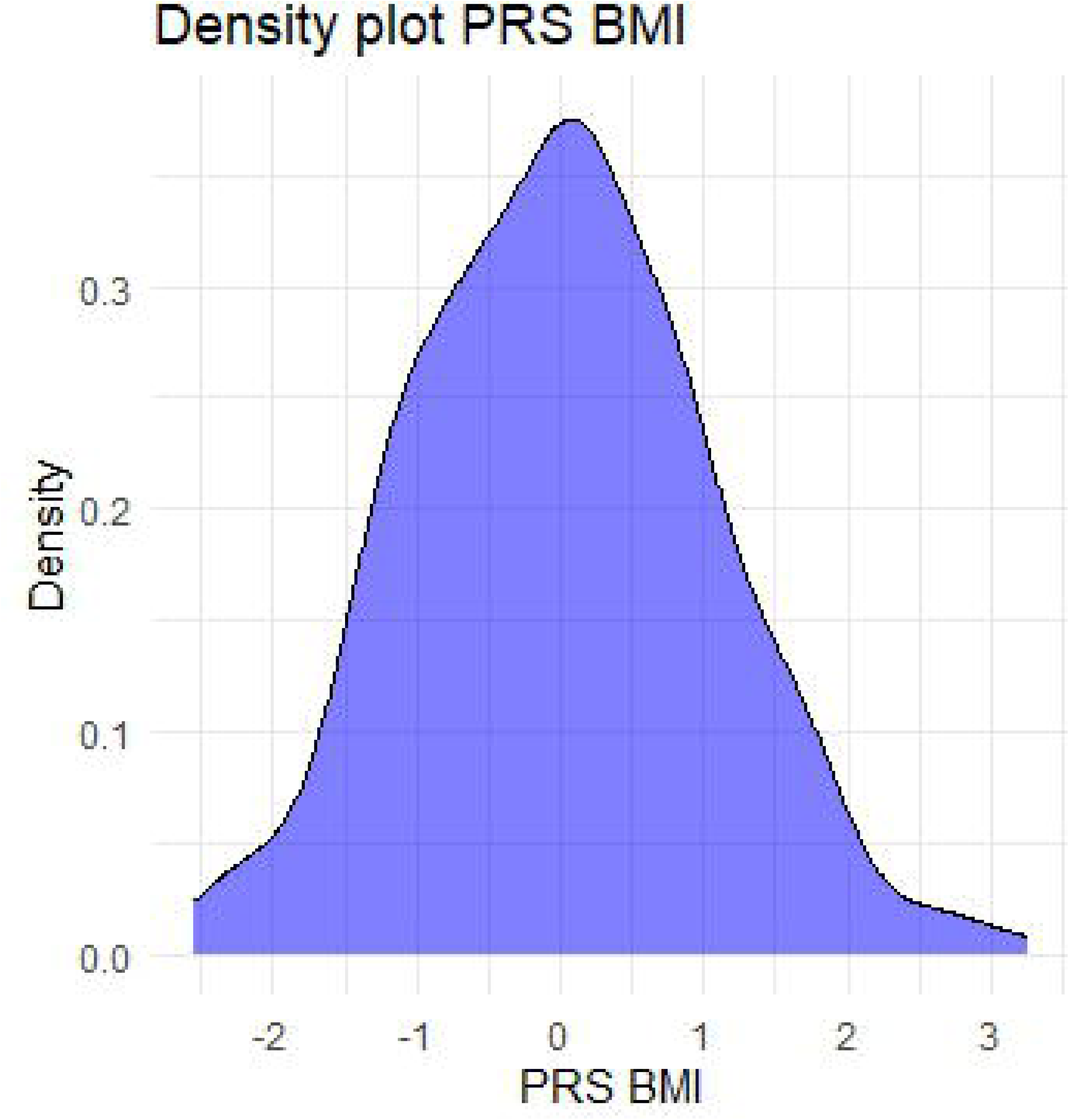
Graphical illustration of the distribution of Polygenic risk score of body mass index of the samples included in the study (N=305). **Caption:** Density plot of the Polygenic risk score of BMI of the individuals of the older Finnish twin cohort included in the study (N=305). **Abbreviations:** BMI: Body mass index; PRS: Polygenic risk score.

**Supplementary Figure 2:**
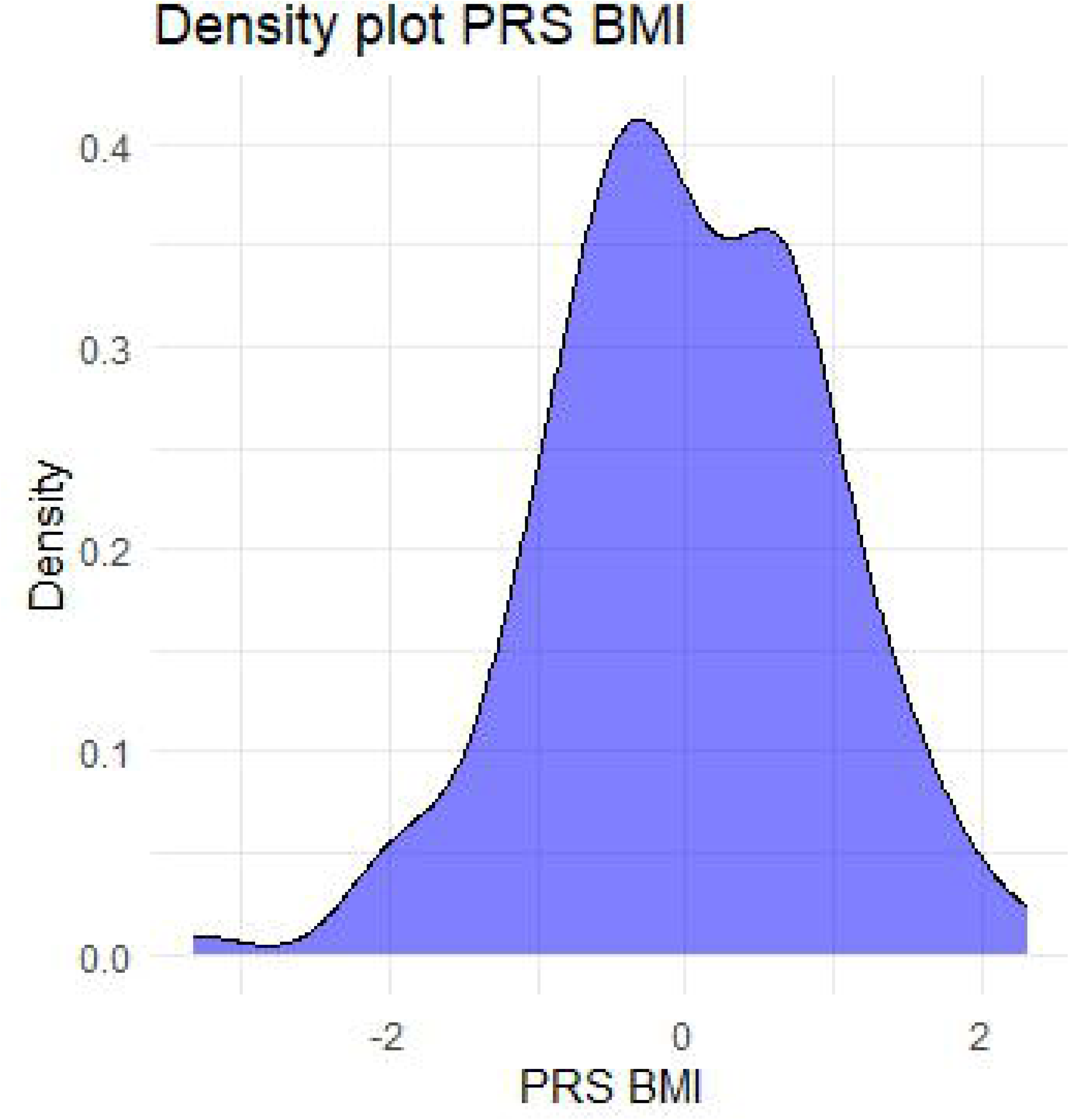
Graphical illustration of the distribution of Polygenic risk score of body mass index of the sample not included in the study (N=175). **Caption:** Density plot of the Polygenic risk score of BMI of the individuals of the older Finnish twin cohort not included in the study (N=175). **Abbreviations:** BMI: Body mass index; PRS: Polygenic risk score.

