## Supplementary tables for "Proteomic associations with fluctuation and long-term changes in BMI: A 40-year follow-up study"

**Supplementary table 1:** Linear regression model to assess selection bias based on the Body masss index.

| Linear Regression results | | | | |
| --- | --- | --- | --- | --- |
| BMI measure | Independent Variable | Estimate (kg/m2) | p value | R-squared |
| BMI 1975 | Inclusion | 0,10 | 0,72 | 0,16 |
| BMI 1981 | Inclusion | 0,35 | 0,33 | 0,17 |
| BMI 1990 | Inclusion | -1,34 | 0,08 | 0,12 |
| BMI 2011 | Inclusion | -0,53 | 0,27 | 0,03 |
| BMI 2015 | Inclusion | -0,62 | 0,26 | 0,01 |

**Caption:** Linear regression results from all the surveys with inclusion/exclusion as independent variable, with the estimate, the p value and the R^2^ values displayed. **Abbreviations:** BMI: Body mass index.

**Supplementary table 2**: List of proteins used in the current study obtained from Olink® Explore.

| **UniProt ID** | **Protein name** | **Gene name** | **Explore 384 panel** |
| --- | --- | --- | --- |
| P16860 | Natriuretic peptides B | NPPB | Cardiometabolic |
| P19429 | Troponin I, cardiac muscle | TNNI3 | Cardiometabolic |
| P61978 | Heterogeneous nuclear ribonucleoprotein K | HNRNPK | Cardiometabolic |
| P17676 | CCAAT/enhancer-binding protein beta | CEBPB | Cardiometabolic |
| P08670 | Vimentin | VIM | Cardiometabolic |
| O96017 | Serine/threonine-protein kinase Chk2 | CHEK2 | Cardiometabolic |
| P34998 | Corticotropin-releasing factor receptor 1 | CRHR1 | Cardiometabolic |
| Q969D9 | Thymic stromal lymphopoietin | TSLP | Cardiometabolic |
| P55082 | Microfibril-associated glycoprotein 3 | MFAP3 | Cardiometabolic |
| O95988 | T-cell leukemia/lymphoma protein 1B | TCL1B | Cardiometabolic |
| Q9NRD8 | Dual oxidase 2 | DUOX2 | Cardiometabolic |
| Q16619 | Cardiotrophin-1 | CTF1 | Cardiometabolic |
| P17516 | Aldo-keto reductase family 1 member C4 | AKR1C4 | Cardiometabolic |
| Q9NRV9 | Heme-binding protein 1 | HEBP1 | Cardiometabolic |
| P36952 | Serpin B5 | SERPINB5 | Cardiometabolic |
| P52789 | Hexokinase-2 | HK2 | Cardiometabolic |
| P34947 | G protein-coupled receptor kinase 5 | GRK5 | Cardiometabolic |
| Q96N03 | V-set and transmembrane domain-containing protein 2-like protein | VSTM2L | Cardiometabolic |
| P31483 | Nucleolysin TIA-1 isoform p40 | TIA1 | Cardiometabolic |
| Q9Y2B0 | Protein canopy homolog 2 | CNPY2 | Cardiometabolic |
| O43186 | Cone-rod homeobox protein | CRX | Cardiometabolic |
| O95183 | Vesicle-associated membrane protein 5 | VAMP5 | Cardiometabolic |
| Q6UWL2 | Sushi domain-containing protein 1 | SUSD1 | Cardiometabolic |
| Q9UKL0 | REST corepressor 1 | RCOR1 | Cardiometabolic |
| P01375 | Tumor necrosis factor | TNF | Cardiometabolic |
| P35218 | Carbonic anhydrase 5A, mitochondrial | CA5A | Cardiometabolic |
| P51161 | Gastrotropin | FABP6 | Cardiometabolic |
| Q15831 | Serine/threonine-protein kinase STK11 | STK11 | Cardiometabolic |
| O60635 | Tetraspanin-1 | TSPAN1 | Cardiometabolic |
| P62736 | Actin, aortic smooth muscle | ACTA2 | Cardiometabolic |
| P58546 | Myotrophin | MTPN | Cardiometabolic |
| O43854 | EGF-like repeat and discoidin I-like domain-containing protein 3 | EDIL3 | Cardiometabolic |
| Q13105 | Zinc finger and BTB domain-containing protein 17 | ZBTB17 | Cardiometabolic |
| P21246 | Pleiotrophin | PTN | Cardiometabolic |
| O95684 | Centrosomal protein 43 | CEP43 | Cardiometabolic |
| Q12912 | Inositol 1,4,5-triphosphate receptor associated 2 | IRAG2 | Cardiometabolic |
| P21964 | Catechol O-methyltransferase | COMT | Cardiometabolic |
| P09237 | Matrilysin | MMP7 | Cardiometabolic |
| Q15165 | Serum paraoxonase/arylesterase 2 | PON2 | Cardiometabolic |
| Q8TE57 | A disintegrin and metalloproteinase with thrombospondin motifs 16 | ADAMTS16 | Cardiometabolic |
| P55259 | Pancreatic secretory granule membrane major glycoprotein GP2 | GP2 | Cardiometabolic |
| Q92558 | Wiskott-Aldrich syndrome protein family member 1 | WASF1 | Cardiometabolic |
| Q99549 | M-phase phosphoprotein 8 | MPHOSPH8 | Cardiometabolic |
| Q8NHS0 | DnaJ homolog subfamily B member 8 | DNAJB8 | Cardiometabolic |
| Q9H5Y7 | SLIT and NTRK-like protein 6 | SLITRK6 | Cardiometabolic |
| O15354 | Prosaposin receptor GPR37 | GPR37 | Cardiometabolic |
| P20718 | Granzyme H | GZMH | Cardiometabolic |
| P13807 | Glycogen [starch] synthase, muscle | GYS1 | Cardiometabolic |
| P40225 | Thrombopoietin | THPO | Cardiometabolic |
| Q8NC01 | C-type lectin domain family 1 member A | CLEC1A | Cardiometabolic |
| O75354 | Ectonucleoside triphosphate diphosphohydrolase 6 | ENTPD6 | Cardiometabolic |
| P05231 | Interleukin-6 | IL6 | Cardiometabolic |
| P31997 | Carcinoembryonic antigen-related cell adhesion molecule 8 | CEACAM8 | Cardiometabolic |
| P25815 | Protein S100-P | S100P | Cardiometabolic |
| O75356 | Ectonucleoside triphosphate diphosphohydrolase 5 | ENTPD5 | Cardiometabolic |
| Q9BYF1 | Angiotensin-converting enzyme 2 | ACE2 | Cardiometabolic |
| P07585 | Decorin | DCN | Cardiometabolic |
| Q04760 | Lactoylglutathione lyase | GLO1 | Cardiometabolic |
| Q9NWQ8 | Phosphoprotein associated with glycosphingolipid-enriched microdomains 1 | PAG1 | Cardiometabolic |
| Q13444 | Disintegrin and metalloproteinase domain-containing protein 15 | ADAM15 | Cardiometabolic |
| P40818 | Ubiquitin carboxyl-terminal hydrolase 8 | USP8 | Cardiometabolic |
| P22004 | Bone morphogenetic protein 6 | BMP6 | Cardiometabolic |
| Q9UKP3 | Integrin beta-1-binding protein 2 | ITGB1BP2 | Cardiometabolic |
| P09668 | Pro-cathepsin H | CTSH | Cardiometabolic |
| P46379 | Large proline-rich protein BAG6 | BAG6 | Cardiometabolic |
| O14793 | Growth/differentiation factor 8 | MSTN | Cardiometabolic |
| Q9BWV1 | Brother of CDO | BOC | Cardiometabolic |
| P08319 | All-trans-retinol dehydrogenase [NAD(+)] ADH4 | ADH4 | Cardiometabolic |
| P09496 | Clathrin light chain A | CLTA | Cardiometabolic |
| P34913 | Bifunctional epoxide hydrolase 2 | EPHX2 | Cardiometabolic |
| Q96A56 | Tumor protein p53-inducible nuclear protein 1 | TP53INP1 | Cardiometabolic |
| Q9Y4X3 | C-C motif chemokine 27 | CCL27 | Cardiometabolic |
| P00568 | Adenylate kinase isoenzyme 1 | AK1 | Cardiometabolic |
| P09525 | Annexin A4 | ANXA4 | Cardiometabolic |
| Q15067 | Peroxisomal acyl-coenzyme A oxidase 1 | ACOX1 | Cardiometabolic |
| NTproBNP | N-terminal prohormone of brain natriuretic peptide | NTproBNP | Cardiometabolic |
| Q05315 | Galectin-10 | CLC | Cardiometabolic |
| Q6PJW8 | Consortin | CNST | Cardiometabolic |
| P48357 | Leptin receptor | LEPR | Cardiometabolic |
| P01222 | Thyrotropin subunit beta | TSHB | Cardiometabolic |
| P31949 | Protein S100-A11 | S100A11 | Cardiometabolic |
| Q9H7M9 | V-type immunoglobulin domain-containing suppressor of T-cell activation | VSIR | Cardiometabolic |
| O14917 | Protocadherin-17 | PCDH17 | Cardiometabolic |
| P16234 | Platelet-derived growth factor receptor alpha | PDGFRA | Cardiometabolic |
| Q12864 | Cadherin-17 | CDH17 | Cardiometabolic |
| Q9Y5X1 | Sorting nexin-9 | SNX9 | Cardiometabolic |
| Q16620 | BDNF/NT-3 growth factors receptor | NTRK2 | Cardiometabolic |
| Q03154 | Aminoacylase-1 | ACY1 | Cardiometabolic |
| O60496 | Docking protein 2 | DOK2 | Cardiometabolic |
| P21549 | Serine--pyruvate aminotransferase | AGXT | Cardiometabolic |
| Q13043 | Serine/threonine-protein kinase 4 | STK4 | Cardiometabolic |
| P21980 | Protein-glutamine gamma-glutamyltransferase 2 | TGM2 | Cardiometabolic |
| Q9UHL4 | Dipeptidyl peptidase 2 | DPP7 | Cardiometabolic |
| O60664 | Perilipin-3 | PLIN3 | Cardiometabolic |
| O94903 | Pyridoxal phosphate homeostasis protein | PLPBP | Cardiometabolic |
| Q9UMF0 | Intercellular adhesion molecule 5 | ICAM5 | Cardiometabolic |
| P10644 | cAMP-dependent protein kinase type I-alpha regulatory subunit | PRKAR1A | Cardiometabolic |
| P40189 | Interleukin-6 receptor subunit beta | IL6ST | Cardiometabolic |
| Q9H773 | dCTP pyrophosphatase 1 | DCTPP1 | Cardiometabolic |
| Q6GTS8 | N-fatty-acyl-amino acid synthase/hydrolase PM20D1 | PM20D1 | Cardiometabolic |
| Q53H82 | Endoribonuclease LACTB2 | LACTB2 | Cardiometabolic |
| Q13158 | FAS-associated death domain protein | FADD | Cardiometabolic |
| Q99674 | Cell growth regulator with EF hand domain protein 1 | CGREF1 | Cardiometabolic |
| P10145 | Interleukin-8 | CXCL8 | Cardiometabolic |
| P09382 | Galectin-1 | LGALS1 | Cardiometabolic |
| Q8WX77 | Insulin-like growth factor-binding protein-like 1 | IGFBPL1 | Cardiometabolic |
| P41218 | Myeloid cell nuclear differentiation antigen | MNDA | Cardiometabolic |
| P55285 | Cadherin-6 | CDH6 | Cardiometabolic |
| Q9UHD0 | Interleukin-19 | IL19 | Cardiometabolic |
| P16112 | Aggrecan core protein | ACAN | Cardiometabolic |
| Q9ULL4 | Plexin-B3 | PLXNB3 | Cardiometabolic |
| Q06418 | Tyrosine-protein kinase receptor TYRO3 | TYRO3 | Cardiometabolic |
| Q13541 | Eukaryotic translation initiation factor 4E-binding protein 1 | EIF4EBP1 | Cardiometabolic |
| O75340 | Programmed cell death protein 6 | PDCD6 | Cardiometabolic |
| P51693 | Amyloid-like protein 1 | APLP1 | Cardiometabolic |
| Q9NY25 | C-type lectin domain family 5 member A | CLEC5A | Cardiometabolic |
| P08263 | Glutathione S-transferase A1 | GSTA1 | Cardiometabolic |
| Q969P0 | Immunoglobulin superfamily member 8 | IGSF8 | Cardiometabolic |
| Q16820 | Meprin A subunit beta | MEP1B | Cardiometabolic |
| P18827 | Syndecan-1 | SDC1 | Cardiometabolic |
| P19022 | Cadherin-2 | CDH2 | Cardiometabolic |
| Q8NI22 | Multiple coagulation factor deficiency protein 2 | MCFD2 | Cardiometabolic |
| Q96LA6 | Fc receptor-like protein 1 | FCRL1 | Cardiometabolic |
| Q14956 | Transmembrane glycoprotein NMB | GPNMB | Cardiometabolic |
| P09417 | Dihydropteridine reductase | QDPR | Cardiometabolic |
| Q9UBU3 | Appetite-regulating hormone | GHRL | Cardiometabolic |
| P41236 | Protein phosphatase inhibitor 2 | PPP1R2 | Cardiometabolic |
| Q9Y5K6 | CD2-associated protein | CD2AP | Cardiometabolic |
| O95544 | NAD kinase | NADK | Cardiometabolic |
| P27352 | Cobalamin binding intrinsic factor | CBLIF | Cardiometabolic |
| Q9GZM7 | Tubulointerstitial nephritis antigen-like | TINAGL1 | Cardiometabolic |
| Q8WVQ1 | Soluble calcium-activated nucleotidase 1 | CANT1 | Cardiometabolic |
| Q8WTU2 | Scavenger receptor cysteine-rich domain-containing group B protein | SSC4D | Cardiometabolic |
| P78380 | Oxidized low-density lipoprotein receptor 1 | OLR1 | Cardiometabolic |
| Q9NR28 | Diablo homolog, mitochondrial | DIABLO | Cardiometabolic |
| Q86VZ4 | Low-density lipoprotein receptor-related protein 11 | LRP11 | Cardiometabolic |
| Q13275 | Semaphorin-3F | SEMA3F | Cardiometabolic |
| P41159 | Leptin | LEP | Cardiometabolic |
| P06858 | Lipoprotein lipase | LPL | Cardiometabolic |
| Q8IZP9 | Adhesion G-protein coupled receptor G2 | ADGRG2 | Cardiometabolic |
| Q9Y286 | Sialic acid-binding Ig-like lectin 7 | SIGLEC7 | Cardiometabolic |
| O95502 | Neuronal pentraxin receptor | NPTXR | Cardiometabolic |
| O75791 | GRB2-related adapter protein 2 | GRAP2 | Cardiometabolic |
| Q9HBB8 | Cadherin-related family member 5 | CDHR5 | Cardiometabolic |
| P52888 | Thimet oligopeptidase | THOP1 | Cardiometabolic |
| P35754 | Glutaredoxin-1 | GLRX | Cardiometabolic |
| P21583 | Kit ligand | KITLG | Cardiometabolic |
| Q9UKJ0 | Paired immunoglobulin-like type 2 receptor beta | PILRB | Cardiometabolic |
| Q15846 | Clusterin-like protein 1 | CLUL1 | Cardiometabolic |
| P23526 | Adenosylhomocysteinase | AHCY | Cardiometabolic |
| P12104 | Fatty acid-binding protein, intestinal | FABP2 | Cardiometabolic |
| P00797 | Renin | REN | Cardiometabolic |
| Q9UK05 | Growth/differentiation factor 2 | GDF2 | Cardiometabolic |
| P12724 | Eosinophil cationic protein | RNASE3 | Cardiometabolic |
| Q9BQB4 | Sclerostin | SOST | Cardiometabolic |
| Q9NQX5 | Neural proliferation differentiation and control protein 1 | NPDC1 | Cardiometabolic |
| Q07108 | Early activation antigen CD69 | CD69 | Cardiometabolic |
| P42830 | C-X-C motif chemokine 5 | CXCL5 | Cardiometabolic |
| P02462 | Collagen alpha-1(IV) chain | COL4A1 | Cardiometabolic |
| A6NI73 | Leukocyte immunoglobulin-like receptor subfamily A member 5 | LILRA5 | Cardiometabolic |
| Q9UEW3 | Macrophage receptor MARCO | MARCO | Cardiometabolic |
| O95841 | Angiopoietin-related protein 1 | ANGPTL1 | Cardiometabolic |
| Q9BQR3 | Serine protease 27 | PRSS27 | Cardiometabolic |
| Q99523 | Sortilin | SORT1 | Cardiometabolic |
| P35247 | Pulmonary surfactant-associated protein D | SFTPD | Cardiometabolic |
| P20711 | Aromatic-L-amino-acid decarboxylase | DDC | Cardiometabolic |
| P31431 | Syndecan-4 | SDC4 | Cardiometabolic |
| P09601 | Heme oxygenase 1 | HMOX1 | Cardiometabolic |
| O00161 | Synaptosomal-associated protein 23 | SNAP23 | Cardiometabolic |
| Q8IW75 | Serpin A12 | SERPINA12 | Cardiometabolic |
| P01241 | Somatotropin | GH1 | Cardiometabolic |
| Q9BUD6 | Spondin-2 | SPON2 | Cardiometabolic |
| Q92692 | Nectin-2 | NECTIN2 | Cardiometabolic |
| Q6WN34 | Chordin-like protein 2 | CHRDL2 | Cardiometabolic |
| Q01973 | Inactive tyrosine-protein kinase transmembrane receptor ROR1 | ROR1 | Cardiometabolic |
| Q8N1Q1 | Carbonic anhydrase 13 | CA13 | Cardiometabolic |
| Q76M96 | Coiled-coil domain-containing protein 80 | CCDC80 | Cardiometabolic |
| P09467 | Fructose-1,6-bisphosphatase 1 | FBP1 | Cardiometabolic |
| P07711 | Cathepsin L1 | CTSL | Cardiometabolic |
| Q92823 | Neuronal cell adhesion molecule | NRCAM | Cardiometabolic |
| P04792 | Heat shock protein beta-1 | HSPB1 | Cardiometabolic |
| P07204 | Thrombomodulin | THBD | Cardiometabolic |
| Q16773 | Kynurenine--oxoglutarate transaminase 1 | KYAT1 | Cardiometabolic |
| Q92520 | Protein FAM3C | FAM3C | Cardiometabolic |
| P19971 | Thymidine phosphorylase | TYMP | Cardiometabolic |
| Q8NBP7 | Proprotein convertase subtilisin/kexin type 9 | PCSK9 | Cardiometabolic |
| Q16270 | Insulin-like growth factor-binding protein 7 | IGFBP7 | Cardiometabolic |
| P07911 | Uromodulin | UMOD | Cardiometabolic |
| P55058 | Phospholipid transfer protein | PLTP | Cardiometabolic |
| Q13361 | Microfibrillar-associated protein 5 | MFAP5 | Cardiometabolic |
| P01130 | Low-density lipoprotein receptor | LDLR | Cardiometabolic |
| P22748 | Carbonic anhydrase 4 | CA4 | Cardiometabolic |
| P54760 | Ephrin type-B receptor 4 | EPHB4 | Cardiometabolic |
| P33151 | Cadherin-5 | CDH5 | Cardiometabolic |
| P23141 | Liver carboxylesterase 1 | CES1 | Cardiometabolic |
| P15090 | Fatty acid-binding protein, adipocyte | FABP4 | Cardiometabolic |
| P08833 | Insulin-like growth factor-binding protein 1 | IGFBP1 | Cardiometabolic |
| P00750 | Tissue-type plasminogen activator | PLAT | Cardiometabolic |
| P13598 | Intercellular adhesion molecule 2 | ICAM2 | Cardiometabolic |
| Q76LX8 | A disintegrin and metalloproteinase with thrombospondin motifs 13 | ADAMTS13 | Cardiometabolic |
| Q01638 | Interleukin-1 receptor-like 1 | IL1RL1 | Cardiometabolic |
| Q99988 | Growth/differentiation factor 15 | GDF15 | Cardiometabolic |
| P04054 | Phospholipase A2 | PLA2G1B | Cardiometabolic |
| Q9UM47 | Neurogenic locus notch homolog protein 3 | NOTCH3 | Cardiometabolic |
| Q14767 | Latent-transforming growth factor beta-binding protein 2 | LTBP2 | Cardiometabolic |
| Q03167 | Transforming growth factor beta receptor type 3 | TGFBR3 | Cardiometabolic |
| P04275 | von Willebrand factor | VWF | Cardiometabolic |
| Q5VY43 | Platelet endothelial aggregation receptor 1 | PEAR1 | Cardiometabolic |
| Q06141 | Regenerating islet-derived protein 3-alpha | REG3A | Cardiometabolic |
| P55808 | Glycoprotein Xg | XG | Cardiometabolic |
| P48960 | Adhesion G protein-coupled receptor E5 | ADGRE5 | Cardiometabolic |
| P04066 | Tissue alpha-L-fucosidase | FUCA1 | Cardiometabolic |
| Q9NNX6 | CD209 antigen | CD209 | Cardiometabolic |
| P02144 | Myoglobin | MB | Cardiometabolic |
| Q9UBP4 | Dickkopf-related protein 3 | DKK3 | Cardiometabolic |
| O14798 | Tumor necrosis factor receptor superfamily member 10C | TNFRSF10C | Cardiometabolic |
| P48304 | Lithostathine-1-beta | REG1B | Cardiometabolic |
| P01589 | Interleukin-2 receptor subunit alpha | IL2RA | Cardiometabolic |
| P09619 | Platelet-derived growth factor receptor beta | PDGFRB | Cardiometabolic |
| P08581 | Hepatocyte growth factor receptor | MET | Cardiometabolic |
| Q96AP7 | Endothelial cell-selective adhesion molecule | ESAM | Cardiometabolic |
| P07451 | Carbonic anhydrase 3 | CA3 | Cardiometabolic |
| Q9Y275 | Tumor necrosis factor ligand superfamily member 13B | TNFSF13B | Cardiometabolic |
| Q13740 | CD166 antigen | ALCAM | Cardiometabolic |
| P14555 | Phospholipase A2, membrane associated | PLA2G2A | Cardiometabolic |
| P08118 | Beta-microseminoprotein | MSMB | Cardiometabolic |
| P07858 | Cathepsin B | CTSB | Cardiometabolic |
| P32942 | Intercellular adhesion molecule 3 | ICAM3 | Cardiometabolic |
| Q07507 | Dermatopontin | DPT | Cardiometabolic |
| Q9NQ79 | Cartilage acidic protein 1 | CRTAC1 | Cardiometabolic |
| P15085 | Carboxypeptidase A1 | CPA1 | Cardiometabolic |
| Q86U17 | Serpin A11 | SERPINA11 | Cardiometabolic |
| Q9H2A7 | C-X-C motif chemokine 16 | CXCL16 | Cardiometabolic |
| O75326 | Semaphorin-7A | SEMA7A | Cardiometabolic |
| P17931 | Galectin-3 | LGALS3 | Cardiometabolic |
| Q9H1U4 | Multiple epidermal growth factor-like domains protein 9 | MEGF9 | Cardiometabolic |
| P31146 | Coronin-1A | CORO1A | Cardiometabolic |
| P17813 | Endoglin | ENG | Cardiometabolic |
| Q13822 | Ectonucleotide pyrophosphatase/phosphodiesterase family member 2 | ENPP2 | Cardiometabolic |
| P15086 | Carboxypeptidase B | CPB1 | Cardiometabolic |
| P16581 | E-selectin | SELE | Cardiometabolic |
| P19021 | Peptidyl-glycine alpha-amidating monooxygenase | PAM | Cardiometabolic |
| P12111 | Collagen alpha-3(VI) chain | COL6A3 | Cardiometabolic |
| Q15828 | Cystatin-M | CST6 | Cardiometabolic |
| P04080 | Cystatin-B | CSTB | Cardiometabolic |
| P80370 | Protein delta homolog 1 | DLK1 | Cardiometabolic |
| P08236 | Beta-glucuronidase | GUSB | Cardiometabolic |
| Q14162 | Scavenger receptor class F member 1 | SCARF1 | Cardiometabolic |
| P09093 | Chymotrypsin-like elastase family member 3A | CELA3A | Cardiometabolic |
| P48745 | CCN family member 3 | CCN3 | Cardiometabolic |
| Q9Y4L1 | Hypoxia up-regulated protein 1 | HYOU1 | Cardiometabolic |
| Q8N423 | Leukocyte immunoglobulin-like receptor subfamily B member 2 | LILRB2 | Cardiometabolic |
| P15907 | Beta-galactoside alpha-2,6-sialyltransferase 1 | ST6GAL1 | Cardiometabolic |
| P35590 | Tyrosine-protein kinase receptor Tie-1 | TIE1 | Cardiometabolic |
| P78324 | Tyrosine-protein phosphatase non-receptor type substrate 1 | SIRPA | Cardiometabolic |
| P42574 | Caspase-3 | CASP3 | Cardiometabolic |
| Q13332 | Receptor-type tyrosine-protein phosphatase S | PTPRS | Cardiometabolic |
| Q12860 | Contactin-1 | CNTN1 | Cardiometabolic |
| Q8TDL5 | BPI fold-containing family B member 1 | BPIFB1 | Cardiometabolic |
| P20160 | Azurocidin | AZU1 | Cardiometabolic |
| P10586 | Receptor-type tyrosine-protein phosphatase F | PTPRF | Cardiometabolic |
| P46531 | Neurogenic locus notch homolog protein 1 | NOTCH1 | Cardiometabolic |
| Q13231 | Chitotriosidase-1 | CHIT1 | Cardiometabolic |
| P04085 | Platelet-derived growth factor subunit A | PDGFA | Cardiometabolic |
| P13686 | Tartrate-resistant acid phosphatase type 5 | ACP5 | Cardiometabolic |
| P05107 | Integrin beta-2 | ITGB2 | Cardiometabolic |
| P25445 | Tumor necrosis factor receptor superfamily member 6 | FAS | Cardiometabolic |
| O15031 | Plexin-B2 | PLXNB2 | Cardiometabolic |
| Q14393 | Growth arrest-specific protein 6 | GAS6 | Cardiometabolic |
| P00533 | Epidermal growth factor receptor | EGFR | Cardiometabolic |
| Q16769 | Glutaminyl-peptide cyclotransferase | QPCT | Cardiometabolic |
| O00584 | Ribonuclease T2 | RNASET2 | Cardiometabolic |
| P10451 | Osteopontin | SPP1 | Cardiometabolic |
| Q8NHL6 | Leukocyte immunoglobulin-like receptor subfamily B member 1 | LILRB1 | Cardiometabolic |
| O75023 | Leukocyte immunoglobulin-like receptor subfamily B member 5 | LILRB5 | Cardiometabolic |
| P18065 | Insulin-like growth factor-binding protein 2 | IGFBP2 | Cardiometabolic |
| P19957 | Elafin | PI3 | Cardiometabolic |
| Q9HD89 | Resistin | RETN | Cardiometabolic |
| Q16663 | C-C motif chemokine 15 | CCL15 | Cardiometabolic |
| P24158 | Myeloblastin | PRTN3 | Cardiometabolic |
| Q12884 | Prolyl endopeptidase FAP | FAP | Cardiometabolic |
| P23284 | Peptidyl-prolyl cis-trans isomerase B | PPIB | Cardiometabolic |
| P39060 | Collagen alpha-1(XVIII) chain | COL18A1 | Cardiometabolic |
| P04746 | Pancreatic alpha-amylase | AMY2A | Cardiometabolic |
| O15467 | C-C motif chemokine 16 | CCL16 | Cardiometabolic |
| P02452 | Collagen alpha-1(I) chain | COL1A1 | Cardiometabolic |
| Q13867 | Bleomycin hydrolase | BLMH | Cardiometabolic |
| P42785 | Lysosomal Pro-X carboxypeptidase | PRCP | Cardiometabolic |
| O75594 | Peptidoglycan recognition protein 1 | PGLYRP1 | Cardiometabolic |
| P13987 | CD59 glycoprotein | CD59 | Cardiometabolic |
| P19961 | Alpha-amylase 2B | AMY2B | Cardiometabolic |
| P20062 | Transcobalamin-2 | TCN2 | Cardiometabolic |
| P05121 | Plasminogen activator inhibitor 1 | SERPINE1 | Cardiometabolic |
| P43121 | Cell surface glycoprotein MUC18 | MCAM | Cardiometabolic |
| P59665 | Neutrophil defensin 1 | DEFA1_DEFA1B | Cardiometabolic |
| Q6EMK4 | Vasorin | VASN | Cardiometabolic |
| Q13508 | Ecto-ADP-ribosyltransferase 3 | ART3 | Cardiometabolic |
| Q96KN2 | Beta-Ala-His dipeptidase | CNDP1 | Cardiometabolic |
| O95998 | Interleukin-18-binding protein | IL18BP | Cardiometabolic |
| P00740 | Coagulation factor IX | F9 | Cardiometabolic |
| P15144 | Aminopeptidase N | ANPEP | Cardiometabolic |
| A1L4H1 | Soluble scavenger receptor cysteine-rich domain-containing protein SSC5D | SSC5D | Cardiometabolic |
| Q06033 | Inter-alpha-trypsin inhibitor heavy chain H3 | ITIH3 | Cardiometabolic |
| P12830 | Cadherin-1 | CDH1 | Cardiometabolic |
| P13591 | Neural cell adhesion molecule 1 | NCAM1 | Cardiometabolic |
| P18428 | Lipopolysaccharide-binding protein | LBP | Cardiometabolic |
| Q99650 | Oncostatin-M-specific receptor subunit beta | OSMR | Cardiometabolic |
| Q12794 | Hyaluronidase-1 | HYAL1 | Cardiometabolic |
| P07339 | Cathepsin D | CTSD | Cardiometabolic |
| P24821 | Tenascin | TNC | Cardiometabolic |
| Q86VB7 | Scavenger receptor cysteine-rich type 1 protein M130 | CD163 | Cardiometabolic |
| Q14515 | SPARC-like protein 1 | SPARCL1 | Cardiometabolic |
| P14543 | Nidogen-1 | NID1 | Cardiometabolic |
| P30530 | Tyrosine-protein kinase receptor UFO | AXL | Cardiometabolic |
| P07478 | Trypsin-2 | PRSS2 | Cardiometabolic |
| Q9UBR2 | Cathepsin Z | CTSZ | Cardiometabolic |
| O00533 | Neural cell adhesion molecule L1-like protein | CHL1 | Cardiometabolic |
| Q9BXJ1 | Complement C1q tumor necrosis factor-related protein 1 | C1QTNF1 | Cardiometabolic |
| P35443 | Thrombospondin-4 | THBS4 | Cardiometabolic |
| P02786 | Transferrin receptor protein 1 | TFRC | Cardiometabolic |
| P10721 | Mast/stem cell growth factor receptor Kit | KIT | Cardiometabolic |
| Q07654 | Trefoil factor 3 | TFF3 | Cardiometabolic |
| P08709 | Coagulation factor VII | F7 | Cardiometabolic |
| Q99969 | Retinoic acid receptor responder protein 2 | RARRES2 | Cardiometabolic |
| O95445 | Apolipoprotein M | APOM | Cardiometabolic |
| Q96H15 | T-cell immunoglobulin and mucin domain-containing protein 4 | TIMD4 | Cardiometabolic |
| P05556 | Integrin beta-1 | ITGB1 | Cardiometabolic |
| P08174 | Complement decay-accelerating factor | CD55 | Cardiometabolic |
| P08571 | Monocyte differentiation antigen CD14 | CD14 | Cardiometabolic |
| P16109 | P-selectin | SELP | Cardiometabolic |
| P15529 | Membrane cofactor protein | CD46 | Cardiometabolic |
| P07359 | Platelet glycoprotein Ib alpha chain | GP1BA | Cardiometabolic |
| Q15485 | Ficolin-2 | FCN2 | Cardiometabolic |
| P98160 | Basement membrane-specific heparan sulfate proteoglycan core protein | HSPG2 | Cardiometabolic |
| Q15113 | Procollagen C-endopeptidase enhancer 1 | PCOLCE | Cardiometabolic |
| P08887 | Interleukin-6 receptor subunit alpha | IL6R | Cardiometabolic |
| P00441 | Superoxide dismutase [Cu-Zn] | SOD1 | Cardiometabolic |
| O75015 | Low affinity immunoglobulin gamma Fc region receptor III-B | FCGR3B | Cardiometabolic |
| P10646 | Tissue factor pathway inhibitor | TFPI | Cardiometabolic |
| Q16853 | Membrane primary amine oxidase | AOC3 | Cardiometabolic |
| O14786 | Neuropilin-1 | NRP1 | Cardiometabolic |
| P12318 | Low affinity immunoglobulin gamma Fc region receptor II-a | FCGR2A | Cardiometabolic |
| P80188 | Neutrophil gelatinase-associated lipocalin | LCN2 | Cardiometabolic |
| P20023 | Complement receptor type 2 | CR2 | Cardiometabolic |
| Q9NZK5 | Adenosine deaminase 2 | ADA2 | Cardiometabolic |
| P55774 | C-C motif chemokine 18 | CCL18 | Cardiometabolic |
| P19320 | Vascular cell adhesion protein 1 | VCAM1 | Cardiometabolic |
| Q9NPY3 | Complement component C1q receptor | CD93 | Cardiometabolic |
| P17936 | Insulin-like growth factor-binding protein 3 | IGFBP3 | Cardiometabolic |
| P36222 | Chitinase-3-like protein 1 | CHI3L1 | Cardiometabolic |
| P01034 | Cystatin-C | CST3 | Cardiometabolic |
| Q16627 | C-C motif chemokine 14 | CCL14 | Cardiometabolic |
| P04070 | Vitamin K-dependent protein C | PROC | Cardiometabolic |
| P03950 | Angiogenin | ANG | Cardiometabolic |
| Q9UGM5 | Fetuin-B | FETUB | Cardiometabolic |
| P49747 | Cartilage oligomeric matrix protein | COMP | Cardiometabolic |
| P27487 | Dipeptidyl peptidase 4 | DPP4 | Cardiometabolic |
| Q9Y5C1 | Angiopoietin-related protein 3 | ANGPTL3 | Cardiometabolic |
| P41222 | Prostaglandin-H2 D-isomerase | PTGDS | Cardiometabolic |
| P00915 | Carbonic anhydrase 1 | CA1 | Cardiometabolic |
| P06681 | Complement C2 | C2 | Cardiometabolic |
| P05362 | Intercellular adhesion molecule 1 | ICAM1 | Cardiometabolic |
| P13501 | C-C motif chemokine 5 | CCL5 | Cardiometabolic |
| P24592 | Insulin-like growth factor-binding protein 6 | IGFBP6 | Cardiometabolic |
| Q12805 | EGF-containing fibulin-like extracellular matrix protein 1 | EFEMP1 | Cardiometabolic |
| P05451 | Lithostathine-1-alpha | REG1A | Cardiometabolic |
| Q92820 | Gamma-glutamyl hydrolase | GGH | Cardiometabolic |
| Q15582 | Transforming growth factor-beta-induced protein ig-h3 | TGFBI | Cardiometabolic |
| P01033 | Metalloproteinase inhibitor 1 | TIMP1 | Cardiometabolic |
| Q8IZC4 | Rhotekin-2 | RTKN2 | Cardiometabolic_II |
| P78524 | DENN domain-containing protein 2B | DENND2B | Cardiometabolic_II |
| Q9H2M3 | S-methylmethionine--homocysteine S-methyltransferase BHMT2 | BHMT2 | Cardiometabolic_II |
| P55769 | NHP2-like protein 1 | SNU13 | Cardiometabolic_II |
| Q9Y2W1 | Thyroid hormone receptor-associated protein 3 | THRAP3 | Cardiometabolic_II |
| O43734 | E3 ubiquitin ligase TRAF3IP2 | TRAF3IP2 | Cardiometabolic_II |
| O00567 | Nucleolar protein 56 | NOP56 | Cardiometabolic_II |
| Q15477 | Helicase SKI2W | SKIV2L | Cardiometabolic_II |
| P25391 | Laminin subunit alpha-1 | LAMA1 | Cardiometabolic_II |
| P06753 | Tropomyosin alpha-3 chain | TPM3 | Cardiometabolic_II |
| P48507 | Glutamate--cysteine ligase regulatory subunit | GCLM | Cardiometabolic_II |
| Q9NZJ5 | Eukaryotic translation initiation factor 2-alpha kinase 3 | EIF2AK3 | Cardiometabolic_II |
| Q9Y623 | Myosin-4 | MYH4 | Cardiometabolic_II |
| P23634 | Plasma membrane calcium-transporting ATPase 4 | ATP2B4 | Cardiometabolic_II |
| O14958 | Calsequestrin-2 | CASQ2 | Cardiometabolic_II |
| O95180 | Voltage-dependent T-type calcium channel subunit alpha-1H | CACNA1H | Cardiometabolic_II |
| P54709 | Sodium/potassium-transporting ATPase subunit beta-3 | ATP1B3 | Cardiometabolic_II |
| Q13503 | Mediator of RNA polymerase II transcription subunit 21 | MED21 | Cardiometabolic_II |
| P08913 | Alpha-2A adrenergic receptor | ADRA2A | Cardiometabolic_II |
| P49755 | Transmembrane emp24 domain-containing protein 10 | TMED10 | Cardiometabolic_II |
| Q96DA2 | Ras-related protein Rab-39B | RAB39B | Cardiometabolic_II |
| P46783 | 40S ribosomal protein S10 | RPS10 | Cardiometabolic_II |
| O00291 | Huntingtin-interacting protein 1 | HIP1 | Cardiometabolic_II |
| P04141 | Granulocyte-macrophage colony-stimulating factor | CSF2 | Cardiometabolic_II |
| A6NCE7 | Microtubule-associated proteins 1A/1B light chain 3 beta 2 | MAP1LC3B2 | Cardiometabolic_II |
| Q9Y3B8 | Oligoribonuclease, mitochondrial | REXO2 | Cardiometabolic_II |
| Q6UWF7 | NXPE family member 4 | NXPE4 | Cardiometabolic_II |
| P55011 | Solute carrier family 12 member 2 | SLC12A2 | Cardiometabolic_II |
| P10109 | Adrenodoxin, mitochondrial | FDX1 | Cardiometabolic_II |
| P30049 | ATP synthase subunit delta, mitochondrial | ATP5F1D | Cardiometabolic_II |
| P33121 | Long-chain-fatty-acid--CoA ligase 1 | ACSL1 | Cardiometabolic_II |
| O60701 | UDP-glucose 6-dehydrogenase | UGDH | Cardiometabolic_II |
| Q9BY32 | Inosine triphosphate pyrophosphatase | ITPA | Cardiometabolic_II |
| Q01780 | Exosome component 10 | EXOSC10 | Cardiometabolic_II |
| Q08499 | cAMP-specific 3',5'-cyclic phosphodiesterase 4D | PDE4D | Cardiometabolic_II |
| P21817 | Ryanodine receptor 1 | RYR1 | Cardiometabolic_II |
| Q96HD9 | N-acyl-aromatic-L-amino acid amidohydrolase | ACY3 | Cardiometabolic_II |
| P35228 | Nitric oxide synthase, inducible | NOS2 | Cardiometabolic_II |
| E2RYF7 | Protein PBMUCL2 | HCG22 | Cardiometabolic_II |
| Q9NVZ3 | Adaptin ear-binding coat-associated protein 2 | NECAP2 | Cardiometabolic_II |
| Q9Y4C8 | Probable RNA-binding protein 19 | RBM19 | Cardiometabolic_II |
| Q07973 | 1,25-dihydroxyvitamin D(3) 24-hydroxylase, mitochondrial | CYP24A1 | Cardiometabolic_II |
| Q04695 | Keratin, type I cytoskeletal 17 | KRT17 | Cardiometabolic_II |
| Q15059 | Bromodomain-containing protein 3 | BRD3 | Cardiometabolic_II |
| B6SEH8 | Endogenous retrovirus group V member 1 Env polyprotein | ERVV-1 | Cardiometabolic_II |
| O95858 | Tetraspanin-15 | TSPAN15 | Cardiometabolic_II |
| Q9H347 | Ubiquilin-3 | UBQLN3 | Cardiometabolic_II |
| P06729 | T-cell surface antigen CD2 | CD2 | Cardiometabolic_II |
| Q96IW2 | SH2 domain-containing adapter protein D | SHD | Cardiometabolic_II |
| A6BM72 | Multiple epidermal growth factor-like domains protein 11 | MEGF11 | Cardiometabolic_II |
| Q9UKX7 | Nuclear pore complex protein Nup50 | NUP50 | Cardiometabolic_II |
| Q96LB8 | Peptidoglycan recognition protein 4 | PGLYRP4 | Cardiometabolic_II |
| Q9NV35 | Nucleotide triphosphate diphosphatase NUDT15 | NUDT15 | Cardiometabolic_II |
| Q10587 | Thyrotroph embryonic factor | TEF | Cardiometabolic_II |
| Q13296 | Mammaglobin-A | SCGB2A2 | Cardiometabolic_II |
| P20929 | Nebulin | NEB | Cardiometabolic_II |
| Q5TA50 | Ceramide-1-phosphate transfer protein | CPTP | Cardiometabolic_II |
| Q86UW2 | Organic solute transporter subunit beta | SLC51B | Cardiometabolic_II |
| Q8WZ42 | Titin | TTN | Cardiometabolic_II |
| Q9UFP1 | Golgi-associated kinase 1A | GASK1A | Cardiometabolic_II |
| Q99707 | Methionine synthase | MTR | Cardiometabolic_II |
| P21673 | Diamine acetyltransferase 1 | SAT1 | Cardiometabolic_II |
| O00425 | Insulin-like growth factor 2 mRNA-binding protein 3 | IGF2BP3 | Cardiometabolic_II |
| O43290 | U4/U6.U5 tri-snRNP-associated protein 1 | SART1 | Cardiometabolic_II |
| Q92935 | Exostosin-like 1 | EXTL1 | Cardiometabolic_II |
| Q8N8E3 | Centrosomal protein of 112 kDa | CEP112 | Cardiometabolic_II |
| P16066 | Atrial natriuretic peptide receptor 1 | NPR1 | Cardiometabolic_II |
| Q6NZY4 | Zinc finger CCHC domain-containing protein 8 | ZCCHC8 | Cardiometabolic_II |
| P14415 | Sodium/potassium-transporting ATPase subunit beta-2 | ATP1B2 | Cardiometabolic_II |
| Q96K76 | Ubiquitin carboxyl-terminal hydrolase 47 | USP47 | Cardiometabolic_II |
| P05976 | Myosin light chain 1/3, skeletal muscle isoform | MYL1 | Cardiometabolic_II |
| Q9Y2Y0 | ADP-ribosylation factor-like protein 2-binding protein | ARL2BP | Cardiometabolic_II |
| Q14088 | Ras-related protein Rab-33A | RAB33A | Cardiometabolic_II |
| P38935 | DNA-binding protein SMUBP-2 | IGHMBP2 | Cardiometabolic_II |
| P05026 | Sodium/potassium-transporting ATPase subunit beta-1 | ATP1B1 | Cardiometabolic_II |
| O15305 | Phosphomannomutase 2 | PMM2 | Cardiometabolic_II |
| Q9BW61 | DET1- and DDB1-associated protein 1 | DDA1 | Cardiometabolic_II |
| Q15370 | Elongin-B | ELOB | Cardiometabolic_II |
| Q8NET8 | Transient receptor potential cation channel subfamily V member 3 | TRPV3 | Cardiometabolic_II |
| P05000 | Interferon omega-1 | IFNW1 | Cardiometabolic_II |
| Q15018 | BRISC complex subunit Abraxas 2 | ABRAXAS2 | Cardiometabolic_II |
| P54296 | Myomesin-2 | MYOM2 | Cardiometabolic_II |
| Q16836 | Hydroxyacyl-coenzyme A dehydrogenase, mitochondrial | HADH | Cardiometabolic_II |
| Q14353 | Guanidinoacetate N-methyltransferase | GAMT | Cardiometabolic_II |
| P23511 | Nuclear transcription factor Y subunit alpha | NFYA | Cardiometabolic_II |
| Q07075 | Glutamyl aminopeptidase | ENPEP | Cardiometabolic_II |
| Q9BV94 | ER degradation-enhancing alpha-mannosidase-like protein 2 | EDEM2 | Cardiometabolic_II |
| Q02127 | Dihydroorotate dehydrogenase | DHODH | Cardiometabolic_II |
| P57078 | Receptor-interacting serine/threonine-protein kinase 4 | RIPK4 | Cardiometabolic_II |
| Q6ZN66 | Guanylate-binding protein 6 | GBP6 | Cardiometabolic_II |
| Q9BZL6 | Serine/threonine-protein kinase D2 | PRKD2 | Cardiometabolic_II |
| A6NHS7 | MANSC domain-containing protein 4 | MANSC4 | Cardiometabolic_II |
| O75521 | Enoyl-CoA delta isomerase 2 | ECI2 | Cardiometabolic_II |
| P12270 | Nucleoprotein TPR | TPR | Cardiometabolic_II |
| Q9NYX4 | Neuron-specific vesicular protein calcyon | CALY | Cardiometabolic_II |
| P37058 | Testosterone 17-beta-dehydrogenase 3 | HSD17B3 | Cardiometabolic_II |
| Q9BZC7 | ATP-binding cassette sub-family A member 2 | ABCA2 | Cardiometabolic_II |
| Q6P4F2 | Ferredoxin-2, mitochondrial | FDX2 | Cardiometabolic_II |
| Q16774 | Guanylate kinase | GUK1 | Cardiometabolic_II |
| Q9UNN8 | Endothelial protein C receptor | PROCR | Cardiometabolic_II |
| P10082 | Peptide YY | PYY | Cardiometabolic_II |
| O15018 | PDZ domain-containing protein 2 | PDZD2 | Cardiometabolic_II |
| Q16206 | Ecto-NOX disulfide-thiol exchanger 2 | ENOX2 | Cardiometabolic_II |
| P0C7L1 | Serine protease inhibitor Kazal-type 8 | SPINK8 | Cardiometabolic_II |
| Q7Z7H5 | Transmembrane emp24 domain-containing protein 4 | TMED4 | Cardiometabolic_II |
| Q9Y2L6 | FERM domain-containing protein 4B | FRMD4B | Cardiometabolic_II |
| P55010 | Eukaryotic translation initiation factor 5 | EIF5 | Cardiometabolic_II |
| Q01581 | Hydroxymethylglutaryl-CoA synthase, cytoplasmic | HMGCS1 | Cardiometabolic_II |
| Q12986 | Transcriptional repressor NF-X1 | NFX1 | Cardiometabolic_II |
| P22033 | Methylmalonyl-CoA mutase, mitochondrial | MMUT | Cardiometabolic_II |
| P19838 | Nuclear factor NF-kappa-B p105 subunit | NFKB1 | Cardiometabolic_II |
| Q01484 | Ankyrin-2 | ANK2 | Cardiometabolic_II |
| P59901 | Leukocyte immunoglobulin-like receptor subfamily A member 4 | LILRA4 | Cardiometabolic_II |
| O43896 | Kinesin-like protein KIF1C | KIF1C | Cardiometabolic_II |
| Q03013 | Glutathione S-transferase Mu 4 | GSTM4 | Cardiometabolic_II |
| O94766 | Galactosylgalactosylxylosylprotein 3-beta-glucuronosyltransferase 3 | B3GAT3 | Cardiometabolic_II |
| Q3SXY8 | ADP-ribosylation factor-like protein 13B | ARL13B | Cardiometabolic_II |
| O95670 | V-type proton ATPase subunit G 2 | ATP6V1G2 | Cardiometabolic_II |
| O00327 | Aryl hydrocarbon receptor nuclear translocator-like protein 1 | ARNTL | Cardiometabolic_II |
| P48668 | Keratin, type II cytoskeletal 6C | KRT6C | Cardiometabolic_II |
| P00966 | Argininosuccinate synthase | ASS1 | Cardiometabolic_II |
| Q96PU4 | E3 ubiquitin-protein ligase UHRF2 | UHRF2 | Cardiometabolic_II |
| P20382 | Pro-MCH | PMCH | Cardiometabolic_II |
| P35606 | Coatomer subunit beta' | COPB2 | Cardiometabolic_II |
| P13224 | Platelet glycoprotein Ib beta chain | GP1BB | Cardiometabolic_II |
| Q14807 | Kinesin-like protein KIF22 | KIF22 | Cardiometabolic_II |
| P50461 | Cysteine and glycine-rich protein 3 | CSRP3 | Cardiometabolic_II |
| Q14781 | Chromobox protein homolog 2 | CBX2 | Cardiometabolic_II |
| Q96A35 | 39S ribosomal protein L24, mitochondrial | MRPL24 | Cardiometabolic_II |
| Q58F21 | Bromodomain testis-specific protein | BRDT | Cardiometabolic_II |
| Q96EU7 | C1GALT1-specific chaperone 1 | C1GALT1C1 | Cardiometabolic_II |
| Q5VVQ6 | Ubiquitin thioesterase OTU1 | YOD1 | Cardiometabolic_II |
| A6NDB9 | Paralemmin-3 | PALM3 | Cardiometabolic_II |
| O75534 | Cold shock domain-containing protein E1 | CSDE1 | Cardiometabolic_II |
| Q13563 | Polycystin-2 | PKD2 | Cardiometabolic_II |
| Q99598 | Translin-associated protein X | TSNAX | Cardiometabolic_II |
| Q86VP3 | Phosphofurin acidic cluster sorting protein 2 | PACS2 | Cardiometabolic_II |
| Q5W0V3 | FHF complex subunit HOOK interacting protein 2A | FHIP2A | Cardiometabolic_II |
| P41227 | N-alpha-acetyltransferase 10 | NAA10 | Cardiometabolic_II |
| Q86VR7 | V-set and immunoglobulin domain-containing protein 10-like | VSIG10L | Cardiometabolic_II |
| Q93052 | Lipoma-preferred partner | LPP | Cardiometabolic_II |
| O75427 | Leucine-rich repeat and calponin homology domain-containing protein 4 | LRCH4 | Cardiometabolic_II |
| P35609 | Alpha-actinin-2 | ACTN2 | Cardiometabolic_II |
| P32241 | Vasoactive intestinal polypeptide receptor 1 | VIPR1 | Cardiometabolic_II |
| Q8ND90 | Paraneoplastic antigen Ma1 | PNMA1 | Cardiometabolic_II |
| Q5JTV8 | Torsin-1A-interacting protein 1 | TOR1AIP1 | Cardiometabolic_II |
| Q9UBV2 | Protein sel-1 homolog 1 | SEL1L | Cardiometabolic_II |
| P46926 | Glucosamine-6-phosphate isomerase 1 | GNPDA1 | Cardiometabolic_II |
| Q8NFP7 | Diphosphoinositol polyphosphate phosphohydrolase 3-alpha | NUDT10 | Cardiometabolic_II |
| Q14324 | Myosin-binding protein C, fast-type | MYBPC2 | Cardiometabolic_II |
| P35520 | Cystathionine beta-synthase | CBS | Cardiometabolic_II |
| O14841 | 5-oxoprolinase | OPLAH | Cardiometabolic_II |
| Q8WXC3 | Pyrin domain-containing protein 1 | PYDC1 | Cardiometabolic_II |
| O43423 | Acidic leucine-rich nuclear phosphoprotein 32 family member C | ANP32C | Cardiometabolic_II |
| Q9BQI0 | Allograft inflammatory factor 1-like | AIF1L | Cardiometabolic_II |
| Q8TER0 | Sushi, nidogen and EGF-like domain-containing protein 1 | SNED1 | Cardiometabolic_II |
| Q9BTK6 | PAXIP1-associated glutamate-rich protein 1 | PAGR1 | Cardiometabolic_II |
| Q9H173 | Nucleotide exchange factor SIL1 | SIL1 | Cardiometabolic_II |
| P20645 | Cation-dependent mannose-6-phosphate receptor | M6PR | Cardiometabolic_II |
| P13929 | Beta-enolase | ENO3 | Cardiometabolic_II |
| Q96ID5 | Immunoglobulin superfamily member 21 | IGSF21 | Cardiometabolic_II |
| P23327 | Sarcoplasmic reticulum histidine-rich calcium-binding protein | HRC | Cardiometabolic_II |
| P29536 | Leiomodin-1 | LMOD1 | Cardiometabolic_II |
| Q13316 | Dentin matrix acidic phosphoprotein 1 | DMP1 | Cardiometabolic_II |
| P35914 | Hydroxymethylglutaryl-CoA lyase, mitochondrial | HMGCL | Cardiometabolic_II |
| Q9Y5X3 | Sorting nexin-5 | SNX5 | Cardiometabolic_II |
| Q14643 | Inositol 1,4,5-trisphosphate receptor type 1 | ITPR1 | Cardiometabolic_II |
| Q99807 | 5-demethoxyubiquinone hydroxylase, mitochondrial | COQ7 | Cardiometabolic_II |
| Q99942 | E3 ubiquitin-protein ligase RNF5 | RNF5 | Cardiometabolic_II |
| P36776 | Lon protease homolog, mitochondrial | LONP1 | Cardiometabolic_II |
| Q14457 | Beclin-1 | BECN1 | Cardiometabolic_II |
| I3L3R5 | Coiled-coil domain-containing glutamate-rich protein 2 | CCER2 | Cardiometabolic_II |
| Q8N668 | COMM domain-containing protein 1 | COMMD1 | Cardiometabolic_II |
| P11532 | Dystrophin | DMD | Cardiometabolic_II |
| P05305 | Endothelin-1 | EDN1 | Cardiometabolic_II |
| Q14160 | Protein scribble homolog | SCRIB | Cardiometabolic_II |
| Q8WZ75 | Roundabout homolog 4 | ROBO4 | Cardiometabolic_II |
| P55809 | Succinyl-CoA:3-ketoacid coenzyme A transferase 1, mitochondrial | OXCT1 | Cardiometabolic_II |
| Q9BY49 | Peroxisomal trans-2-enoyl-CoA reductase | PECR | Cardiometabolic_II |
| Q9NZN3 | EH domain-containing protein 3 | EHD3 | Cardiometabolic_II |
| P14902 | Indoleamine 2,3-dioxygenase 1 | IDO1 | Cardiometabolic_II |
| Q96C92 | Endosome-associated-trafficking regulator 1 | ENTR1 | Cardiometabolic_II |
| O75506 | Heat shock factor-binding protein 1 | HSBP1 | Cardiometabolic_II |
| P01225 | Follitropin subunit beta | FSHB | Cardiometabolic_II |
| O95980 | Reversion-inducing cysteine-rich protein with Kazal motifs | RECK | Cardiometabolic_II |
| Q8NC42 | E3 ubiquitin-protein ligase RNF149 | RNF149 | Cardiometabolic_II |
| Q9H7C9 | Mth938 domain-containing protein | AAMDC | Cardiometabolic_II |
| Q8TAE8 | Growth arrest and DNA damage-inducible proteins-interacting protein 1 | GADD45GIP1 | Cardiometabolic_II |
| Q5GAN6 | Inactive ribonuclease-like protein 10 | RNASE10 | Cardiometabolic_II |
| P30084 | Enoyl-CoA hydratase, mitochondrial | ECHS1 | Cardiometabolic_II |
| Q5SW79 | Centrosomal protein of 170 kDa | CEP170 | Cardiometabolic_II |
| P50053 | Ketohexokinase | KHK | Cardiometabolic_II |
| O75348 | V-type proton ATPase subunit G 1 | ATP6V1G1 | Cardiometabolic_II |
| Q53T59 | HCLS1-binding protein 3 | HS1BP3 | Cardiometabolic_II |
| Q8IVF2 | Protein AHNAK2 | AHNAK2 | Cardiometabolic_II |
| P12829 | Myosin light chain 4 | MYL4 | Cardiometabolic_II |
| Q0VD83 | Apolipoprotein B receptor | APOBR | Cardiometabolic_II |
| O75061 | Putative tyrosine-protein phosphatase auxilin | DNAJC6 | Cardiometabolic_II |
| Q8WUF8 | Cotranscriptional regulator FAM172A | FAM172A | Cardiometabolic_II |
| Q13137 | Calcium-binding and coiled-coil domain-containing protein 2 | CALCOCO2 | Cardiometabolic_II |
| P58107 | Epiplakin | EPPK1 | Cardiometabolic_II |
| P08590 | Myosin light chain 3 | MYL3 | Cardiometabolic_II |
| Q86X76 | Deaminated glutathione amidase | NIT1 | Cardiometabolic_II |
| P0DPI2 | Glutamine amidotransferase-like class 1 domain-containing protein 3B, mitochondrial | GATD3 | Cardiometabolic_II |
| Q12841 | Follistatin-related protein 1 | FSTL1 | Cardiometabolic_II |
| Q9NR61 | Delta-like protein 4 | DLL4 | Cardiometabolic_II |
| Q6YN16 | Hydroxysteroid dehydrogenase-like protein 2 | HSDL2 | Cardiometabolic_II |
| Q6ZRY4 | RNA-binding protein with multiple splicing 2 | RBPMS2 | Cardiometabolic_II |
| P43487 | Ran-specific GTPase-activating protein | RANBP1 | Cardiometabolic_II |
| Q92835 | Phosphatidylinositol 3,4,5-trisphosphate 5-phosphatase 1 | INPP5D | Cardiometabolic_II |
| Q09666 | Neuroblast differentiation-associated protein AHNAK | AHNAK | Cardiometabolic_II |
| Q9NYZ4 | Sialic acid-binding Ig-like lectin 8 | SIGLEC8 | Cardiometabolic_II |
| O60476 | Mannosyl-oligosaccharide 1,2-alpha-mannosidase IB | MAN1A2 | Cardiometabolic_II |
| P07355 | Annexin A2 | ANXA2 | Cardiometabolic_II |
| P07492 | Gastrin-releasing peptide | GRP | Cardiometabolic_II |
| P21754 | Zona pellucida sperm-binding protein 3 | ZP3 | Cardiometabolic_II |
| P07098 | Gastric triacylglycerol lipase | LIPF | Cardiometabolic_II |
| Q9H3K6 | BolA-like protein 2 | BOLA2_BOLA2B | Cardiometabolic_II |
| Q16621 | Transcription factor NF-E2 45 kDa subunit | NFE2 | Cardiometabolic_II |
| O94979 | Protein transport protein Sec31A | SEC31A | Cardiometabolic_II |
| P20042 | Eukaryotic translation initiation factor 2 subunit 2 | EIF2S2 | Cardiometabolic_II |
| Q9UJ70 | N-acetyl-D-glucosamine kinase | NAGK | Cardiometabolic_II |
| O95825 | Quinone oxidoreductase-like protein 1 | CRYZL1 | Cardiometabolic_II |
| Q6UWR7 | Glycerophosphocholine cholinephosphodiesterase ENPP6 | ENPP6 | Cardiometabolic_II |
| Q9BV79 | Enoyl-[acyl-carrier-protein] reductase, mitochondrial | MECR | Cardiometabolic_II |
| Q6UY14 | ADAMTS-like protein 4 | ADAMTSL4 | Cardiometabolic_II |
| Q5FWE3 | Proline-rich transmembrane protein 3 | PRRT3 | Cardiometabolic_II |
| Q8IWT1 | Sodium channel subunit beta-4 | SCN4B | Cardiometabolic_II |
| Q9HB40 | Retinoid-inducible serine carboxypeptidase | SCPEP1 | Cardiometabolic_II |
| Q96DR5 | BPI fold-containing family A member 2 | BPIFA2 | Cardiometabolic_II |
| P07942 | Laminin subunit beta-1 | LAMB1 | Cardiometabolic_II |
| Q8NFL0 | UDP-GlcNAc:betaGal beta-1,3-N-acetylglucosaminyltransferase 7 | B3GNT7 | Cardiometabolic_II |
| P01189 | Pro-opiomelanocortin | POMC | Cardiometabolic_II |
| Q7L266 | Isoaspartyl peptidase/L-asparaginase | ASRGL1 | Cardiometabolic_II |
| P50914 | 60S ribosomal protein L14 | RPL14 | Cardiometabolic_II |
| P09543 | 2',3'-cyclic-nucleotide 3'-phosphodiesterase | CNP | Cardiometabolic_II |
| P02458 | Collagen alpha-1(II) chain | COL2A1 | Cardiometabolic_II |
| Q24JP5 | Transmembrane protein 132A | TMEM132A | Cardiometabolic_II |
| P09681 | Gastric inhibitory polypeptide | GIP | Cardiometabolic_II |
| Q9UBQ7 | Glyoxylate reductase/hydroxypyruvate reductase | GRHPR | Cardiometabolic_II |
| Q9BVM4 | Gamma-glutamylaminecyclotransferase | GGACT | Cardiometabolic_II |
| P33681 | T-lymphocyte activation antigen CD80 | CD80 | Cardiometabolic_II |
| Q12982 | BCL2/adenovirus E1B 19 kDa protein-interacting protein 2 | BNIP2 | Cardiometabolic_II |
| Q96DC8 | Enoyl-CoA hydratase domain-containing protein 3, mitochondrial | ECHDC3 | Cardiometabolic_II |
| Q8TCD5 | 5'(3')-deoxyribonucleotidase, cytosolic type | NT5C | Cardiometabolic_II |
| Q7Z7M9 | Polypeptide N-acetylgalactosaminyltransferase 5 | GALNT5 | Cardiometabolic_II |
| Q00872 | Myosin-binding protein C, slow-type | MYBPC1 | Cardiometabolic_II |
| Q14914 | Prostaglandin reductase 1 | PTGR1 | Cardiometabolic_II |
| Q6UW49 | Sperm equatorial segment protein 1 | SPESP1 | Cardiometabolic_II |
| P51511 | Matrix metalloproteinase-15 | MMP15 | Cardiometabolic_II |
| P08138 | Tumor necrosis factor receptor superfamily member 16 | NGFR | Cardiometabolic_II |
| P15502 | Elastin | ELN | Cardiometabolic_II |
| Q8NDI1 | EH domain-binding protein 1 | EHBP1 | Cardiometabolic_II |
| O43681 | ATPase GET3 | GET3 | Cardiometabolic_II |
| O75223 | Gamma-glutamylcyclotransferase | GGCT | Cardiometabolic_II |
| O43405 | Cochlin | COCH | Cardiometabolic_II |
| Q9Y2E5 | Epididymis-specific alpha-mannosidase | MAN2B2 | Cardiometabolic_II |
| P02008 | Hemoglobin subunit zeta | HBZ | Cardiometabolic_II |
| P23919 | Thymidylate kinase | DTYMK | Cardiometabolic_II |
| Q7Z304 | MAM domain-containing protein 2 | MAMDC2 | Cardiometabolic_II |
| P78539 | Sushi repeat-containing protein SRPX | SRPX | Cardiometabolic_II |
| Q8N4F0 | BPI fold-containing family B member 2 | BPIFB2 | Cardiometabolic_II |
| P54687 | Branched-chain-amino-acid aminotransferase, cytosolic | BCAT1 | Cardiometabolic_II |
| Q969H8 | Myeloid-derived growth factor | MYDGF | Cardiometabolic_II |
| Q96EM0 | Trans-3-hydroxy-L-proline dehydratase | L3HYPDH | Cardiometabolic_II |
| Q9BXN1 | Asporin | ASPN | Cardiometabolic_II |
| Q13428 | Treacle protein | TCOF1 | Cardiometabolic_II |
| P14854 | Cytochrome c oxidase subunit 6B1 | COX6B1 | Cardiometabolic_II |
| P16035 | Metalloproteinase inhibitor 2 | TIMP2 | Cardiometabolic_II |
| P53674 | Beta-crystallin B1 | CRYBB1 | Cardiometabolic_II |
| O14960 | Leukocyte cell-derived chemotaxin-2 | LECT2 | Cardiometabolic_II |
| O14933 | Ubiquitin/ISG15-conjugating enzyme E2 L6 | UBE2L6 | Cardiometabolic_II |
| Q8N436 | Inactive carboxypeptidase-like protein X2 | CPXM2 | Cardiometabolic_II |
| Q13442 | 28 kDa heat- and acid-stable phosphoprotein | PDAP1 | Cardiometabolic_II |
| P23467 | Receptor-type tyrosine-protein phosphatase beta | PTPRB | Cardiometabolic_II |
| O75154 | Rab11 family-interacting protein 3 | RAB11FIP3 | Cardiometabolic_II |
| Q6NUS6 | Tectonic-3 | TCTN3 | Cardiometabolic_II |
| Q96FZ7 | Charged multivesicular body protein 6 | CHMP6 | Cardiometabolic_II |
| P98161 | Polycystin-1 | PKD1 | Cardiometabolic_II |
| Q9BXD5 | N-acetylneuraminate lyase | NPL | Cardiometabolic_II |
| Q9P2J2 | Protein turtle homolog A | IGSF9 | Cardiometabolic_II |
| Q9BW04 | Specifically androgen-regulated gene protein | SARG | Cardiometabolic_II |
| Q8WWV6 | High affinity immunoglobulin alpha and immunoglobulin mu Fc receptor | FCAMR | Cardiometabolic_II |
| O75711 | Scrapie-responsive protein 1 | SCRG1 | Cardiometabolic_II |
| Q6UXI7 | Vitrin | VIT | Cardiometabolic_II |
| P29692 | Elongation factor 1-delta | EEF1D | Cardiometabolic_II |
| Q9NQR4 | Omega-amidase NIT2 | NIT2 | Cardiometabolic_II |
| Q9BQS7 | Hephaestin | HEPH | Cardiometabolic_II |
| Q9H3S4 | Thiamin pyrophosphokinase 1 | TPK1 | Cardiometabolic_II |
| Q96C24 | Synaptotagmin-like protein 4 | SYTL4 | Cardiometabolic_II |
| O60234 | Glia maturation factor gamma | GMFG | Cardiometabolic_II |
| P51688 | N-sulphoglucosamine sulphohydrolase | SGSH | Cardiometabolic_II |
| Q969X0 | RILP-like protein 2 | RILPL2 | Cardiometabolic_II |
| P23471 | Receptor-type tyrosine-protein phosphatase zeta | PTPRZ1 | Cardiometabolic_II |
| P52209 | 6-phosphogluconate dehydrogenase, decarboxylating | PGD | Cardiometabolic_II |
| P32320 | Cytidine deaminase | CDA | Cardiometabolic_II |
| Q6PI73 | Leukocyte immunoglobulin-like receptor subfamily A member 6 | LILRA6 | Cardiometabolic_II |
| P08582 | Melanotransferrin | MELTF | Cardiometabolic_II |
| Q96MK3 | Pseudokinase FAM20A | FAM20A | Cardiometabolic_II |
| Q8IZF2 | Adhesion G protein-coupled receptor F5 | ADGRF5 | Cardiometabolic_II |
| P49593 | Protein phosphatase 1F | PPM1F | Cardiometabolic_II |
| P05413 | Fatty acid-binding protein, heart | FABP3 | Cardiometabolic_II |
| Q9UBR1 | Beta-ureidopropionase | UPB1 | Cardiometabolic_II |
| Q15388 | Mitochondrial import receptor subunit TOM20 homolog | TOMM20 | Cardiometabolic_II |
| O00194 | Ras-related protein Rab-27B | RAB27B | Cardiometabolic_II |
| Q6ZMM2 | ADAMTS-like protein 5 | ADAMTSL5 | Cardiometabolic_II |
| P45954 | Short/branched chain specific acyl-CoA dehydrogenase, mitochondrial | ACADSB | Cardiometabolic_II |
| P61026 | Ras-related protein Rab-10 | RAB10 | Cardiometabolic_II |
| P62072 | Mitochondrial import inner membrane translocase subunit Tim10 | TIMM10 | Cardiometabolic_II |
| P07093 | Glia-derived nexin | SERPINE2 | Cardiometabolic_II |
| Q7Z7K0 | COX assembly mitochondrial protein homolog | CMC1 | Cardiometabolic_II |
| Q96AG4 | Leucine-rich repeat-containing protein 59 | LRRC59 | Cardiometabolic_II |
| P40199 | Carcinoembryonic antigen-related cell adhesion molecule 6 | CEACAM6 | Cardiometabolic_II |
| P16410 | Cytotoxic T-lymphocyte protein 4 | CTLA4 | Cardiometabolic_II |
| P07288 | Prostate-specific antigen | KLK3 | Cardiometabolic_II |
| Q6H9L7 | Isthmin-2 | ISM2 | Cardiometabolic_II |
| Q9Y303 | N-acetylglucosamine-6-phosphate deacetylase | AMDHD2 | Cardiometabolic_II |
| P13796 | Plastin-2 | LCP1 | Cardiometabolic_II |
| P02730 | Band 3 anion transport protein | SLC4A1 | Cardiometabolic_II |
| Q86TH1 | ADAMTS-like protein 2 | ADAMTSL2 | Cardiometabolic_II |
| P30046 | D-dopachrome decarboxylase | DDT | Cardiometabolic_II |
| P23560 | Brain-derived neurotrophic factor | BDNF | Cardiometabolic_II |
| Q9NRR1 | Cytokine-like protein 1 | CYTL1 | Cardiometabolic_II |
| P13667 | Protein disulfide-isomerase A4 | PDIA4 | Cardiometabolic_II |
| Q8N6C8 | Leukocyte immunoglobulin-like receptor subfamily A member 3 | LILRA3 | Cardiometabolic_II |
| Q02817 | Mucin-2 | MUC2 | Cardiometabolic_II |
| P98095 | Fibulin-2 | FBLN2 | Cardiometabolic_II |
| P02461 | Collagen alpha-1(III) chain | COL3A1 | Cardiometabolic_II |
| P08575 | Receptor-type tyrosine-protein phosphatase C | PTPRC | Cardiometabolic_II |
| P13727 | Bone marrow proteoglycan | PRG2 | Cardiometabolic_II |
| P35579 | Myosin-9 | MYH9 | Cardiometabolic_II |
| Q9Y2Y8 | Proteoglycan 3 | PRG3 | Cardiometabolic_II |
| P32971 | Tumor necrosis factor ligand superfamily member 8 | TNFSF8 | Cardiometabolic_II |
| P30405 | Peptidyl-prolyl cis-trans isomerase F, mitochondrial | PPIF | Cardiometabolic_II |
| Q6IBS0 | Twinfilin-2 | TWF2 | Cardiometabolic_II |
| Q8N114 | Protein shisa-5 | SHISA5 | Cardiometabolic_II |
| O43280 | Trehalase | TREH | Cardiometabolic_II |
| P02818 | Osteocalcin | BGLAP | Cardiometabolic_II |
| P47972 | Neuronal pentraxin-2 | NPTX2 | Cardiometabolic_II |
| Q96NZ9 | Proline-rich acidic protein 1 | PRAP1 | Cardiometabolic_II |
| Q96CG8 | Collagen triple helix repeat-containing protein 1 | CTHRC1 | Cardiometabolic_II |
| Q9HCU0 | Endosialin | CD248 | Cardiometabolic_II |
| Q9BYJ0 | Fibroblast growth factor-binding protein 2 | FGFBP2 | Cardiometabolic_II |
| P39059 | Collagen alpha-1(XV) chain | COL15A1 | Cardiometabolic_II |
| Q6UVK1 | Chondroitin sulfate proteoglycan 4 | CSPG4 | Cardiometabolic_II |
| Q6UWP8 | Suprabasin | SBSN | Cardiometabolic_II |
| O75339 | Cartilage intermediate layer protein 1 | CILP | Cardiometabolic_II |
| P12277 | Creatine kinase B-type | CKB | Cardiometabolic_II |
| Q8WWQ8 | Stabilin-2 | STAB2 | Cardiometabolic_II |
| Q8TDY8 | Immunoglobulin superfamily DCC subclass member 4 | IGDCC4 | Cardiometabolic_II |
| P10645 | Chromogranin-A | CHGA | Cardiometabolic_II |
| P55000 | Secreted Ly-6/uPAR-related protein 1 | SLURP1 | Cardiometabolic_II |
| Q9H2X3 | C-type lectin domain family 4 member M | CLEC4M | Cardiometabolic_II |
| Q9Y646 | Carboxypeptidase Q | CPQ | Cardiometabolic_II |
| Q04721 | Neurogenic locus notch homolog protein 2 | NOTCH2 | Cardiometabolic_II |
| O95965 | Integrin beta-like protein 1 | ITGBL1 | Cardiometabolic_II |
| Q9Y251 | Heparanase | HPSE | Cardiometabolic_II |
| Q15063 | Periostin | POSTN | Cardiometabolic_II |
| P08217 | Chymotrypsin-like elastase family member 2A | CELA2A | Cardiometabolic_II |
| Q9UQP3 | Tenascin-N | TNN | Cardiometabolic_II |
| P17900 | Ganglioside GM2 activator | GM2A | Cardiometabolic_II |
| P37837 | Transaldolase | TALDO1 | Cardiometabolic_II |
| Q13510 | Acid ceramidase | ASAH1 | Cardiometabolic_II |
| P11279 | Lysosome-associated membrane glycoprotein 1 | LAMP1 | Cardiometabolic_II |
| P17174 | Aspartate aminotransferase, cytoplasmic | GOT1 | Cardiometabolic_II |
| P61916 | NPC intracellular cholesterol transporter 2 | NPC2 | Cardiometabolic_II |
| P07602 | Prosaposin | PSAP | Cardiometabolic_II |
| P60568 | Interleukin-2 | IL2 | Inflammation |
| Q13651 | Interleukin-10 receptor subunit alpha | IL10RA | Inflammation |
| Q13219 | Pappalysin-1 | PAPPA | Inflammation |
| Q9UHF4 | Interleukin-20 receptor subunit alpha | IL20RA | Inflammation |
| P63241 | Eukaryotic translation initiation factor 5A-1 | EIF5A | Inflammation |
| P05412 | Transcription factor AP-1 | JUN | Inflammation |
| Q96AX2 | Ras-related protein Rab-37 | RAB37 | Inflammation |
| P05112 | Interleukin-4 | IL4 | Inflammation |
| P01584 | Interleukin-1 beta | IL1B | Inflammation |
| O95760 | Interleukin-33 | IL33 | Inflammation |
| Q8NHJ6 | Leukocyte immunoglobulin-like receptor subfamily B member 4 | LILRB4 | Inflammation |
| P35225 | Interleukin-13 | IL13 | Inflammation |
| P22301 | Interleukin-10 | IL10 | Inflammation |
| P27540 | Aryl hydrocarbon receptor nuclear translocator | ARNT | Inflammation |
| O95379 | Tumor necrosis factor alpha-induced protein 8 | TNFAIP8 | Inflammation |
| Q8WV07 | Protein LTO1 homolog | LTO1 | Inflammation |
| O43707 | Alpha-actinin-4 | ACTN4 | Inflammation |
| P28838 | Cytosol aminopeptidase | LAP3 | Inflammation |
| Q9NP70 | Ameloblastin | AMBN | Inflammation |
| Q6UXK5 | Leucine-rich repeat neuronal protein 1 | LRRN1 | Inflammation |
| Q9HCU5 | Prolactin regulatory element-binding protein | PREB | Inflammation |
| Q13007 | Interleukin-24 | IL24 | Inflammation |
| Q9UPV0 | Centrosomal protein of 164 kDa | CEP164 | Inflammation |
| O60934 | Nibrin | NBN | Inflammation |
| Q96P31 | Fc receptor-like protein 3 | FCRL3 | Inflammation |
| Q9Y478 | 5'-AMP-activated protein kinase subunit beta-1 | PRKAB1 | Inflammation |
| Q8TCS8 | Polyribonucleotide nucleotidyltransferase 1, mitochondrial | PNPT1 | Inflammation |
| Q5T4W7 | Artemin | ARTN | Inflammation |
| Q5R372 | Rab GTPase-activating protein 1-like | RABGAP1L | Inflammation |
| Q969V3 | Nicalin | NCLN | Inflammation |
| Q8N6P7 | Interleukin-22 receptor subunit alpha-1 | IL22RA1 | Inflammation |
| P14784 | Interleukin-2 receptor subunit beta | IL2RB | Inflammation |
| Q13459 | Unconventional myosin-IXb | MYO9B | Inflammation |
| P19801 | Amiloride-sensitive amine oxidase [copper-containing] | AOC1 | Inflammation |
| Q9NYY1 | Interleukin-20 | IL20 | Inflammation |
| P57771 | Regulator of G-protein signaling 8 | RGS8 | Inflammation |
| P20809 | Interleukin-11 | IL11 | Inflammation |
| Q96PD4 | Interleukin-17F | IL17F | Inflammation |
| O76038 | Secretagogin | SCGN | Inflammation |
| O95715 | C-X-C motif chemokine 14 | CXCL14 | Inflammation |
| Q03426 | Mevalonate kinase | MVK | Inflammation |
| O14904 | Protein Wnt-9a | WNT9A | Inflammation |
| P26951 | Interleukin-3 receptor subunit alpha | IL3RA | Inflammation |
| Q9Y3P8 | Signaling threshold-regulating transmembrane adapter 1 | SIT1 | Inflammation |
| Q96DB9 | FXYD domain-containing ion transport regulator 5 | FXYD5 | Inflammation |
| P48061 | Stromal cell-derived factor 1 | CXCL12 | Inflammation |
| Q99748 | Neurturin | NRTN | Inflammation |
| Q13574 | Diacylglycerol kinase zeta | DGKZ | Inflammation |
| Q9Y2J8 | Protein-arginine deiminase type-2 | PADI2 | Inflammation |
| Q04759 | Protein kinase C theta type | PRKCQ | Inflammation |
| Q16552 | Interleukin-17A | IL17A | Inflammation |
| Q12968 | Nuclear factor of activated T-cells, cytoplasmic 3 | NFATC3 | Inflammation |
| Q14435 | Polypeptide N-acetylgalactosaminyltransferase 3 | GALNT3 | Inflammation |
| P05113 | Interleukin-5 | IL5 | Inflammation |
| P01375 | Tumor necrosis factor | TNF | Inflammation |
| Q92844 | TRAF family member-associated NF-kappa-B activator | TANK | Inflammation |
| O43597 | Protein sprouty homolog 2 | SPRY2 | Inflammation |
| P13693 | Translationally-controlled tumor protein | TPT1 | Inflammation |
| Q9P0M4 | Interleukin-17C | IL17C | Inflammation |
| Q7Z739 | YTH domain-containing family protein 3 | YTHDF3 | Inflammation |
| P42768 | Wiskott-Aldrich syndrome protein | WAS | Inflammation |
| Q96RJ3 | Tumor necrosis factor receptor superfamily member 13C | TNFRSF13C | Inflammation |
| Q8TAD2 | Interleukin-17D | IL17D | Inflammation |
| Q7Z6M3 | Allergin-1 | MILR1 | Inflammation |
| P30048 | Thioredoxin-dependent peroxide reductase, mitochondrial | PRDX3 | Inflammation |
| Q05084 | Islet cell autoantigen 1 | ICA1 | Inflammation |
| P51617 | Interleukin-1 receptor-associated kinase 1 | IRAK1 | Inflammation |
| P42701 | Interleukin-12 receptor subunit beta-1 | IL12RB1 | Inflammation |
| Q9HB29 | Interleukin-1 receptor-like 2 | IL1RL2 | Inflammation |
| P01583 | Interleukin-1 alpha | IL1A | Inflammation |
| P32456 | Guanylate-binding protein 2 | GBP2 | Inflammation |
| P12034 | Fibroblast growth factor 5 | FGF5 | Inflammation |
| P09919 | Granulocyte colony-stimulating factor | CSF3 | Inflammation |
| Q9BXJ7 | Protein amnionless | AMN | Inflammation |
| P18564 | Integrin beta-6 | ITGB6 | Inflammation |
| P01591 | Immunoglobulin J chain | JCHAIN | Inflammation |
| P01579 | Interferon gamma | IFNG | Inflammation |
| Q13291 | Signaling lymphocytic activation molecule | SLAMF1 | Inflammation |
| Q8N8S7 | Protein enabled homolog | ENAH | Inflammation |
| Q13261 | Interleukin-15 receptor subunit alpha | IL15RA | Inflammation |
| P09874 | Poly [ADP-ribose] polymerase 1 | PARP1 | Inflammation |
| Q0Z7S8 | Fatty acid-binding protein 9 | FABP9 | Inflammation |
| P78362 | SRSF protein kinase 2 | SRPK2 | Inflammation |
| P09038 | Fibroblast growth factor 2 | FGF2 | Inflammation |
| O43736 | Integral membrane protein 2A | ITM2A | Inflammation |
| O14867 | Transcription regulator protein BACH1 | BACH1 | Inflammation |
| Q8IU57 | Interferon lambda receptor 1 | IFNLR1 | Inflammation |
| Q12933 | TNF receptor-associated factor 2 | TRAF2 | Inflammation |
| Q06520 | Bile salt sulfotransferase | SULT2A1 | Inflammation |
| O60575 | Serine protease inhibitor Kazal-type 4 | SPINK4 | Inflammation |
| Q9Y5A7 | NEDD8 ultimate buster 1 | NUB1 | Inflammation |
| O60542 | Persephin | PSPN | Inflammation |
| P30838 | Aldehyde dehydrogenase, dimeric NADP-preferring | ALDH3A1 | Inflammation |
| O43521-2 | Bcl-2-like protein 11, Isoform BimL | BCL2L11 | Inflammation |
| O60880 | SH2 domain-containing protein 1A | SH2D1A | Inflammation |
| Q12778 | Forkhead box protein O1 | FOXO1 | Inflammation |
| Q7L8A9 | Tubulinyl-Tyr carboxypeptidase 1 | VASH1 | Inflammation |
| P55957 | BH3-interacting domain death agonist | BID | Inflammation |
| Q6UB28 | Methionine aminopeptidase 1D, mitochondrial | METAP1D | Inflammation |
| P01903 | HLA class II histocompatibility antigen, DR alpha chain | HLA-DRA | Inflammation |
| Q92609 | TBC1 domain family member 5 | TBC1D5 | Inflammation |
| P01588 | Erythropoietin | EPO | Inflammation |
| P80098 | C-C motif chemokine 7 | CCL7 | Inflammation |
| Q9UN19 | Dual adapter for phosphotyrosine and 3-phosphotyrosine and 3-phosphoinositide | DAPP1 | Inflammation |
| Q9UNE0 | Tumor necrosis factor receptor superfamily member EDAR | EDAR | Inflammation |
| Q9C035 | Tripartite motif-containing protein 5 | TRIM5 | Inflammation |
| Q8N608 | Inactive dipeptidyl peptidase 10 | DPP10 | Inflammation |
| P23229 | Integrin alpha-6 | ITGA6 | Inflammation |
| Q6DN72 | Fc receptor-like protein 6 | FCRL6 | Inflammation |
| P33241 | Lymphocyte-specific protein 1 | LSP1 | Inflammation |
| Q9UNK0 | Syntaxin-8 | STX8 | Inflammation |
| P13747 | HLA class I histocompatibility antigen, alpha chain E | HLA-E | Inflammation |
| P19474 | E3 ubiquitin-protein ligase TRIM21 | TRIM21 | Inflammation |
| O75475 | PC4 and SFRS1-interacting protein | PSIP1 | Inflammation |
| P13232 | Interleukin-7 | IL7 | Inflammation |
| Q8IVG5 | Sterile alpha motif domain-containing protein 9-like | SAMD9L | Inflammation |
| Q96LC7 | Sialic acid-binding Ig-like lectin 10 | SIGLEC10 | Inflammation |
| B1AKI9 | Isthmin-1 | ISM1 | Inflammation |
| Q6ZMH5 | Zinc transporter ZIP5 | SLC39A5 | Inflammation |
| P78410 | Butyrophilin subfamily 3 member A2 | BTN3A2 | Inflammation |
| P12872 | Promotilin | MLN | Inflammation |
| Q12765 | Secernin-1 | SCRN1 | Inflammation |
| P58294 | Prokineticin-1 | PROK1 | Inflammation |
| Q9Y6K9 | NF-kappa-B essential modulator | IKBKG | Inflammation |
| O95644 | Nuclear factor of activated T-cells, cytoplasmic 1 | NFATC1 | Inflammation |
| Q9Y258 | C-C motif chemokine 26 | CCL26 | Inflammation |
| Q8WTT0 | C-type lectin domain family 4 member C | CLEC4C | Inflammation |
| Q3KPI0 | Carcinoembryonic antigen-related cell adhesion molecule 21 | CEACAM21 | Inflammation |
| Q9BT73 | Proteasome assembly chaperone 3 | PSMG3 | Inflammation |
| P20849 | Collagen alpha-1(IX) chain | COL9A1 | Inflammation |
| Q9HD26 | Golgi-associated PDZ and coiled-coil motif-containing protein | GOPC | Inflammation |
| P52564 | Dual specificity mitogen-activated protein kinase kinase 6 | MAP2K6 | Inflammation |
| Q9H0P0 | Cytosolic 5'-nucleotidase 3A | NT5C3A | Inflammation |
| Q9NZN5 | Rho guanine nucleotide exchange factor 12 | ARHGEF12 | Inflammation |
| P42575 | Caspase-2 | CASP2 | Inflammation |
| Q9UHC6 | Contactin-associated protein-like 2 | CNTNAP2 | Inflammation |
| P45984 | Mitogen-activated protein kinase 9 | MAPK9 | Inflammation |
| P11274 | Breakpoint cluster region protein | BCR | Inflammation |
| Q9UDT6 | CAP-Gly domain-containing linker protein 2 | CLIP2 | Inflammation |
| Q14242 | P-selectin glycoprotein ligand 1 | SELPLG | Inflammation |
| P78310 | Coxsackievirus and adenovirus receptor | CXADR | Inflammation |
| P40933 | Interleukin-15 | IL15 | Inflammation |
| P05231 | Interleukin-6 | IL6 | Inflammation |
| P24071 | Immunoglobulin alpha Fc receptor | FCAR | Inflammation |
| Q01151 | CD83 antigen | CD83 | Inflammation |
| O76036 | Natural cytotoxicity triggering receptor 1 | NCR1 | Inflammation |
| P19878 | Neutrophil cytosol factor 2 | NCF2 | Inflammation |
| P23582 | C-type natriuretic peptide | NPPC | Inflammation |
| Q9NRJ3 | C-C motif chemokine 28 | CCL28 | Inflammation |
| P26022 | Pentraxin-related protein PTX3 | PTX3 | Inflammation |
| Q03431 | Parathyroid hormone/parathyroid hormone-related peptide receptor | PTH1R | Inflammation |
| Q9GZT9 | Egl nine homolog 1 | EGLN1 | Inflammation |
| Q9UMR7 | C-type lectin domain family 4 member A | CLEC4A | Inflammation |
| P13725 | Oncostatin-M | OSM | Inflammation |
| P28845 | Corticosteroid 11-beta-dehydrogenase isozyme 1 | HSD11B1 | Inflammation |
| P24394 | Interleukin-4 receptor subunit alpha | IL4R | Inflammation |
| Q9NWZ3 | Interleukin-1 receptor-associated kinase 4 | IRAK4 | Inflammation |
| Q14773 | Intercellular adhesion molecule 4 | ICAM4 | Inflammation |
| Q16698 | 2,4-dienoyl-CoA reductase, mitochondrial | DECR1 | Inflammation |
| O60449 | Lymphocyte antigen 75 | LY75 | Inflammation |
| Q9UKX5 | Integrin alpha-11 | ITGA11 | Inflammation |
| O15169 | Axin-1 | AXIN1 | Inflammation |
| P50995 | Annexin A11 | ANXA11 | Inflammation |
| P01730 | T-cell surface glycoprotein CD4 | CD4 | Inflammation |
| Q9NRM6 | Interleukin-17 receptor B | IL17RB | Inflammation |
| P01374 | Lymphotoxin-alpha | LTA | Inflammation |
| Q6ZUJ8 | Phosphoinositide 3-kinase adapter protein 1 | PIK3AP1 | Inflammation |
| P16455 | Methylated-DNA--protein-cysteine methyltransferase | MGMT | Inflammation |
| O94992 | Protein HEXIM1 | HEXIM1 | Inflammation |
| Q6UXB4 | C-type lectin domain family 4 member G | CLEC4G | Inflammation |
| P20783 | Neurotrophin-3 | NTF3 | Inflammation |
| O14788 | Tumor necrosis factor ligand superfamily member 11 | TNFSF11 | Inflammation |
| Q29980_Q29983 | MHC class I polypeptide-related sequence A_MHC class I polypeptide-related sequence B | MICB_MICA | Inflammation |
| Q8NDB2 | B-cell scaffold protein with ankyrin repeats | BANK1 | Inflammation |
| Q8TD46 | Cell surface glycoprotein CD200 receptor 1 | CD200R1 | Inflammation |
| P08727 | Keratin, type I cytoskeletal 19 | KRT19 | Inflammation |
| Q9HCM2 | Plexin-A4 | PLXNA4 | Inflammation |
| P28827 | Receptor-type tyrosine-protein phosphatase mu | PTPRM | Inflammation |
| P32970 | CD70 antigen | CD70 | Inflammation |
| P01135 | Protransforming growth factor alpha | TGFA | Inflammation |
| Q01344 | Interleukin-5 receptor subunit alpha | IL5RA | Inflammation |
| Q9NQ25 | SLAM family member 7 | SLAMF7 | Inflammation |
| P41217 | OX-2 membrane glycoprotein | CD200 | Inflammation |
| P10144 | Granzyme B | GZMB | Inflammation |
| P24001 | Interleukin-32 | IL32 | Inflammation |
| P42702 | Leukemia inhibitory factor receptor | LIFR | Inflammation |
| Q9UIB8 | SLAM family member 5 | CD84 | Inflammation |
| P20340 | Ras-related protein Rab-6A | RAB6A | Inflammation |
| Q8WXI8 | C-type lectin domain family 4 member D | CLEC4D | Inflammation |
| P10147 | C-C motif chemokine 3 | CCL3 | Inflammation |
| P50591 | Tumor necrosis factor ligand superfamily member 10 | TNFSF10 | Inflammation |
| O15455 | Toll-like receptor 3 | TLR3 | Inflammation |
| P80162 | C-X-C motif chemokine 6 | CXCL6 | Inflammation |
| Q08174 | Protocadherin-1 | PCDH1 | Inflammation |
| P37235 | Hippocalcin-like protein 1 | HPCAL1 | Inflammation |
| P29965 | CD40 ligand | CD40LG | Inflammation |
| Q07065 | Cytoskeleton-associated protein 4 | CKAP4 | Inflammation |
| P68106 | Peptidyl-prolyl cis-trans isomerase FKBP1B | FKBP1B | Inflammation |
| P22304 | Iduronate 2-sulfatase | IDS | Inflammation |
| O00273 | DNA fragmentation factor subunit alpha | DFFA | Inflammation |
| P01137 | Transforming growth factor beta-1 proprotein | TGFB1 | Inflammation |
| Q6UXB2 | C-X-C motif chemokine 17 | CXCL17 | Inflammation |
| Q9Y266 | Nuclear migration protein nudC | NUDC | Inflammation |
| O43508 | Tumor necrosis factor ligand superfamily member 12 | TNFSF12 | Inflammation |
| Q04637 | Eukaryotic translation initiation factor 4 gamma 1 | EIF4G1 | Inflammation |
| P35613 | Basigin | BSG | Inflammation |
| O60884 | DnaJ homolog subfamily A member 2 | DNAJA2 | Inflammation |
| Q9BZW8 | Natural killer cell receptor 2B4 | CD244 | Inflammation |
| Q12918 | Killer cell lectin-like receptor subfamily B member 1 | KLRB1 | Inflammation |
| P50452 | Serpin B8 | SERPINB8 | Inflammation |
| P10145 | Interleukin-8 | CXCL8 | Inflammation |
| Q13241 | Natural killer cells antigen CD94 | KLRD1 | Inflammation |
| Q14005 | Pro-interleukin-16 | IL16 | Inflammation |
| O94856 | Neurofascin | NFASC | Inflammation |
| P40259 | B-cell antigen receptor complex-associated protein beta chain | CD79B | Inflammation |
| Q9BXN2 | C-type lectin domain family 7 member A | CLEC7A | Inflammation |
| P20273 | B-cell receptor CD22 | CD22 | Inflammation |
| Q9UQV4 | Lysosome-associated membrane glycoprotein 3 | LAMP3 | Inflammation |
| Q96LA5 | Fc receptor-like protein 2 | FCRL2 | Inflammation |
| O43561 | Linker for activation of T-cells family member 1 | LAT | Inflammation |
| P36959 | GMP reductase 1 | GMPR | Inflammation |
| Q15661 | Tryptase alpha/beta-1 | TPSAB1 | Inflammation |
| Q96SB3 | Neurabin-2 | PPP1R9B | Inflammation |
| P30044 | Peroxiredoxin-5, mitochondrial | PRDX5 | Inflammation |
| P00813 | Adenosine deaminase | ADA | Inflammation |
| Q9Y6Q6 | Tumor necrosis factor receptor superfamily member 11A | TNFRSF11A | Inflammation |
| O95971 | CD160 antigen | CD160 | Inflammation |
| P14317 | Hematopoietic lineage cell-specific protein | HCLS1 | Inflammation |
| P30203 | T-cell differentiation antigen CD6 | CD6 | Inflammation |
| P15692 | Vascular endothelial growth factor A | VEGFA | Inflammation |
| O75077 | Disintegrin and metalloproteinase domain-containing protein 23 | ADAM23 | Inflammation |
| Q13478 | Interleukin-18 receptor 1 | IL18R1 | Inflammation |
| P43489 | Tumor necrosis factor receptor superfamily member 4 | TNFRSF4 | Inflammation |
| P02745 | Complement C1q subcomponent subunit A | C1QA | Inflammation |
| Q99616 | C-C motif chemokine 13 | CCL13 | Inflammation |
| P14210 | Hepatocyte growth factor | HGF | Inflammation |
| Q12866 | Tyrosine-protein kinase Mer | MERTK | Inflammation |
| O00253 | Agouti-related protein | AGRP | Inflammation |
| P43234 | Cathepsin O | CTSO | Inflammation |
| P49771 | Fms-related tyrosine kinase 3 ligand | FLT3LG | Inflammation |
| O43915 | Vascular endothelial growth factor D | VEGFD | Inflammation |
| P12544 | Granzyme A | GZMA | Inflammation |
| Q9H4D0 | Calsyntenin-2 | CLSTN2 | Inflammation |
| P48023 | Tumor necrosis factor ligand superfamily member 6 | FASLG | Inflammation |
| P29460 | Interleukin-12 subunit beta | IL12B | Inflammation |
| Q15517 | Corneodesmosin | CDSN | Inflammation |
| P51671 | Eotaxin | CCL11 | Inflammation |
| Q16719 | Kynureninase | KYNU | Inflammation |
| Q9HBG7 | T-lymphocyte surface antigen Ly-9 | LY9 | Inflammation |
| P78556 | C-C motif chemokine 20 | CCL20 | Inflammation |
| P03956 | Interstitial collagenase | MMP1 | Inflammation |
| P49763 | Placenta growth factor | PGF | Inflammation |
| O15444 | C-C motif chemokine 25 | CCL25 | Inflammation |
| Q07325 | C-X-C motif chemokine 9 | CXCL9 | Inflammation |
| Q92484 | Acid sphingomyelinase-like phosphodiesterase 3a | SMPDL3A | Inflammation |
| P21709 | Ephrin type-A receptor 1 | EPHA1 | Inflammation |
| P15260 | Interferon gamma receptor 1 | IFNGR1 | Inflammation |
| P53634 | Dipeptidyl peptidase 1 | CTSC | Inflammation |
| Q5ZPR3 | CD276 antigen | CD276 | Inflammation |
| Q9UJU6 | Drebrin-like protein | DBNL | Inflammation |
| P30613 | Pyruvate kinase PKLR | PKLR | Inflammation |
| O43639 | Cytoplasmic protein NCK2 | NCK2 | Inflammation |
| P35625 | Metalloproteinase inhibitor 3 | TIMP3 | Inflammation |
| P47712 | Cytosolic phospholipase A2 | PLA2G4A | Inflammation |
| O00585 | C-C motif chemokine 21 | CCL21 | Inflammation |
| P09238 | Stromelysin-2 | MMP10 | Inflammation |
| P11684 | Uteroglobin | SCGB1A1 | Inflammation |
| Q6UWV6 | Ectonucleotide pyrophosphatase/phosphodiesterase family member 7 | ENPP7 | Inflammation |
| Q9BZZ2 | Sialoadhesin | SIGLEC1 | Inflammation |
| P25116 | Proteinase-activated receptor 1 | F2R | Inflammation |
| P09326 | CD48 antigen | CD48 | Inflammation |
| P55773 | C-C motif chemokine 23 | CCL23 | Inflammation |
| Q14116 | Interleukin-18 | IL18 | Inflammation |
| P13236 | C-C motif chemokine 4 | CCL4 | Inflammation |
| Q08334 | Interleukin-10 receptor subunit beta | IL10RB | Inflammation |
| P02778 | C-X-C motif chemokine 10 | CXCL10 | Inflammation |
| P01133 | Pro-epidermal growth factor | EGF | Inflammation |
| O75462 | Cytokine receptor-like factor 1 | CRLF1 | Inflammation |
| P18510 | Interleukin-1 receptor antagonist protein | IL1RN | Inflammation |
| Q9NZV1 | Cysteine-rich motor neuron 1 protein | CRIM1 | Inflammation |
| O14836 | Tumor necrosis factor receptor superfamily member 13B | TNFRSF13B | Inflammation |
| Q9BY76 | Angiopoietin-related protein 4 | ANGPTL4 | Inflammation |
| O76096 | Cystatin-F | CST7 | Inflammation |
| P21860 | Receptor tyrosine-protein kinase erbB-3 | ERBB3 | Inflammation |
| Q99435 | Protein kinase C-binding protein NELL2 | NELL2 | Inflammation |
| P55145 | Mesencephalic astrocyte-derived neurotrophic factor | MANF | Inflammation |
| Q14210 | Lymphocyte antigen 6D | LY6D | Inflammation |
| P29279 | CCN family member 2 | CCN2 | Inflammation |
| Q9HC38 | Glyoxalase domain-containing protein 4 | GLOD4 | Inflammation |
| Q99685 | Monoglyceride lipase | MGLL | Inflammation |
| Q8NFT8 | Delta and Notch-like epidermal growth factor-related receptor | DNER | Inflammation |
| Q7KYR7 | Butyrophilin subfamily 2 member A1 | BTN2A1 | Inflammation |
| P34896 | Serine hydroxymethyltransferase, cytosolic | SHMT1 | Inflammation |
| O95750 | Fibroblast growth factor 19 | FGF19 | Inflammation |
| P19256 | Lymphocyte function-associated antigen 3 | CD58 | Inflammation |
| P29350 | Tyrosine-protein phosphatase non-receptor type 6 | PTPN6 | Inflammation |
| P0DMV8 | Heat shock 70 kDa protein 1A | HSPA1A | Inflammation |
| P09603 | Macrophage colony-stimulating factor 1 | CSF1 | Inflammation |
| O43291 | Kunitz-type protease inhibitor 2 | SPINT2 | Inflammation |
| P12532 | Creatine kinase U-type, mitochondrial | CKMT1A_CKMT1B | Inflammation |
| Q9Y3D6 | Mitochondrial fission 1 protein | FIS1 | Inflammation |
| Q96PL1 | Secretoglobin family 3A member 2 | SCGB3A2 | Inflammation |
| P25942 | Tumor necrosis factor receptor superfamily member 5 | CD40 | Inflammation |
| Q99983 | Osteomodulin | OMD | Inflammation |
| Q9UKU9 | Angiopoietin-related protein 2 | ANGPTL2 | Inflammation |
| P22466 | Galanin peptides | GAL | Inflammation |
| Q8WXD2 | Secretogranin-3 | SCG3 | Inflammation |
| Q9NR12 | PDZ and LIM domain protein 7 | PDLIM7 | Inflammation |
| Q9H3U7 | SPARC-related modular calcium-binding protein 2 | SMOC2 | Inflammation |
| Q9NZC2 | Triggering receptor expressed on myeloid cells 2 | TREM2 | Inflammation |
| Q8WU39 | Marginal zone B- and B1-cell-specific protein | MZB1 | Inflammation |
| O75888 | Tumor necrosis factor ligand superfamily member 13 | TNFSF13 | Inflammation |
| Q9Y6N7 | Roundabout homolog 1 | ROBO1 | Inflammation |
| O00300 | Tumor necrosis factor receptor superfamily member 11B | TNFRSF11B | Inflammation |
| Q9UJA9 | Ectonucleotide pyrophosphatase/phosphodiesterase family member 5 | ENPP5 | Inflammation |
| Q6GTX8 | Leukocyte-associated immunoglobulin-like receptor 1 | LAIR1 | Inflammation |
| Q5KU26 | Collectin-12 | COLEC12 | Inflammation |
| O00241 | Signal-regulatory protein beta-1 | SIRPB1 | Inflammation |
| Q15389 | Angiopoietin-1 | ANGPT1 | Inflammation |
| P01127 | Platelet-derived growth factor subunit B | PDGFB | Inflammation |
| P46109 | Crk-like protein | CRKL | Inflammation |
| P16422 | Epithelial cell adhesion molecule | EPCAM | Inflammation |
| O43598 | 2'-deoxynucleoside 5'-phosphate N-hydrolase 1 | DNPH1 | Inflammation |
| Q92583 | C-C motif chemokine 17 | CCL17 | Inflammation |
| Q96KG7 | Multiple epidermal growth factor-like domains protein 10 | MEGF10 | Inflammation |
| P24387 | Corticotropin-releasing factor-binding protein | CRHBP | Inflammation |
| P56470 | Galectin-4 | LGALS4 | Inflammation |
| Q9H008 | Phospholysine phosphohistidine inorganic pyrophosphate phosphatase | LHPP | Inflammation |
| O14773 | Tripeptidyl-peptidase 1 | TPP1 | Inflammation |
| Q6UXH1 | Protein disulfide isomerase CRELD2 | CRELD2 | Inflammation |
| Q99895 | Chymotrypsin-C | CTRC | Inflammation |
| Q9NQ76 | Matrix extracellular phosphoglycoprotein | MEPE | Inflammation |
| Q4KMG0 | Cell adhesion molecule-related/down-regulated by oncogenes | CDON | Inflammation |
| Q9UHX3 | Adhesion G protein-coupled receptor E2 | ADGRE2 | Inflammation |
| Q15109 | Advanced glycosylation end product-specific receptor | AGER | Inflammation |
| P27930 | Interleukin-1 receptor type 2 | IL1R2 | Inflammation |
| Q9NQ30 | Endothelial cell-specific molecule 1 | ESM1 | Inflammation |
| Q9HCB6 | Spondin-1 | SPON1 | Inflammation |
| Q9UII2 | ATPase inhibitor, mitochondrial | ATP5IF1 | Inflammation |
| O75563 | Src kinase-associated phosphoprotein 2 | SKAP2 | Inflammation |
| P09341 | Growth-regulated alpha protein | CXCL1 | Inflammation |
| Q16651 | Prostasin | PRSS8 | Inflammation |
| Q03405 | Urokinase plasminogen activator surface receptor | PLAUR | Inflammation |
| O00626 | C-C motif chemokine 22 | CCL22 | Inflammation |
| P51888 | Prolargin | PRELP | Inflammation |
| O00339 | Matrilin-2 | MATN2 | Inflammation |
| P36941 | Tumor necrosis factor receptor superfamily member 3 | LTBR | Inflammation |
| Q16363 | Laminin subunit alpha-4 | LAMA4 | Inflammation |
| P19883 | Follistatin | FST | Inflammation |
| Q9BU40 | Chordin-like protein 1 | CHRDL1 | Inflammation |
| O00175 | C-C motif chemokine 24 | CCL24 | Inflammation |
| Q99538 | Legumain | LGMN | Inflammation |
| Q14118 | Dystroglycan | DAG1 | Inflammation |
| P54317 | Pancreatic lipase-related protein 2 | PNLIPRP2 | Inflammation |
| Q8IYS5 | Osteoclast-associated immunoglobulin-like receptor | OSCAR | Inflammation |
| Q15166 | Serum paraoxonase/lactonase 3 | PON3 | Inflammation |
| P07148 | Fatty acid-binding protein, liver | FABP1 | Inflammation |
| O95866 | Megakaryocyte and platelet inhibitory receptor G6b | MPIG6B | Inflammation |
| P15291 | Beta-1,4-galactosyltransferase 1 | B4GALT1 | Inflammation |
| O00182 | Galectin-9 | LGALS9 | Inflammation |
| O95633 | Follistatin-related protein 3 | FSTL3 | Inflammation |
| Q92956 | Tumor necrosis factor receptor superfamily member 14 | TNFRSF14 | Inflammation |
| Q9BYZ8 | Regenerating islet-derived protein 4 | REG4 | Inflammation |
| Q8TEU8 | WAP, Kazal, immunoglobulin, Kunitz and NTR domain-containing protein 2 | WFIKKN2 | Inflammation |
| O00468 | Agrin | AGRN | Inflammation |
| Q03403 | Trefoil factor 2 | TFF2 | Inflammation |
| P19876 | C-X-C motif chemokine 3 | CXCL3 | Inflammation |
| Q13232 | Nucleoside diphosphate kinase 3 | NME3 | Inflammation |
| Q8N907 | DAN domain family member 5 | DAND5 | Inflammation_II |
| P24530 | Endothelin receptor type B | EDNRB | Inflammation_II |
| Q99665 | Interleukin-12 receptor subunit beta-2 | IL12RB2 | Inflammation_II |
| Q7Z698 | Sprouty-related, EVH1 domain-containing protein 2 | SPRED2 | Inflammation_II |
| Q9NRR2 | Tryptase gamma | TPSG1 | Inflammation_II |
| Q16665 | Hypoxia-inducible factor 1-alpha | HIF1A | Inflammation_II |
| Q9Y4C1 | Lysine-specific demethylase 3A | KDM3A | Inflammation_II |
| Q9H832 | Ubiquitin-conjugating enzyme E2 Z | UBE2Z | Inflammation_II |
| O00206 | Toll-like receptor 4 | TLR4 | Inflammation_II |
| P10070 | Zinc finger protein GLI2 | GLI2 | Inflammation_II |
| Q04609 | Glutamate carboxypeptidase 2 | FOLH1 | Inflammation_II |
| Q9NQI0 | Probable ATP-dependent RNA helicase DDX4 | DDX4 | Inflammation_II |
| Q13190 | Syntaxin-5 | STX5 | Inflammation_II |
| Q6R327 | Rapamycin-insensitive companion of mTOR | RICTOR | Inflammation_II |
| P03372 | Estrogen receptor | ESR1 | Inflammation_II |
| P41273 | Tumor necrosis factor ligand superfamily member 9 | TNFSF9 | Inflammation_II |
| Q9BY41 | Histone deacetylase 8 | HDAC8 | Inflammation_II |
| P43378 | Tyrosine-protein phosphatase non-receptor type 9 | PTPN9 | Inflammation_II |
| Q09472 | Histone acetyltransferase p300 | EP300 | Inflammation_II |
| Q96EB6 | NAD-dependent protein deacetylase sirtuin-1 | SIRT1 | Inflammation_II |
| P24928 | DNA-directed RNA polymerase II subunit RPB1 | POLR2A | Inflammation_II |
| Q9HBE5 | Interleukin-21 receptor | IL21R | Inflammation_II |
| Q92185 | Alpha-N-acetylneuraminide alpha-2,8-sialyltransferase | ST8SIA1 | Inflammation_II |
| P10767 | Fibroblast growth factor 6 | FGF6 | Inflammation_II |
| Q8WX93 | Palladin | PALLD | Inflammation_II |
| Q9UPW0 | Forkhead box protein J3 | FOXJ3 | Inflammation_II |
| P25490 | Transcriptional repressor protein YY1 | YY1 | Inflammation_II |
| Q6UXZ4 | Netrin receptor UNC5D | UNC5D | Inflammation_II |
| P09693 | T-cell surface glycoprotein CD3 gamma chain | CD3G | Inflammation_II |
| Q05329 | Glutamate decarboxylase 2 | GAD2 | Inflammation_II |
| Q6UXM1 | Leucine-rich repeats and immunoglobulin-like domains protein 3 | LRIG3 | Inflammation_II |
| P15531 | Nucleoside diphosphate kinase A | NME1 | Inflammation_II |
| P62834 | Ras-related protein Rap-1A | RAP1A | Inflammation_II |
| P15927 | Replication protein A 32 kDa subunit | RPA2 | Inflammation_II |
| P61328 | Fibroblast growth factor 12 | FGF12 | Inflammation_II |
| P46013 | Proliferation marker protein Ki-67 | MKI67 | Inflammation_II |
| P24864 | G1/S-specific cyclin-E1 | CCNE1 | Inflammation_II |
| P85299 | Proline-rich protein 5 | PRR5 | Inflammation_II |
| P49715 | CCAAT/enhancer-binding protein alpha | CEBPA | Inflammation_II |
| Q9NP95 | Fibroblast growth factor 20 | FGF20 | Inflammation_II |
| P06401 | Progesterone receptor | PGR | Inflammation_II |
| Q6UXL0 | Interleukin-20 receptor subunit beta | IL20RB | Inflammation_II |
| Q9NP85 | Podocin | NPHS2 | Inflammation_II |
| Q01201 | Transcription factor RelB | RELB | Inflammation_II |
| Q9NZS2 | Killer cell lectin-like receptor subfamily F member 1 | KLRF1 | Inflammation_II |
| Q9Y2I7 | 1-phosphatidylinositol 3-phosphate 5-kinase | PIKFYVE | Inflammation_II |
| Q9NWV8 | BRISC and BRCA1-A complex member 1 | BABAM1 | Inflammation_II |
| O00401 | Neural Wiskott-Aldrich syndrome protein | WASL | Inflammation_II |
| Q04837 | Single-stranded DNA-binding protein, mitochondrial | SSBP1 | Inflammation_II |
| O75365 | Protein tyrosine phosphatase type IVA 3 | PTP4A3 | Inflammation_II |
| Q12888 | TP53-binding protein 1 | TP53BP1 | Inflammation_II |
| O75173 | A disintegrin and metalloproteinase with thrombospondin motifs 4 | ADAMTS4 | Inflammation_II |
| Q9UHA7 | Interleukin-36 alpha | IL36A | Inflammation_II |
| Q96EP0 | E3 ubiquitin-protein ligase RNF31 | RNF31 | Inflammation_II |
| O95429 | BAG family molecular chaperone regulator 4 | BAG4 | Inflammation_II |
| Q8NI17 | Interleukin-31 receptor subunit alpha | IL31RA | Inflammation_II |
| Q9NS62 | Thrombospondin type-1 domain-containing protein 1 | THSD1 | Inflammation_II |
| Q9NY59 | Sphingomyelin phosphodiesterase 3 | SMPD3 | Inflammation_II |
| Q92574 | Hamartin | TSC1 | Inflammation_II |
| Q13114-2 | TNF receptor-associated factor 3, Isoform 2 | TRAF3 | Inflammation_II |
| P54274 | Telomeric repeat-binding factor 1 | TERF1 | Inflammation_II |
| Q96MM7 | Heparan-sulfate 6-O-sulfotransferase 2 | HS6ST2 | Inflammation_II |
| P35219 | Carbonic anhydrase-related protein | CA8 | Inflammation_II |
| Q9GZN4 | Brain-specific serine protease 4 | PRSS22 | Inflammation_II |
| Q9NQ66 | 1-phosphatidylinositol 4,5-bisphosphate phosphodiesterase beta-1 | PLCB1 | Inflammation_II |
| P16671 | Platelet glycoprotein 4 | CD36 | Inflammation_II |
| Q8NBK3 | Formylglycine-generating enzyme | SUMF1 | Inflammation_II |
| Q6EBC2 | Interleukin-31 | IL31 | Inflammation_II |
| Q8NHP1 | Aflatoxin B1 aldehyde reductase member 4 | AKR7L | Inflammation_II |
| Q9Y3D3 | 28S ribosomal protein S16, mitochondrial | MRPS16 | Inflammation_II |
| P98170 | E3 ubiquitin-protein ligase XIAP | XIAP | Inflammation_II |
| O43184 | Growth/differentiation factor 15 | ADAM12 | Inflammation_II |
| P48730 | Casein kinase I isoform delta | CSNK1D | Inflammation_II |
| O43583 | Density-regulated protein | DENR | Inflammation_II |
| O00148 | ATP-dependent RNA helicase DDX39A | DDX39A | Inflammation_II |
| P36551 | Oxygen-dependent coproporphyrinogen-III oxidase, mitochondrial | CPOX | Inflammation_II |
| Q96D71 | RalBP1-associated Eps domain-containing protein 1 | REPS1 | Inflammation_II |
| P09923 | Intestinal-type alkaline phosphatase | ALPI | Inflammation_II |
| Q5TBC7 | Bcl-2-like protein 15 | BCL2L15 | Inflammation_II |
| Q9UHN6 | Cell surface hyaluronidase | CEMIP2 | Inflammation_II |
| Q6B9Z1 | Insulin growth factor-like family member 4 | IGFL4 | Inflammation_II |
| Q8IX19 | Mast cell-expressed membrane protein 1 | MCEMP1 | Inflammation_II |
| P36897 | TGF-beta receptor type-1 | TGFBR1 | Inflammation_II |
| Q14511 | Enhancer of filamentation 1 | NEDD9 | Inflammation_II |
| Q8WVV4 | Protein POF1B | POF1B | Inflammation_II |
| Q9UIK4 | Death-associated protein kinase 2 | DAPK2 | Inflammation_II |
| Q8IYW5 | E3 ubiquitin-protein ligase RNF168 | RNF168 | Inflammation_II |
| Q93062 | RNA-binding protein with multiple splicing | RBPMS | Inflammation_II |
| Q15697 | Zinc finger protein 174 | ZNF174 | Inflammation_II |
| P17050 | Alpha-N-acetylgalactosaminidase | NAGA | Inflammation_II |
| Q96PX8 | SLIT and NTRK-like protein 1 | SLITRK1 | Inflammation_II |
| P56645 | Period circadian protein homolog 3 | PER3 | Inflammation_II |
| Q9H0U9 | Testis-specific Y-encoded-like protein 1 | TSPYL1 | Inflammation_II |
| P26436 | Acrosomal protein SP-10 | ACRV1 | Inflammation_II |
| Q5JS54 | Proteasome assembly chaperone 4 | PSMG4 | Inflammation_II |
| Q7Z5L3 | Complement C1q-like protein 2 | C1QL2 | Inflammation_II |
| Q15465 | Sonic hedgehog protein | SHH | Inflammation_II |
| P17643 | 5,6-dihydroxyindole-2-carboxylic acid oxidase | TYRP1 | Inflammation_II |
| Q8IV38 | Ankyrin repeat and MYND domain-containing protein 2 | ANKMY2 | Inflammation_II |
| O60447 | Ecotropic viral integration site 5 protein homolog | EVI5 | Inflammation_II |
| Q16718 | NADH dehydrogenase [ubiquinone] 1 alpha subcomplex subunit 5 | NDUFA5 | Inflammation_II |
| Q96PU5 | E3 ubiquitin-protein ligase NEDD4-like | NEDD4L | Inflammation_II |
| O94916 | Nuclear factor of activated T-cells 5 | NFAT5 | Inflammation_II |
| O95835 | Serine/threonine-protein kinase LATS1 | LATS1 | Inflammation_II |
| P24666 | Low molecular weight phosphotyrosine protein phosphatase | ACP1 | Inflammation_II |
| O60238 | BCL2/adenovirus E1B 19 kDa protein-interacting protein 3-like | BNIP3L | Inflammation_II |
| Q9H171 | Z-DNA-binding protein 1 | ZBP1 | Inflammation_II |
| Q99062 | Granulocyte colony-stimulating factor receptor | CSF3R | Inflammation_II |
| Q9UHI8 | A disintegrin and metalloproteinase with thrombospondin motifs 1 | ADAMTS1 | Inflammation_II |
| Q99584 | Protein S100-A13 | S100A13 | Inflammation_II |
| P55211 | Caspase-9 | CASP9 | Inflammation_II |
| Q9H3T2 | Semaphorin-6C | SEMA6C | Inflammation_II |
| Q15399 | Toll-like receptor 1 | TLR1 | Inflammation_II |
| Q9Y2X7 | ARF GTPase-activating protein GIT1 | GIT1 | Inflammation_II |
| P81534 | Beta-defensin 103 | DEFB103A_DEFB103B | Inflammation_II |
| O43320 | Fibroblast growth factor 16 | FGF16 | Inflammation_II |
| P52630 | Signal transducer and activator of transcription 2 | STAT2 | Inflammation_II |
| P20701 | Integrin alpha-L | ITGAL | Inflammation_II |
| Q86UE4 | Protein LYRIC | MTDH | Inflammation_II |
| Q96PL5 | Erythroid membrane-associated protein | ERMAP | Inflammation_II |
| O60500 | Nephrin | NPHS1 | Inflammation_II |
| Q8WV28 | B-cell linker protein | BLNK | Inflammation_II |
| P48546 | Gastric inhibitory polypeptide receptor | GIPR | Inflammation_II |
| P09564 | T-cell antigen CD7 | CD7 | Inflammation_II |
| P11487 | Fibroblast growth factor 3 | FGF3 | Inflammation_II |
| Q15223 | Nectin-1 | NECTIN1 | Inflammation_II |
| P23743 | Diacylglycerol kinase alpha | DGKA | Inflammation_II |
| P29536 | Leiomodin-1 | LMOD1 | Inflammation_II |
| O75688 | Protein phosphatase 1B | PPM1B | Inflammation_II |
| Q16643 | Drebrin | DBN1 | Inflammation_II |
| Q5T2W1 | Na(+)/H(+) exchange regulatory cofactor NHE-RF3 | PDZK1 | Inflammation_II |
| Q9NZH8 | Interleukin-36 gamma | IL36G | Inflammation_II |
| Q5QGZ9 | C-type lectin domain family 12 member A | CLEC12A | Inflammation_II |
| P32927 | Cytokine receptor common subunit beta | CSF2RB | Inflammation_II |
| P49765 | Vascular endothelial growth factor B | VEGFB | Inflammation_II |
| Q9BV40 | Vesicle-associated membrane protein 8 | VAMP8 | Inflammation_II |
| Q15762 | CD226 antigen | CD226 | Inflammation_II |
| P06730 | Eukaryotic translation initiation factor 4E | EIF4E | Inflammation_II |
| Q14160 | Protein scribble homolog | SCRIB | Inflammation_II |
| Q02880 | DNA topoisomerase 2-beta | TOP2B | Inflammation_II |
| O15400 | Syntaxin-7 | STX7 | Inflammation_II |
| Q9H7Z7 | Prostaglandin E synthase 2 | PTGES2 | Inflammation_II |
| P14902 | Indoleamine 2,3-dioxygenase 1 | IDO1 | Inflammation_II |
| O60437 | Periplakin | PPL | Inflammation_II |
| O95157 | Neurexophilin-3 | NXPH3 | Inflammation_II |
| P06213 | Insulin receptor | INSR | Inflammation_II |
| P11234 | Ras-related protein Ral-B | RALB | Inflammation_II |
| P49757 | Protein numb homolog | NUMB | Inflammation_II |
| Q8N556 | Actin filament-associated protein 1 | AFAP1 | Inflammation_II |
| P17301 | Integrin alpha-2 | ITGA2 | Inflammation_II |
| Q02952 | A-kinase anchor protein 12 | AKAP12 | Inflammation_II |
| Q4VCS5 | Angiomotin | AMOT | Inflammation_II |
| P30101 | Protein disulfide-isomerase A3 | PDIA3 | Inflammation_II |
| P10912 | Growth hormone receptor | GHR | Inflammation_II |
| O95393 | Bone morphogenetic protein 10 | BMP10 | Inflammation_II |
| Q9UGN4 | CMRF35-like molecule 8 | CD300A | Inflammation_II |
| Q96QR1 | Secretoglobin family 3A member 1 | SCGB3A1 | Inflammation_II |
| P30040 | Endoplasmic reticulum resident protein 29 | ERP29 | Inflammation_II |
| Q9ULI3 | Protein HEG homolog 1 | HEG1 | Inflammation_II |
| Q8TEA8 | D-aminoacyl-tRNA deacylase 1 | DTD1 | Inflammation_II |
| P01037 | Cystatin-SN | CST1 | Inflammation_II |
| Q9H7Y0 | Divergent protein kinase domain 2B | DIPK2B | Inflammation_II |
| Q9BRK3 | Matrix remodeling-associated protein 8 | MXRA8 | Inflammation_II |
| O76074 | cGMP-specific 3',5'-cyclic phosphodiesterase | PDE5A | Inflammation_II |
| P22303 | Acetylcholinesterase | ACHE | Inflammation_II |
| Q9BX67 | Junctional adhesion molecule C | JAM3 | Inflammation_II |
| P20908 | Collagen alpha-1(V) chain | COL5A1 | Inflammation_II |
| Q6QNK2 | Adhesion G-protein coupled receptor D1 | ADGRD1 | Inflammation_II |
| Q02223 | Tumor necrosis factor receptor superfamily member 17 | TNFRSF17 | Inflammation_II |
| Q8NEU8 | DCC-interacting protein 13-beta | APPL2 | Inflammation_II |
| Q9BUH6 | Protein PAXX | PAXX | Inflammation_II |
| Q8NDA2 | Hemicentin-2 | HMCN2 | Inflammation_II |
| Q9HCU4 | Cadherin EGF LAG seven-pass G-type receptor 2 | CELSR2 | Inflammation_II |
| P06280 | Alpha-galactosidase A | GLA | Inflammation_II |
| Q9BUN1 | Protein MENT | MENT | Inflammation_II |
| Q9H939 | Proline-serine-threonine phosphatase-interacting protein 2 | PSTPIP2 | Inflammation_II |
| Q8TDQ7 | Glucosamine-6-phosphate isomerase 2 | GNPDA2 | Inflammation_II |
| P55083 | Microfibril-associated glycoprotein 4 | MFAP4 | Inflammation_II |
| Q9NS98 | Semaphorin-3G | SEMA3G | Inflammation_II |
| P04406 | Glyceraldehyde-3-phosphate dehydrogenase | GAPDH | Inflammation_II |
| P04090 | Prorelaxin H2 | RLN2 | Inflammation_II |
| Q5TDH0 | Protein DDI1 homolog 2 | DDI2 | Inflammation_II |
| P06132 | Uroporphyrinogen decarboxylase | UROD | Inflammation_II |
| Q99574 | Neuroserpin | SERPINI1 | Inflammation_II |
| P27348 | 14-3-3 protein theta | YWHAQ | Inflammation_II |
| Q9UI42 | Carboxypeptidase A4 | CPA4 | Inflammation_II |
| Q16378 | Proline-rich protein 4 | PRR4 | Inflammation_II |
| Q6P5S2 | Protein LEG1 homolog | LEG1 | Inflammation_II |
| O43399 | Tumor protein D54 | TPD52L2 | Inflammation_II |
| P61457 | Pterin-4-alpha-carbinolamine dehydratase | PCBD1 | Inflammation_II |
| P40197 | Platelet glycoprotein V | GP5 | Inflammation_II |
| P00325 | All-trans-retinol dehydrogenase | ADH1B | Inflammation_II |
| O75190 | DnaJ homolog subfamily B member 6 | DNAJB6 | Inflammation_II |
| Q5JS37 | NHL repeat-containing protein 3 | NHLRC3 | Inflammation_II |
| Q8N8U9 | BMP-binding endothelial regulator protein | BMPER | Inflammation_II |
| O75347 | Tubulin-specific chaperone A | TBCA | Inflammation_II |
| P04083 | Annexin A1 | ANXA1 | Inflammation_II |
| Q9BXJ0 | Complement C1q tumor necrosis factor-related protein 5 | C1QTNF5 | Inflammation_II |
| Q96HD1 | Protein disulfide isomerase CRELD1 | CRELD1 | Inflammation_II |
| P0C862 | Complement C1q and tumor necrosis factor-related protein 9A | C1QTNF9 | Inflammation_II |
| Q13976 | cGMP-dependent protein kinase 1 | PRKG1 | Inflammation_II |
| P54764 | Ephrin type-A receptor 4 | EPHA4 | Inflammation_II |
| Q9P2T1 | GMP reductase 2 | GMPR2 | Inflammation_II |
| P50502 | Hsc70-interacting protein | ST13 | Inflammation_II |
| P09529 | Inhibin beta B chain | INHBB | Inflammation_II |
| A2VDF0 | Fucose mutarotase | FUOM | Inflammation_II |
| P20155 | Serine protease inhibitor Kazal-type 2 | SPINK2 | Inflammation_II |
| P14091 | Cathepsin E | CTSE | Inflammation_II |
| Q6P589 | Tumor necrosis factor alpha-induced protein 8-like protein 2 | TNFAIP8L2 | Inflammation_II |
| O14745 | Na(+)/H(+) exchange regulatory cofactor NHE-RF1 | SLC9A3R1 | Inflammation_II |
| P30047 | GTP cyclohydrolase 1 feedback regulatory protein | GCHFR | Inflammation_II |
| Q53FA7 | Quinone oxidoreductase PIG3 | TP53I3 | Inflammation_II |
| Q5VTT5 | Myomesin-3 | MYOM3 | Inflammation_II |
| Q8IUZ5 | 5-phosphohydroxy-L-lysine phospho-lyase | PHYKPL | Inflammation_II |
| P52758 | 2-iminobutanoate/2-iminopropanoate deaminase | RIDA | Inflammation_II |
| Q96A49 | Synapse-associated protein 1 | SYAP1 | Inflammation_II |
| Q9NQ48 | Leucine zipper transcription factor-like protein 1 | LZTFL1 | Inflammation_II |
| O96007 | Molybdopterin synthase catalytic subunit | MOCS2 | Inflammation_II |
| Q8NHV1 | GTPase IMAP family member 7 | GIMAP7 | Inflammation_II |
| Q96AJ9 | Vesicle transport through interaction with t-SNAREs homolog 1A | VTI1A | Inflammation_II |
| P21854 | B-cell differentiation antigen CD72 | CD72 | Inflammation_II |
| P07311 | Acylphosphatase-1 | ACYP1 | Inflammation_II |
| Q9UK23 | N-acetylglucosamine-1-phosphodiester alpha-N-acetylglucosaminidase | NAGPA | Inflammation_II |
| Q9NRS6 | Sorting nexin-15 | SNX15 | Inflammation_II |
| P37840 | Alpha-synuclein | SNCA | Inflammation_II |
| P22455 | Fibroblast growth factor receptor 4 | FGFR4 | Inflammation_II |
| Q15276 | Rab GTPase-binding effector protein 1 | RABEP1 | Inflammation_II |
| Q6FHJ7 | Secreted frizzled-related protein 4 | SFRP4 | Inflammation_II |
| Q9HB71 | Calcyclin-binding protein | CACYBP | Inflammation_II |
| O60279 | Sushi domain-containing protein 5 | SUSD5 | Inflammation_II |
| A6NC86 | phospholipase A2 inhibitor and Ly6/PLAUR domain-containing protein | PINLYP | Inflammation_II |
| P49862 | Kallikrein-7 | KLK7 | Inflammation_II |
| Q92619 | Rho GTPase-activating protein 45 | ARHGAP45 | Inflammation_II |
| Q04323 | UBX domain-containing protein 1 | UBXN1 | Inflammation_II |
| P35611 | Alpha-adducin | ADD1 | Inflammation_II |
| Q8N0X7 | Spartin | SPART | Inflammation_II |
| P32321 | Deoxycytidylate deaminase | DCTD | Inflammation_II |
| P56192 | Methionine--tRNA ligase, cytoplasmic | MARS1 | Inflammation_II |
| Q6GMV3 | Putative peptidyl-tRNA hydrolase PTRHD1 | PTRHD1 | Inflammation_II |
| O15335 | Chondroadherin | CHAD | Inflammation_II |
| P25686 | DnaJ homolog subfamily B member 2 | DNAJB2 | Inflammation_II |
| Q58EX2 | Protein sidekick-2 | SDK2 | Inflammation_II |
| Q86SX6 | Glutaredoxin-related protein 5, mitochondrial | GLRX5 | Inflammation_II |
| Q9Y4D1 | Disheveled-associated activator of morphogenesis 1 | DAAM1 | Inflammation_II |
| Q6UX06 | Olfactomedin-4 | OLFM4 | Inflammation_II |
| P01210 | Proenkephalin-A | PENK | Inflammation_II |
| P0DML2 | Chorionic somatomammotropin hormone 1 | CSH1 | Inflammation_II |
| P32119 | Peroxiredoxin-2 | PRDX2 | Inflammation_II |
| P40925 | Malate dehydrogenase, cytoplasmic | MDH1 | Inflammation_II |
| P10599 | Thioredoxin | TXN | Inflammation_II |
| P0DN86 | Choriogonadotropin subunit beta 3 | CGB3_CGB5_CGB8 | Inflammation_II |
| P07998 | Ribonuclease pancreatic | RNASE1 | Inflammation_II |
| P04114 | Apolipoprotein B-100 | APOB | Inflammation_II |
| Q92686 | Neurogranin | NRGN | Inflammation_II |
| P16442 | Histo-blood group ABO system transferase | ABO | Inflammation_II |
| Q6UXB8 | Peptidase inhibitor 16 | PI16 | Inflammation_II |
| P04180 | Phosphatidylcholine-sterol acyltransferase | LCAT | Inflammation_II |
| Q0ZGT2 | Nexilin | NEXN | Inflammation_II |
| P07307 | Asialoglycoprotein receptor 2 | ASGR2 | Inflammation_II |
| P06744 | Glucose-6-phosphate isomerase | GPI | Inflammation_II |
| P07333 | Macrophage colony-stimulating factor 1 receptor | CSF1R | Inflammation_II |
| Q86YW5 | Trem-like transcript 1 protein | TREML1 | Inflammation_II |
| P08294 | Extracellular superoxide dismutase | SOD3 | Inflammation_II |
| P00742 | Coagulation factor X | F10 | Inflammation_II |
| P04278 | Sex hormone-binding globulin | SHBG | Inflammation_II |
| P02652 | Apolipoprotein A-II | APOA2 | Inflammation_II |
| P20061 | Transcobalamin-1 | TCN1 | Inflammation_II |
| P12955 | Xaa-Pro dipeptidase | PEPD | Inflammation_II |
| Q93091 | Ribonuclease K6 | RNASE6 | Inflammation_II |
| O00602 | Ficolin-1 | FCN1 | Inflammation_II |
| O43493 | Trans-Golgi network integral membrane protein 2 | TGOLN2 | Inflammation_II |
| P50552 | Vasodilator-stimulated phosphoprotein | VASP | Inflammation_II |
| P16233 | Pancreatic triacylglycerol lipase | PNLIP | Inflammation_II |
| P22897 | Macrophage mannose receptor 1 | MRC1 | Inflammation_II |
| P12821 | Angiotensin-converting enzyme | ACE | Inflammation_II |
| P17927 | Complement receptor type 1 | CR1 | Inflammation_II |
| P02748 | Complement component C9 | C9 | Inflammation_II |
| P0DJD7 | Pepsin A-4 | PGA4 | Inflammation_II |
| P00390 | Glutathione reductase, mitochondrial | GSR | Inflammation_II |
| Q13790 | Apolipoprotein F | APOF | Inflammation_II |
| Q08830 | Fibrinogen-like protein 1 | FGL1 | Inflammation_II |
| P09172 | Dopamine beta-hydroxylase | DBH | Inflammation_II |
| P27169 | Serum paraoxonase/arylesterase 1 | PON1 | Inflammation_II |
| P34096 | Ribonuclease 4 | RNASE4 | Inflammation_II |
| P02765 | Alpha-2-HS-glycoprotein | AHSG | Inflammation_II |
| P0DUB6_P0DTE7_P0DTE8 | Alpha-amylase 1A_Alpha-amylase 1B_Alpha-amylase 1C | AMY1A_AMY1B_AMY1C | Inflammation_II |
| O14791 | Apolipoprotein L1 | APOL1 | Inflammation_II |
| P01019 | Angiotensinogen | AGT | Inflammation_II |
| Q01459 | Di-N-acetylchitobiase | CTBS | Inflammation_II |
| O95497 | Pantetheinase | VNN1 | Inflammation_II |
| P04040 | Catalase | CAT | Inflammation_II |
| P26927 | Hepatocyte growth factor-like protein | MST1 | Inflammation_II |
| P06396 | Gelsolin | GSN | Inflammation_II |
| P54108 | Cysteine-rich secretory protein 3 | CRISP3 | Inflammation_II |
| Q9BXR6 | Complement factor H-related protein 5 | CFHR5 | Inflammation_II |
| P06276 | Cholinesterase | BCHE | Inflammation_II |
| Q04756 | Hepatocyte growth factor activator | HGFAC | Inflammation_II |
| P07358 | Complement component C8 beta chain | C8B | Inflammation_II |
| P36980 | Complement factor H-related protein 2 | CFHR2 | Inflammation_II |
| Q9NZP8 | Complement C1r subcomponent-like protein | C1RL | Inflammation_II |
| P19827 | Inter-alpha-trypsin inhibitor heavy chain H1 | ITIH1 | Inflammation_II |
| P00734 | Prothrombin | F2 | Inflammation_II |
| P05543 | Thyroxine-binding globulin | SERPINA7 | Inflammation_II |
| P61769 | Beta-2-microglobulin | B2M | Inflammation_II |
| P35542 | Serum amyloid A-4 protein | SAA4 | Inflammation_II |
| P02649 | Apolipoprotein E | APOE | Inflammation_II |
| Q9Y5Y7 | Lymphatic vessel endothelial hyaluronic acid receptor 1 | LYVE1 | Inflammation_II |
| O00391 | Sulfhydryl oxidase 1 | QSOX1 | Inflammation_II |
| P20742 | Pregnancy zone protein | PZP | Inflammation_II |
| P09871 | Complement C1s subcomponent | C1S | Inflammation_II |
| P10909 | Clusterin | CLU | Inflammation_II |
| P06727 | Apolipoprotein A-IV | APOA4 | Inflammation_II |
| Q96IY4 | Carboxypeptidase B2 | CPB2 | Inflammation_II |
| O75882-2 | Attractin, Isoform 2 | ATRN | Inflammation_II |
| Q16610 | Extracellular matrix protein 1 | ECM1 | Inflammation_II |
| P07225 | Vitamin K-dependent protein S | PROS1 | Inflammation_II |
| P00746 | Complement factor D | CFD | Inflammation_II |
| P00748 | Coagulation factor XII | F12 | Inflammation_II |
| P0DOY2 | Immunoglobulin lambda constant 2 | IGLC2 | Inflammation_II |
| P43652 | Afamin | AFM | Inflammation_II |
| Q96PD5 | N-acetylmuramoyl-L-alanine amidase | PGLYRP2 | Inflammation_II |
| P08185 | Corticosteroid-binding globulin | SERPINA6 | Inflammation_II |
| P01031 | Complement C5 | C5 | Inflammation_II |
| P49908 | Selenoprotein P | SELENOP | Inflammation_II |
| P05090 | Apolipoprotein D | APOD | Inflammation_II |
| P08519 | Apolipoprotein(a) | LPA | Inflammation_II |
| Q15848 | Adiponectin | ADIPOQ | Inflammation_II |
| P02654 | Apolipoprotein C-I | APOC1 | Inflammation_II |
| P01009 | Alpha-1-antitrypsin | SERPINA1 | Inflammation_II |
| P02750 | Leucine-rich alpha-2-glycoprotein | LRG1 | Inflammation_II |
| P14151 | L-selectin | SELL | Inflammation_II |
| P00736 | Complement C1r subcomponent | C1R | Inflammation_II |
| P43251 | Biotinidase | BTD | Inflammation_II |
| P00751 | Complement factor B | CFB | Inflammation_II |
| P05452 | Tetranectin | CLEC3B | Inflammation_II |
| P36955 | Pigment epithelium-derived factor | SERPINF1 | Inflammation_II |
| Q92496 | Complement factor H-related protein 4 | CFHR4 | Inflammation_II |
| P11226 | Mannose-binding protein C | MBL2 | Inflammation_II |
| P03952 | Plasma kallikrein | KLKB1 | Inflammation_II |
| P02775 | Platelet basic protein | PPBP | Inflammation_II |
| Q14624 | Inter-alpha-trypsin inhibitor heavy chain H4 | ITIH4 | Inflammation_II |
| P05154 | Plasma serine protease inhibitor | SERPINA5 | Inflammation_II |
| P02776 | Platelet factor 4 | PF4 | Inflammation_II |
| Q08380 | Galectin-3-binding protein | LGALS3BP | Inflammation_II |
| P10643 | Complement component C7 | C7 | Inflammation_II |
| P05546 | Heparin cofactor 2 | SERPIND1 | Inflammation_II |
| P02763 | Alpha-1-acid glycoprotein 1 | ORM1 | Inflammation_II |
| P02647 | Apolipoprotein A-I | APOA1 | Inflammation_II |
| P04196 | Histidine-rich glycoprotein | HRG | Inflammation_II |
| P04217 | Alpha-1B-glycoprotein | A1BG | Inflammation_II |
| P05156 | Complement factor I | CFI | Inflammation_II |
| P03951 | Coagulation factor XI | F11 | Inflammation_II |
| P29622 | Kallistatin | SERPINA4 | Inflammation_II |
| O43866 | CD5 antigen-like | CD5L | Inflammation_II |
| P01024 | Complement C3 | C3 | Inflammation_II |
| P05155 | Plasma protease C1 inhibitor | SERPING1 | Inflammation_II |
| P02743 | Serum amyloid P-component | APCS | Inflammation_II |
| P08697 | Alpha-2-antiplasmin | SERPINF2 | Inflammation_II |
| P02774 | Vitamin D-binding protein | GC | Inflammation_II |
| P05160 | Coagulation factor XIII B chain | F13B | Inflammation_II |
| P02766 | Transthyretin | TTR | Inflammation_II |
| P02787 | Serotransferrin | TF | Inflammation_II |
| P27918 | Properdin | CFP | Inflammation_II |
| P00747 | Plasminogen | PLG | Inflammation_II |
| P01011 | Alpha-1-antichymotrypsin | SERPINA3 | Inflammation_II |
| P02751 | Fibronectin | FN1 | Inflammation_II |
| P02671 | Fibrinogen alpha chain | FGA | Inflammation_II |
| P01008 | Antithrombin-III | SERPINC1 | Inflammation_II |
| P08603 | Complement factor H | CFH | Inflammation_II |

**Caption:** Protein is characterised with UniProt ID, Protein name, Gene name and the explore panel to which they belong.

**Supplementary table 3:** Linear mixed-effects models to assess associations of proteins at ~62 years old with BMI changes during adulthood (24 to 62 years old).

| **Protein description** | **Protein ID** | **Estimate** | **SE** | **p value** | | Protein  panel |
| --- | --- | --- | --- | --- | --- | --- |
|  |  |  |  | **Nominal** | **Bonferroni** |  |
| Ectonucleotide pyrophosphatase/phosphodiesterase family member 7 | Q6UWV6 | 0.024 | 0.004 | 8.42e-08 | 0.000104 | Inflammation |
| Coagulation factor IX | P00740 | 0.108 | 0.02 | 8.75e-08 | 0.000108 | Cardiometabolic |
| Thrombospondin-4 | P35443 | 0.046 | 0.009 | 1.12e-07 | 0.000138 | Cardiometabolic |
| Lysosomal Pro-X carboxypeptidase | P42785 | 0.072 | 0.013 | 1.32e-07 | 0.000163 | Cardiometabolic |
| Platelet glycoprotein 4 | P16671 | 0.073 | 0.014 | 1.41e-07 | 0.000174 | Inflammation |
| C-C motif chemokine 16 | O15467 | 0.051 | 0.01 | 1.61e-07 | 0.000198 | Cardiometabolic |
| Complement factor D | P00746 | 0.108 | 0.02 | 1.84e-07 | 0.000227 | Inflammation |
| Angiotensin-converting enzyme 2 | Q9BYF1 | 0.039 | 0.007 | 2.29e-07 | 0.000282 | Cardiometabolic |
| SLIT and NTRK-like protein 1 | Q96PX8 | -0.069 | 0.013 | 2.57e-07 | 0.000317 | Inflammation |
| Angiomotin | Q4VCS5 | 0.047 | 0.009 | 2.72e-07 | 0.000335 | Inflammation |
| Macrophage colony-stimulating factor 1 | P09603 | 0.091 | 0.017 | 2.82e-07 | 0.000347 | Inflammation |
| Fructose-1,6-bisphosphatase 1 | P09467 | 0.027 | 0.005 | 2.96e-07 | 0.000365 | Cardiometabolic |
| Neurexophilin-3 | O95157 | -0.067 | 0.013 | 3.64e-07 | 0.000448 | Inflammation |
| BPI fold-containing family B member 1 | Q8TDL5 | -0.033 | 0.006 | 4.09e-07 | 0.000504 | Cardiometabolic |
| N-acetylneuraminate lyase | Q9BXD5 | 0.047 | 0.009 | 4.12e-07 | 0.000508 | Cardiometabolic |
| Collectin-12 | Q5KU26 | 0.095 | 0.018 | 4.19e-07 | 0.000516 | Inflammation |
| Dickkopf-related protein 3 | Q9UBP4 | -0.067 | 0.013 | 4.31e-07 | 0.000532 | Cardiometabolic |
| Macrophage colony-stimulating factor 1 receptor | P07333 | 0.047 | 0.009 | 4.64e-07 | 0.000572 | Inflammation |
| Glycerophosphocholine cholinephosphodiesterase ENPP6 | Q6UWR7 | -0.057 | 0.011 | 4.77e-07 | 0.000587 | Cardiometabolic |
| Leptin receptor | P48357 | -0.08 | 0.016 | 5.66e-07 | 0.000698 | Cardiometabolic |
| Somatotropin | P01241 | -0.012 | 0.002 | 5.91e-07 | 0.000728 | Cardiometabolic |
| Tissue-type plasminogen activator | P00750 | 0.03 | 0.006 | 6.45e-07 | 0.000795 | Cardiometabolic |
| Gamma-glutamyl hydrolase | Q92820 | 0.057 | 0.011 | 6.64e-07 | 0.000818 | Cardiometabolic |
| Phospholipid transfer protein | P55058 | -0.063 | 0.012 | 7.34e-07 | 0.000904 | Cardiometabolic |
| Dermatopontin | Q07507 | 0.08 | 0.016 | 7.55e-07 | 0.00093 | Cardiometabolic |
| Alpha-amylase 1A_Alpha-amylase 1B_Alpha-amylase 1C | P0DUB6_P0DTE7_P0DTE8 | -0.047 | 0.009 | 8.46e-07 | 0.00104 | Inflammation |
| Cytidine deaminase | P32320 | 0.054 | 0.011 | 8.61e-07 | 0.00106 | Cardiometabolic |
| Alpha-amylase 2B | P19961 | -0.044 | 0.009 | 1.07e-06 | 0.00131 | Cardiometabolic |
| Pantetheinase | O95497 | 0.028 | 0.006 | 1.21e-06 | 0.00149 | Inflammation |
| Pancreatic alpha-amylase | P04746 | -0.044 | 0.009 | 1.26e-06 | 0.00155 | Cardiometabolic |
| Uromodulin | P07911 | -0.039 | 0.008 | 1.49e-06 | 0.00183 | Cardiometabolic |
| BPI fold-containing family A member 2 | Q96DR5 | -0.02 | 0.004 | 1.82e-06 | 0.00225 | Cardiometabolic |
| B-cell differentiation antigen CD72 | P21854 | 0.049 | 0.01 | 2.63e-06 | 0.00324 | Inflammation |
| Egl nine homolog 1 | Q9GZT9 | 0.042 | 0.009 | 3.32e-06 | 0.00409 | Inflammation |
| Secretoglobin family 3A member 2 | Q96PL1 | -0.024 | 0.005 | 4.25e-06 | 0.00524 | Inflammation |
| Ficolin-2 | Q15485 | 0.049 | 0.01 | 4.63e-06 | 0.00571 | Cardiometabolic |
| Complement factor B | P00751 | 0.065 | 0.014 | 4.8e-06 | 0.00591 | Inflammation |
| C-C motif chemokine 27 | Q9Y4X3 | -0.041 | 0.009 | 4.86e-06 | 0.00598 | Cardiometabolic |
| Protein tyrosine phosphatase type IVA 3 | O75365 | 0.036 | 0.008 | 5.17e-06 | 0.00637 | Inflammation |
| Thimet oligopeptidase | P52888 | 0.064 | 0.014 | 5.39e-06 | 0.00664 | Cardiometabolic |
| D-dopachrome decarboxylase | P30046 | 0.046 | 0.01 | 5.89e-06 | 0.00726 | Cardiometabolic |
| Mevalonate kinase | Q03426 | 0.023 | 0.005 | 6.25e-06 | 0.0077 | Inflammation |
| Fatty acid-binding protein, heart | P05413 | 0.041 | 0.009 | 6.6e-06 | 0.00813 | Cardiometabolic |
| Collagen triple helix repeat-containing protein 1 | Q96CG8 | 0.061 | 0.013 | 7.06e-06 | 0.0087 | Cardiometabolic |
| Agrin | O00468 | 0.062 | 0.014 | 9.63e-06 | 0.0119 | Inflammation |
| Plasminogen activator inhibitor 1 | P05121 | 0.031 | 0.007 | 1e-05 | 0.0124 | Cardiometabolic |
| Tectonic-3 | Q6NUS6 | 0.056 | 0.013 | 1.02e-05 | 0.0126 | Cardiometabolic |
| Ganglioside GM2 activator | P17900 | 0.066 | 0.015 | 1.06e-05 | 0.0131 | Cardiometabolic |
| Delta and Notch-like epidermal growth factor-related receptor | Q8NFT8 | -0.073 | 0.016 | 1.11e-05 | 0.0137 | Inflammation |
| Cell surface hyaluronidase | Q9UHN6 | 0.059 | 0.013 | 1.17e-05 | 0.0144 | Inflammation |
| Receptor-type tyrosine-protein phosphatase zeta | P23471 | -0.053 | 0.012 | 1.18e-05 | 0.0145 | Cardiometabolic |
| Kynureninase | Q16719 | 0.043 | 0.01 | 1.2e-05 | 0.0148 | Inflammation |
| Interleukin-12 subunit beta | P29460 | 0.033 | 0.007 | 1.36e-05 | 0.0168 | Inflammation |
| Insulin-like growth factor-binding protein-like 1 | Q8WX77 | 0.062 | 0.014 | 1.47e-05 | 0.0181 | Cardiometabolic |
| Aflatoxin B1 aldehyde reductase member 4 | Q8NHP1 | 0.02 | 0.005 | 1.62e-05 | 0.02 | Inflammation |
| Serine--pyruvate aminotransferase | P21549 | 0.025 | 0.006 | 1.67e-05 | 0.0206 | Cardiometabolic |
| Stabilin-2 | Q8WWQ8 | 0.067 | 0.015 | 1.78e-05 | 0.0219 | Cardiometabolic |
| Multiple epidermal growth factor-like domains protein 9 | Q9H1U4 | 0.091 | 0.021 | 1.95e-05 | 0.024 | Cardiometabolic |
| Corticosteroid 11-beta-dehydrogenase isozyme 1 | P28845 | -0.055 | 0.013 | 2.01e-05 | 0.0247 | Inflammation |
| Leukocyte immunoglobulin-like receptor subfamily A member 5 | A6NI73 | 0.055 | 0.013 | 2.7e-05 | 0.0332 | Cardiometabolic |
| Secretogranin-3 | Q8WXD2 | -0.058 | 0.014 | 2.8e-05 | 0.0345 | Inflammation |
| Pseudokinase FAM20A | Q96MK3 | 0.051 | 0.012 | 2.93e-05 | 0.0361 | Cardiometabolic |
| Advanced glycosylation end product-specific receptor | Q15109 | -0.041 | 0.01 | 3e-05 | 0.037 | Inflammation |
| Angiopoietin-related protein 2 | Q9UKU9 | 0.042 | 0.01 | 3.02e-05 | 0.0372 | Inflammation |
| Interleukin-10 receptor subunit beta | Q08334 | 0.067 | 0.016 | 3.7e-05 | 0.0455 | Inflammation |
| Antithrombin-III | P01008 | -0.098 | 0.023 | 3.92e-05 | 0.0483 | Inflammation |
| Na(+)/H(+) exchange regulatory cofactor NHE-RF3 | Q5T2W1 | 0.044 | 0.006 | 8.24e-14 | 1.01e-10 | Inflammation |
| Sialoadhesin | Q9BZZ2 | 0.068 | 0.011 | 8.32e-10 | 1.02e-06 | Inflammation |
| Ketohexokinase | P50053 | 0.047 | 0.007 | 8.45e-10 | 1.04e-06 | Cardiometabolic |
| Leukocyte-associated immunoglobulin-like receptor 1 | Q6GTX8 | 0.082 | 0.012 | 8.41e-11 | 1.04e-07 | Inflammation |
| WAP, Kazal, immunoglobulin, Kunitz and NTR domain-containing protein 2 | Q8TEU8 | -0.072 | 0.012 | 8.64e-09 | 1.06e-05 | Inflammation |
| Carbonic anhydrase 5A, mitochondrial | P35218 | 0.025 | 0.004 | 9.02e-10 | 1.11e-06 | Cardiometabolic |
| Creatine kinase B-type | P12277 | -0.057 | 0.007 | 9.04e-16 | 1.11e-12 | Cardiometabolic |
| Proline-rich acidic protein 1 | Q96NZ9 | 0.079 | 0.009 | 9.55e-17 | 1.18e-13 | Cardiometabolic |
| Scavenger receptor cysteine-rich domain-containing group B protein | Q8WTU2 | 0.021 | 0.003 | 9.88e-11 | 1.22e-07 | Cardiometabolic |
| Bile salt sulfotransferase | Q06520 | 0.031 | 0.005 | 1.03e-08 | 1.27e-05 | Inflammation |
| Interleukin-18 receptor 1 | Q13478 | 0.081 | 0.011 | 1.03e-11 | 1.27e-08 | Inflammation |
| Insulin-like growth factor-binding protein 1 | P08833 | -0.041 | 0.004 | 1.05e-17 | 1.29e-14 | Cardiometabolic |
| BPI fold-containing family B member 2 | Q8N4F0 | 0.028 | 0.005 | 1.13e-08 | 1.39e-05 | Cardiometabolic |
| Galectin-9 | O00182 | 0.08 | 0.013 | 1.17e-09 | 1.44e-06 | Inflammation |
| Afamin | P43652 | 0.106 | 0.014 | 1.17e-13 | 1.44e-10 | Inflammation |
| Beta-glucuronidase | P08236 | 0.038 | 0.006 | 1.18e-10 | 1.46e-07 | Cardiometabolic |
| Interleukin-1 receptor antagonist protein | P18510 | 0.061 | 0.005 | 1.2e-25 | 1.48e-22 | Inflammation |
| Tumor necrosis factor receptor superfamily member 11A | Q9Y6Q6 | 0.069 | 0.01 | 1.23e-11 | 1.51e-08 | Inflammation |
| Complement factor H | P08603 | 0.131 | 0.018 | 1.24e-12 | 1.53e-09 | Inflammation |
| Galectin-3-binding protein | Q08380 | 0.065 | 0.009 | 1.29e-11 | 1.58e-08 | Inflammation |
| Scavenger receptor cysteine-rich type 1 protein M130 | Q86VB7 | 0.052 | 0.009 | 1.4e-08 | 1.73e-05 | Cardiometabolic |
| Pigment epithelium-derived factor | P36955 | 0.186 | 0.023 | 1.51e-14 | 1.86e-11 | Inflammation |
| Retinoic acid receptor responder protein 2 | Q99969 | 0.063 | 0.01 | 1.46e-09 | 1.8e-06 | Cardiometabolic |
| Collagen alpha-3(VI) chain | P12111 | 0.078 | 0.013 | 8.15e-09 | 1e-05 | Cardiometabolic |
| Leptin | P41159 | 0.052 | 0.003 | 1.66e-51 | 2.04e-48 | Cardiometabolic |
| Galectin-1 | P09382 | 0.086 | 0.014 | 1.84e-09 | 2.27e-06 | Cardiometabolic |
| Serum paraoxonase/lactonase 3 | Q15166 | -0.142 | 0.016 | 1.86e-16 | 2.29e-13 | Cardiometabolic |
| Spondin-2 | Q9BUD6 | 0.085 | 0.015 | 1.9e-08 | 2.34e-05 | Cardiometabolic |
| ADAMTS-like protein 2 | Q86TH1 | 0.073 | 0.011 | 1.91e-10 | 2.35e-07 | Cardiometabolic |
| Leukocyte immunoglobulin-like receptor subfamily B member 4 | Q8NHJ6 | 0.063 | 0.011 | 2.03e-08 | 2.5e-05 | Inflammation |
| Serum amyloid P-component | P02743 | 0.063 | 0.011 | 2.12e-08 | 2.62e-05 | Inflammation |
| Growth/differentiation factor 15 | O43184 | 0.069 | 0.009 | 2.18e-13 | 2.68e-10 | Inflammation |
| Prostaglandin reductase 1 | Q14914 | 0.032 | 0.005 | 2.29e-11 | 2.82e-08 | Cardiometabolic |
| Ribonuclease pancreatic | P07998 | 0.119 | 0.016 | 2.32e-12 | 2.86e-09 | Inflammation |
| Insulin-like growth factor-binding protein 2 | P18065 | -0.057 | 0.006 | 2.47e-17 | 3.04e-14 | Cardiometabolic |
| Glutathione S-transferase A1 | P08263 | 0.031 | 0.003 | 2.51e-17 | 3.09e-14 | Cardiometabolic |
| All-trans-retinol dehydrogenase [NAD(+)] ADH4 | P08319 | 0.036 | 0.005 | 2.59e-12 | 3.19e-09 | Cardiometabolic |
| 2-iminobutanoate/2-iminopropanoate deaminase | P52758 | 0.051 | 0.006 | 2.59e-15 | 3.19e-12 | Inflammation |
| Zinc transporter ZIP5 | Q6ZMH5 | 0.043 | 0.007 | 2.64e-09 | 3.26e-06 | Inflammation |
| Matrix remodeling-associated protein 8 | Q9BRK3 | -0.072 | 0.013 | 2.66e-08 | 3.27e-05 | Inflammation |
| Growth hormone receptor | P10912 | 0.091 | 0.011 | 3.06e-14 | 3.77e-11 | Inflammation |
| Cathepsin D | P07339 | 0.075 | 0.01 | 3.09e-13 | 3.81e-10 | Cardiometabolic |
| Liver carboxylesterase 1 | P23141 | 0.035 | 0.005 | 3.15e-12 | 3.88e-09 | Cardiometabolic |
| All-trans-retinol dehydrogenase | P00325 | 0.033 | 0.004 | 3.37e-12 | 4.15e-09 | Inflammation |
| Apolipoprotein D | P05090 | -0.072 | 0.012 | 3.39e-09 | 4.18e-06 | Inflammation |
| Beta-ureidopropionase | Q9UBR1 | 0.025 | 0.004 | 3.57e-10 | 4.39e-07 | Cardiometabolic |
| Protein turtle homolog A | Q9P2J2 | 0.036 | 0.004 | 3.59e-18 | 4.42e-15 | Cardiometabolic |
| NHL repeat-containing protein 3 | Q5JS37 | 0.11 | 0.016 | 3.62e-11 | 4.46e-08 | Inflammation |
| Sex hormone-binding globulin | P04278 | -0.053 | 0.008 | 3.79e-11 | 4.67e-08 | Inflammation |
| Low-density lipoprotein receptor | P01130 | 0.048 | 0.009 | 3.88e-08 | 4.78e-05 | Cardiometabolic |
| Protein disulfide isomerase CRELD1 | Q96HD1 | 0.095 | 0.014 | 3.92e-11 | 4.82e-08 | Inflammation |
| Cadherin-2 | P19022 | 0.068 | 0.012 | 4.36e-08 | 5.37e-05 | Cardiometabolic |
| Complement C3 | P01024 | 0.071 | 0.01 | 4.48e-11 | 5.52e-08 | Inflammation |
| Apolipoprotein F | Q13790 | -0.148 | 0.02 | 4.77e-13 | 5.88e-10 | Inflammation |
| E-selectin | P16581 | 0.042 | 0.008 | 5.13e-08 | 6.32e-05 | Cardiometabolic |
| Pterin-4-alpha-carbinolamine dehydratase | P61457 | 0.047 | 0.007 | 5.16e-11 | 6.36e-08 | Inflammation |
| High affinity immunoglobulin alpha and immunoglobulin mu Fc receptor | Q8WWV6 | 0.042 | 0.006 | 5.2e-13 | 6.4e-10 | Cardiometabolic |
| Sialic acid-binding Ig-like lectin 7 | Q9Y286 | 0.086 | 0.014 | 5.7e-09 | 7.03e-06 | Cardiometabolic |
| Hepatocyte growth factor | P14210 | 0.07 | 0.01 | 5.92e-12 | 7.29e-09 | Inflammation |
| Fatty acid-binding protein, liver | P07148 | 0.027 | 0.004 | 5.96e-09 | 7.34e-06 | Inflammation |
| Complement factor I | P05156 | 0.152 | 0.025 | 6.06e-09 | 7.47e-06 | Inflammation |
| Phospholipase A2 | P04054 | -0.048 | 0.009 | 6.07e-08 | 7.48e-05 | Cardiometabolic |
| Follistatin-related protein 3 | O95633 | 0.081 | 0.013 | 6.33e-10 | 7.8e-07 | Inflammation |
| CD59 glycoprotein | P13987 | 0.12 | 0.02 | 6.41e-09 | 7.9e-06 | Cardiometabolic |
| Fatty acid-binding protein, adipocyte | P15090 | 0.073 | 0.007 | 6.97e-23 | 8.59e-20 | Cardiometabolic |
| Basement membrane-specific heparan sulfate proteoglycan core protein | P98160 | 0.112 | 0.018 | 7.27e-10 | 8.95e-07 | Cardiometabolic |
| Microfibrillar-associated protein 5 | Q13361 | 0.069 | 0.012 | 7.78e-09 | 9.59e-06 | Cardiometabolic |
| Aminoacylase-1 | Q03154 | 0.052 | 0.007 | 7.83e-14 | 9.64e-11 | Cardiometabolic |
| Semaphorin-3F | Q13275 | 0.106 | 0.018 | 8.07e-09 | 9.94e-06 | Cardiometabolic |

**Caption:** significant association results displayed from the use of LME model: Change_BMI ~ Sex + Age at blood sample + Protein + BMI_baseline + (1|Family ID) where changes in BMI is the outcome and the protein level at ~62 years old, sex, the age of blood sampling and the baseline BMI (~24 years old) as fixed effects. **Abbreviations:** SE: Standard error.

**Supplementary table 4:** Linear mixed-effects models to assess associations of proteins at ~62 years old with BMI changes during adulthood (24 to 62 years old).

|  |  |  |  | **p value** | | **Protein** |
| --- | --- | --- | --- | --- | --- | --- |
| **Proteindescription** | **Protein ID** | **Estimate** | **SE** | **Nominal** | **Bonferroni** | **panel** |
| Leptin | P41159 | 0,039841 | 0,002664 | 2,36E-35 | 2,90E-32 | Cardiometabolic |
| Fatty acid-binding protein, adipocyte | P15090 | 0,048604 | 0,00645 | 1,14E-12 | 1,40E-09 | Cardiometabolic |
| Interleukin-1 receptor antagonist protein | P18510 | 0,041266 | 0,005556 | 2,29E-12 | 2,82E-09 | Inflammation |
| Insulin-like growth factor-binding protein 2 | P18065 | -0,04031 | 0,00579 | 3,52E-11 | 4,34E-08 | Cardiometabolic |
| Insulin-like growth factor-binding protein 1 | P08833 | -0,02827 | 0,004105 | 5,55E-11 | 6,84E-08 | Cardiometabolic |
| Creatine kinase B-type | P12277 | -0,04053 | 0,005983 | 1,07E-10 | 1,31E-07 | Cardiometabolic |
| Sex hormone-binding globulin | P04278 | -0,04146 | 0,006639 | 2,11E-09 | 2,60E-06 | Inflammation |
| Proline-rich acidic protein 1 | Q96NZ9 | 0,047757 | 0,00807 | 1,20E-08 | 1,48E-05 | Cardiometabolic |
| Growth hormone receptor | P10912 | 0,058409 | 0,009905 | 1,35E-08 | 1,67E-05 | Inflammation |
| Serum paraoxonase/lactonase 3 | Q15166 | -0,0963 | 0,017241 | 6,83E-08 | 8,41E-05 | Cardiometabolic |
| Tissue-type plasminogen activator | P00750 | 0,027459 | 0,005075 | 1,60E-07 | 0,000197 | Cardiometabolic |
| Protein turtle homolog A | Q9P2J2 | 0,019812 | 0,003696 | 2,10E-07 | 0,000259 | Cardiometabolic |
| Phospholipid transfer protein | P55058 | -0,05719 | 0,010869 | 3,49E-07 | 0,00043 | Cardiometabolic |
| Pigment epithelium-derived factor | P36955 | 0,110446 | 0,021757 | 8,04E-07 | 0,00099 | Inflammation |
| Disintegrin and metalloproteinase domain-containing protein 12 | O43184 | 0,043626 | 0,008622 | 8,71E-07 | 0,001073 | Inflammation |
| 2-iminobutanoate/2-iminopropanoate deaminase | P52758 | 0,03051 | 0,006062 | 1,06E-06 | 0,001305 | Inflammation |
| Golgi-associated kinase 1A | Q9UFP1 | 0,036852 | 0,007546 | 1,96E-06 | 0,002418 | Cardiometabolic |
| Galectin-9 | O00182 | 0,054034 | 0,011177 | 2,46E-06 | 0,003032 | Inflammation |
| Scavenger receptor cysteine-rich domain-containing group B protein | Q8WTU2 | 0,013885 | 0,002893 | 2,96E-06 | 0,003643 | Cardiometabolic |
| Carbonic anhydrase 5A, mitochondrial | P35218 | 0,016813 | 0,003557 | 4,03E-06 | 0,004963 | Cardiometabolic |
| Zinc transporter ZIP5 | Q6ZMH5 | 0,029943 | 0,006349 | 4,19E-06 | 0,005162 | Inflammation |
| Glutathione S-transferase A1 | P08263 | 0,016872 | 0,003579 | 4,37E-06 | 0,005383 | Cardiometabolic |
| WAP, Kazal, immunoglobulin, Kunitz and NTR domain-containing protein 2 | Q8TEU8 | -0,04882 | 0,010392 | 4,93E-06 | 0,006071 | Inflammation |
| Low-density lipoprotein receptor | P01130 | 0,034603 | 0,007534 | 7,24E-06 | 0,008914 | Cardiometabolic |
| CD59 glycoprotein | P13987 | 0,08474 | 0,018654 | 9,05E-06 | 0,011153 | Cardiometabolic |
| Apolipoprotein D | P05090 | -0,04685 | 0,010295 | 9,11E-06 | 0,011228 | Inflammation |
| Semaphorin-3F | Q13275 | 0,069431 | 0,015304 | 9,33E-06 | 0,011498 | Cardiometabolic |
| NHL repeat-containing protein 3 | Q5JS37 | 0,067235 | 0,015 | 1,18E-05 | 0,014526 | Inflammation |
| Ribonuclease pancreatic | P07998 | 0,067757 | 0,015307 | 1,49E-05 | 0,018332 | Inflammation |
| Uromodulin | P07911 | -0,0319 | 0,007257 | 1,77E-05 | 0,021796 | Cardiometabolic |
| Neurexophilin-3 | O95157 | -0,04645 | 0,010585 | 1,79E-05 | 0,022055 | Inflammation |
| Interleukin-18 receptor 1 | Q13478 | 0,045494 | 0,010426 | 1,94E-05 | 0,023916 | Inflammation |
| Leukocyte immunoglobulin-like receptor subfamily B member 4 | Q8NHJ6 | 0,04071 | 0,009366 | 2,11E-05 | 0,025985 | Inflammation |
| Serum amyloid P-component | P02743 | 0,040887 | 0,009419 | 2,14E-05 | 0,026326 | Inflammation |
| SLIT and NTRK-like protein 1 | Q96PX8 | -0,04738 | 0,010921 | 2,18E-05 | 0,026856 | Inflammation |
| Secretoglobin family 3A member 1 | Q96QR1 | -0,04407 | 0,01018 | 2,25E-05 | 0,027674 | Inflammation |
| Phospholipase A2 | P04054 | -0,03207 | 0,007531 | 3,01E-05 | 0,037138 | Cardiometabolic |
| Afamin | P43652 | 0,054438 | 0,01281 | 3,12E-05 | 0,038437 | Inflammation |
| Complement C3 | P01024 | 0,039021 | 0,009283 | 3,88E-05 | 0,04777 | Inflammation |
| Sialoadhesin | Q9BZZ2 | 0,042594 | 0,010167 | 4,01E-05 | 0,049401 | Inflammation |

**Caption:** significant association results displayed from the use of LME model: Change_BMI ~ Sex + Age at blood sample + Protein + BMI_baseline + (1|Family ID) after removing samples with BMI>30 kg/m2. **Abbreviations:** SE: Standard error.

**Supplementary table 5:** Linear mixed-effects models to assess associations of BMI fluctuation and proteins at ~62 years old.

| **Protein Description** | **Protein ID** | **Estimate** | **SE** | **p value** | | **Protein panel** |
| --- | --- | --- | --- | --- | --- | --- |
|  |  |  |  | **Nominal** | **Bonferroni** |  |
| Leptin receptor | P48357 | 0,76 | 0,34 | 0,03 | 1 | Cardiometabolic |
| Interleukin-1 receptor antagonist protein | P18510 | 0,35 | 0,15 | 0,02 | 1 | Inflammation |
| Fatty acid-binding protein, adipocyte | P15090 | 0,31 | 0,19 | 0,1 | 1 | Cardiometabolic |
| Angiopoietin-related protein 4 | Q9BY76 | 0,7 | 0,23 | 2,10E-03 | 1 | Inflammation |
| Disintegrin and metalloproteinase domain-containing protein 12 | O43184 | 0,43 | 0,21 | 0,04 | 1 | Inflammation |
| Na(+)/H(+) exchange regulatory cofactor NHE-RF3 | Q5T2W1 | 0,36 | 0,13 | 6,50E-03 | 1 | Inflammation |
| Growth/differentiation factor 15 | Q99988 | 0,59 | 0,22 | 7,00E-03 | 1 | Inflammation |
| Bile salt sulfotransferase | Q06520 | 0,34 | 0,11 | 2,80E-03 | 1 | Inflammation |
| 2-iminobutanoate/2-iminopropanoate deaminase | P52758 | 0,35 | 0,15 | 0,02 | 1 | Inflammation |
| Coiled-coil domain-containing protein 80 | Q76M96 | 0,82 | 0,21 | 1,23E-04 | 0,15 | Cardiometabolic |
| Interleukin-10 receptor subunit beta | Q08334 | 0,97 | 0,33 | 3,21E-03 | 1 | Inflammation |
| Aflatoxin B1 aldehyde reductase member 4 | Q8NHP1 | 0,29 | 0,1 | 2,94E-03 | 1 | Inflammation |
| Pterin-4-alpha-carbinolamine dehydratase | P61457 | 0,41 | 0,16 | 0,01 | 1 | Inflammation |
| Scavenger receptor cysteine-rich type 1 protein M130 | Q86VB7 | 0,54 | 0,19 | 5,26E-03 | 1 | Cardiometabolic |
| Pantetheinase | O95497 | 0,31 | 0,12 | 9,44E-03 | 1 | Inflammation |
| Glutathione S-transferase A1 | P08263 | 0,15 | 0,09 | 0,08 | 1 | Cardiometabolic |
| E-selectin | P16581 | 0,34 | 0,16 | 0,04 | 1 | Cardiometabolic |

**Caption:** significant association results displayed from the use of LME model: BMI_fluctuation ~ Sex + Age at blood sample + Protein + (1|Family ID) after adding BMI baseline and changes in BMI during adulthood (i.e. slope) as a fixed effect as part of sensitivity test. **Abbreviations:** SE: Standard error.

**Supplementary table 6:** Linear mixed-effects models to assess associations of BMI changes with polygenic risk score (PRS) for BMI.

| **Variables comparison** | **Estimate** | **SE** | **p value** | **95%CI** | |
| --- | --- | --- | --- | --- | --- |
|  |  |  |  | **LB** | **UB** |
| BMI changes – PRS_BMI_ | 0,01 | 4,80E-03 | 0,01 | 2,00E-03 | 0,02 |
| BMI fluctuations – PRS_BMI_ | -0,09 | 0,09 | 0,33 | -0,22 | 0,35 |

**Caption:** Significant association results displayed from the use of LME model: Change_BMI (or BMI_fluctuation) ~ Sex + Age at blood sample + PRS_BMI_ + BMI_baseline + (1|Family ID) where changes in BMI is the outcome and the protein level at ~62 years old, sex, the age of blood sampling, the baseline BMI (~24 years old) and the Polygenic risk score were fixed effects. **Abbreviations:** SE: Standard error; LB: Lower bound; UB: Upper bound.

**Supplementary table 7:** Linear regression models used in within-pair analysis to assess which of the previously identified associations between proteins and BMI changes during adulthood remained significant when controlling for all genetic confounding.

| **Protein Description** | **Protein ID** | **Estimate** | **SE** | **p value** | | **Protein**  **panel** |
| --- | --- | --- | --- | --- | --- | --- |
|  |  |  |  | **Nominal** | **Bonferroni** |  |
| Leptin | P41159 | 0,05 | 0,007991747 | 7,78896E-07 | 0,000105151 | Cardiometabolic |
| Apolipoprotein F | Q13790 | -0,31 | 0,061949825 | 1,19526E-05 | 0,001613605 | Inflammation |
| Growth hormone receptor | P10912 | 0,14 | 0,028998154 | 2,97011E-05 | 0,004009651 | Inflammation |
| High affinity immunoglobulin alpha and immunoglobulin mu Fc receptor | Q8WWV6 | 0,07 | 0,016135343 | 4,79332E-05 | 0,006470979 | Cardiometabolic |
| Insulin-like growth factor-binding protein 2 | P18065 | -0,07 | 0,015954507 | 8,74034E-05 | 0,011799463 | Cardiometabolic |
| Creatine kinase B-type | P12277 | -0,07 | 0,015566715 | 9,99464E-05 | 0,013492762 | Cardiometabolic |
| Somatotropin | P01241 | -0,02 | 0,004534068 | 0,000126662 | 0,01709935 | Cardiometabolic |
| BPI fold-containing family B member 1 | Q8TDL5 | -0,07 | 0,01731678 | 0,000271315 | 0,036627512 | Cardiometabolic |
| Ectonucleotide pyrophosphatase/phosphodiesterase family member 7 | Q6UWV6 | 0,06 | 0,016291831 | 0,000293549 | 0,03962906 | Inflammation |
| Insulin-like growth factor-binding protein 1 | P08833 | -0,04 | 0,009657006 | 0,000351033 | 0,047389496 | Cardiometabolic |

**Caption:** significant association results displayed from the use of Linear regression model: ∆ Change_BMI ~ sex of the pair + Age at blood sample + ∆ Protein levels + mean Baseline_BMI. **Abbreviations:** SE: Standard error.

**Supplementary table 8:** Linear regression models used in within-pair analysis to assess which of the previously identified associations between proteins and BMI fluctuation during adulthood remained significant when controlling for genetic confounding.

| **Protein Description** | **Protein ID** | **Estimate** | **SE** | **p value** | | **Protein panel** |
| --- | --- | --- | --- | --- | --- | --- |
|  |  |  |  | **Nominal** | **Bonferroni** |  |
| Na(+)/H(+) exchange regulatory cofactor NHE-RF3 | Q5T2W1 | 1,128874475 | 0,325847505 | 0,00116087 | 0,018573917 | Inflammation |
| E-selectin | P16581 | 1,972037277 | 0,631986687 | 0,003116475 | 0,049863598 | Cardiometabolic |

**Caption:** significant association results displayed from the use of Linear regression model: ∆ BMI_fluctuation ~ sex of the pair + Age at blood sample + ∆ Protein levels. **Abbreviations:** SE: Standard error.
